# Advancing the communication of genetic risk for cardiometabolic diseases: A critical interpretive synthesis

**DOI:** 10.1101/2023.05.12.23289038

**Authors:** Jing Hui Law, Najia Sultan, Sarah Finer, Nina Fudge

**Affiliations:** Wolfson Institute of Population Health, Queen Mary University of London, London, United Kingdom; Barts Health NHS Trust, London, United Kingdom

**Keywords:** genetic risk, family history, cognitive appraisals, risk perceptions, polygenic risk scores, health behaviours, cardiometabolic diseases, type 2 diabetes, cardiovascular disease, obesity

## Abstract

**Background:** Genetics play an important role in risk for cardiometabolic diseases—including type 2 diabetes, cardiovascular disease and obesity. Existing research has explored the clinical utility of genetic risk tools such as polygenic risk scores—and whether interventions communicating genetic risk information using these tools can impact on individuals’ cognitive appraisals of disease risk and/or preventative health behaviours. Previous systematic reviews exploring the evidence base suggest mixed results. To expand current understanding and address knowledge gaps, we undertook a reflexive method of evidence synthesis to the literature—questioning the theoretical basis behind current interventions that communicate genetic risk information and exploring how the effects of genetic risk tools can be fully harnessed for cardiometabolic diseases.

**Methods:** We obtained 188 records from a combination of database, website and grey literature searches—supplemented with reference chaining and expert subject knowledge within the review team. Using pre-defined critical interpretive synthesis methods, quantitative and qualitative evidence was synthesised and critiqued alongside theoretical understanding from surrounding fields of behavioural and social sciences.

**Findings:** Existing interventions communicating genetic risk information focus predominantly on the “self”, targeting individual-level cognitive appraisals, such as perceived risk and perceived behavioural control. This approach risks neglecting the role of contextual factors and upstream determinants that can reinforce individuals’ interpretations of risk. It also assumes target populations to embody an “ascetic subject of compliance”—the idea of a patient who strives to comply diligently with professional medical advice, logically and rationally adopting any recommended lifestyle changes. We developed a synthesising argument—beyond the “ascetic subject of compliance”—grounded in three major limitations of this perspective: (1) Difficulty applying existing theories/models to diverse populations; (2) The role of familial variables and (3) The need for a life course perspective.

**Conclusions:** Interventions communicating genetic risk information should account for wider influences that can affect individuals’ responses to risk at different levels—including through interactions with their family systems, socio-cultural environments and wider health provision.

**Protocol registration:** PROSPERO CRD42021289269

## Background

For most common cardiometabolic diseases such as type 2 diabetes (T2D) and cardiovascular disease (CVD), multiple factors such as genes and lifestyle interact to play a causal role in an individual’s risk (1, 2). Genomic advances over the past decade—particularly with genome-wide association studies (GWASs)—have identified the contribution of inherited variants to disease risk and allowed for the advancement of genetic risk tools such as polygenic risk scores (PRSs) (3, 4). PRSs are formed by combining multiple independent genetic risk variants associated with a certain health condition in an individual—which is in turn used to generate a score estimating their genetic risk for that particular disease (3). PRSs have been shown to be useful for the prediction of disease risk—and recent efforts have shown their potential utility in clinical contexts (3-7). For example, PRSs can help enhance the stratification and management of individuals at high risk of chronic diseases, facilitating referrals onto screening programmes, lifestyle interventions and/or preventative treatment (3, 4, 8-11).

Current evidence supports the integration of PRSs with existing clinical risk tools that are already widely used in routine care for the prediction of cardiometabolic diseases (5-9). In particular, they seem to offer benefits in identifying high-risk individuals at younger ages—prior to the development of clinical symptoms or risk factors—as PRSs are able to capture a component of risk that is fixed lifelong. This offers health services a means to identify and act on individuals’ risk more efficiently, through better allocation of preventative care, based on earlier indications of risk. However, there are major questions to ask regarding how individuals would receive, interpret and respond to genetic risk information—especially at younger ages than is typical for risk factor screening and management— and how this would be managed in routine clinical care systems. In other words, the effective implementation of PRSs relies on understanding the relevant behavioural science—to identify how genetic risk tools can exert the most direct impact on individuals receiving risk information.

Many interventions have explored the effect of communicating genetic risk information for health conditions on shifting individuals’ cognitive appraisals of disease risk (e.g. perceived risk, perceived behavioural control) and/or encouraging the adoption of preventative health behaviours (4, 12-14). Earlier studies on the prediction of breast and colon cancers have shown that providing genetic risk information can help promote patients’ screening attendance and medication adherence (13, 15). Existing systematic reviews indicate, however, that the evidence is less clear for lifestyle behaviours such as physical activity and diet, which need to be adopted and sustained across time to reduce the risk of developing cardiometabolic diseases (13, 15). This raises important questions over the value of wide-scale integration of PRSs into healthcare systems for common diseases—as their use must be determined on clear clinical utility. As such, establishing and understanding the evidence base on individual-level perceptions and behaviours towards genetic risk information is crucial to help clarify the role of genetic risk tools in clinical care—and fully leverage their application for common diseases on a population-wide basis.

We conducted a critical interpretive synthesis (CIS) to advance current understanding on the communication of genetic risk for cardiometabolic diseases. A CIS is a method of evidence synthesis known for its critical and reflexive nature—where the central feature is in adopting an investigative lens to the literature concerned (16). We applied this approach to explore the premise behind interventions that have been proposed to modify cognitive appraisals and/or health behaviours via the provision of genetic risk information. In doing so, we questioned how interventions have traditionally framed the uses and purposes of genetic risk communication, the assumptions that they have drawn from, as well as why current evidence seems to generate mixed results. This critical approach has particular strength in highlighting new and unique perspectives within a research area, allowing us to expand beyond the findings of conventional systematic reviews that are already available on the topic (1, 2, 13, 15)—and further identify potential areas for future efforts to improve risk communication. The broad aims set for this CIS were threefold. Firstly, we aimed to summarise existing evidence on cognitive appraisals that may be particularly important or salient for individuals receiving genetic risk information—how these cognitive appraisals been studied in relation to cardiometabolic diseases and whether they can, in turn, impact on individuals’ adoption of preventative health behaviours. Secondly, we aimed to investigate knowledge gaps in this research area—particularly exploring why existing interventions that target individuals’ cognitive appraisals (and/or health behaviours) via the provision of genetic risk information suggest mixed results. Finally, we aimed consolidate these findings to consider how the effects of genetic risk tools can be fully harnessed to help mitigating cardiometabolic disease risk.

## Methods

### Search strategy

We followed principles and methods first defined by Dixon-Woods et al., in their CIS conceptualising how vulnerable groups in the UK access and utilise healthcare services (16). Our literature search combined a broad number of strategies and included searching electronic databases, websites, NHS reports and reference chaining. Expertise within the multidisciplinary review team was further utilised to identify relevant work from adjacent fields not immediately or obviously relevant to the communication of genetic risk. This team comprised researchers working in the fields of psychology and behavioural sciences, anthropology and social sciences—as well as healthcare professionals in primary care.

An initial search strategy was piloted on Ovid MEDLINE in September 2021, based on search terms used in previous systematic reviews (1, 2, 13, 15) and then refined for the purposes of this CIS. Our search initially focused on evidence solely related to the communication of *genetic* risk—but this retrieved a considerable number of records on the communication of *familial* risk. Upon inspection, the review team agreed that there was substantial overlap—many cognitive appraisals implicated in the communication of genetic risk were similarly raised in research on the communication of familial risk. We thus updated our search strategy to explicitly include this body of work.

We applied the finalised search strategy (Additional file 1) to the following databases in November 2021—Ovid MEDLINE (1946 to November 2021), EMBASE (1980 to November 2021), PsycINFO (1967 to November 2021), Scopus (1960 to November 2021), Web of Science (1950 to November 2021) and the Cochrane Central Register of Controlled Trials (CENTRAL). We consulted with an academic librarian for the validation of the final search terms across these databases.

### Inclusion/exclusion criteria

A hallmark of the CIS method is to avoid appraising studies based solely on the type of design—thus allowing for a diverse and interdisciplinary body of evidence to be synthesised. We included a range of quantitative, qualitative and mixed-methods studies examining various cognitive appraisals that have been implicated in genetic/familial risk for cardiometabolic diseases (including T2D, CVD and obesity). As described in our protocol, we expected these cognitive appraisals to encompass factors such as perceived risk, perceived behavioural control and intention to engage in preventative health behaviours. Study designs ranged from interventions that combine (real or hypothetical) genetic/familial risk information with lifestyle advice to examine participants’ behavioural outcomes— to interview studies exploring participants’ thoughts about their family history of disease. Populations of interest included members of the general population, clinically at-risk individuals, first-degree relatives of patients with cardiometabolic diseases or unaffected family members who may be at risk themselves. We also included systematic reviews that fit our topic, along with potentially relevant grey literature—such as reports and policy documents, commentaries and opinion pieces, theses and dissertations, as well as conference papers and proceedings. We excluded published study protocols and studies that were incomplete and/or had no outcomes reported.

Given the broad inclusion criteria usually adopted in a CIS, we anticipated the number of eligible records to be very high from the start. Here, an exhaustive summary of all the data retrieved is not expected—since the main goal of a CIS is to generate a theoretical structure, or a conceptual framework that Dixon-Woods et al. have termed “synthesising argument” (16). Synthesising arguments are produced through detailed and iterative analysis—a process comparable to analysis processes conducted in primary qualitative research (16). It represents an overarching idea that encompasses the body of evidence described in a CIS—functioning to provide a more “insightful, formalised and generalisable” way to understand the literature (16). The flowchart of our record selection process is described in Figure 1 below. At the initial screening stage, all members of the review team screened the 378 records obtained from our database searches, based on titles and abstracts. There were 286 records deemed eligible for inclusion. We then developed and applied the following purposive sampling criteria—following the methods outlined by Dixon-Woods et al. (16), alongside a three-step purposive sampling framework adapted from the vaccination communication literature (17)—as a starting point to help us reach a manageable sampling frame for data extraction:

1. Maximum variation—Research that addressed the topic of interest in diverse settings and/or populations (e.g. in underrepresented geographic areas and/or populations);
2. Data richness—Mixed-methods and/or qualitative work that provide in-depth and conceptually rich insights into the phenomenon of interest; and
3. Match of scope—Records with the most direct relevance to our research questions.

**Figure 1.**
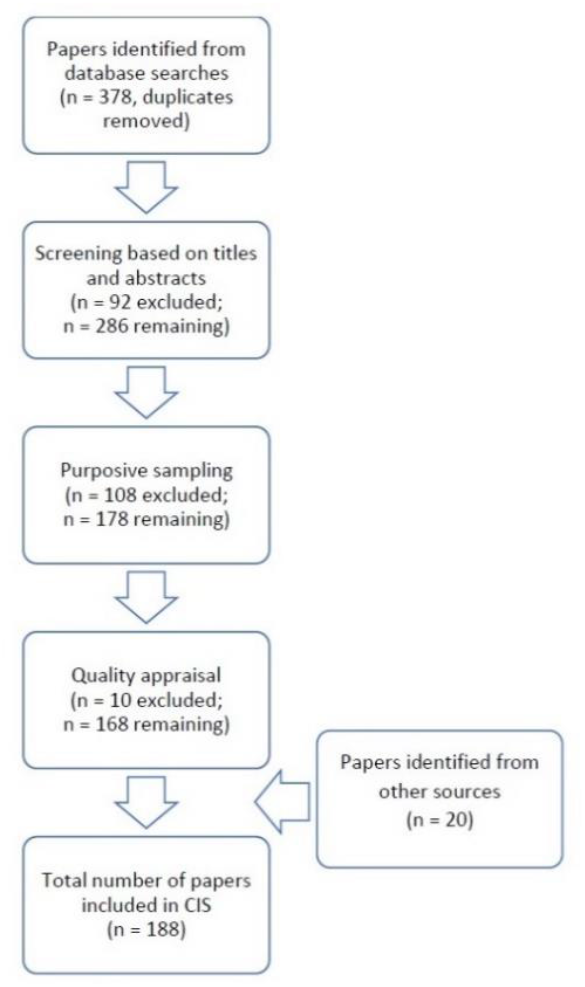
Flowchart of record selection process.

Using these criteria, the review team filtered through different subsets of the records that passed initial screening. A citation management tool was used to keep track of all records screened. We also set up a shared document to facilitate the communication of reflections and notes on the records selected for inclusion (Additional file 2). Any conflicts in decisions at this stage were resolved via further notes and discussions. This process helped us refine our initial focus onto a smaller subset of records that were deemed key to the CIS (i.e. records that fit all three of the purposive sampling criteria above).

### Quality appraisal

As a method, a CIS prioritises relevance to research questions over particular study methodologies. Whereas traditional methods of quality assessment for systematic reviews would often adopt a “hierarchy of evidence” approach—judging certain study designs or methodological standards (e.g. randomised controlled trials) as being more valuable than others (e.g. cross-sectional studies)—such an approach risks discounting important studies that may still be conceptually rich and relevant, despite their supposed methodological “inferiority” (16). Thus, to maximise the inclusion and contribution of a wider variety of work, we used the original appraisal prompts described by Dixon-Woods et al. to guide our decisions on data quality and relevance—in place of more formal quality assessment criteria (Figure 2) (16). A small number of records retrieved from our searches that did not involve primary data collection were not assessed using these appraisal prompts (e.g. reports and policy documents, commentaries and opinion pieces). Of the remaining records containing quantitative, qualitative and mixed-methods studies, we applied these prompts alongside our purposive sampling strategy to assess their overall quality. The majority met all the criteria outlined by Dixon-Woods et al.—only a small number (n = 10) were excluded on the basis of limitations related to research design and/or procedures (Figure 1; see also Additional file 2).

**Figure 2.**
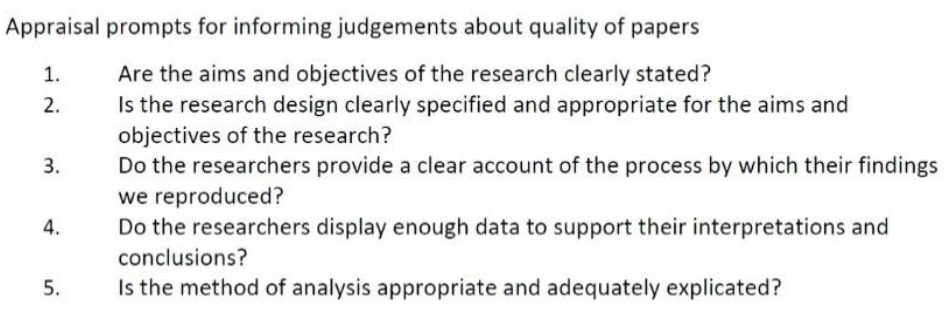
Appraisal prompts to determine paper quality for a CIS, adapted from (16).

### Data extraction

Data extraction was conducted using NVivo. We first imported the relevant paragraphs and notes from the records as data into the software—and created different nodes to represent each idea or theme we identified (example in Additional file 3). Using the NVivo node hierarchies function, we were then able to organise the coding of recurring themes and constructs across the different records and map out the relationships between them. When we began to see an emerging conceptual framework in the data, we worked to identify records from other sources that can add to, test, or elaborate the emerging analysis—and also address any conceptual gaps (Figure 1). We treated these records—and the themes derived from them—as the central point of our CIS, gradually expanding our scope and working outwards to identify more records, as the CIS evolved. Throughout these processes, ideas for generating a synthesising argument were continuously discussed between members of the multidisciplinary review team. This allowed an iterative, inductive process of analysis, synthesis and refining of the research questions to achieve theoretical saturation and generate our synthesising argument.

### Findings

This CIS involved a final total of 188 records (Additional file 4). These contained original quantitative (n = 114), qualitative (n = 30) and mixed-methods studies (n = 11), systematic reviews (n = 13), reports and policy documents (n = 4), commentaries and opinion pieces (n = 5), theses and dissertations (n = 7), conference papers and proceedings (n = 3), as well as a book chapter (n = 1). Original studies covered various geographical ranges—with the majority of them based in Northern America (n = 91) and Europe (n = 55). Further details of the included studies can be found in the supplementary material (Additional file 4).

### Understanding the communication of genetic risk in the literature

The original focus of this CIS was on exploring the various cognitive appraisals that have been studied in relation to genetic/familial risk communication for cardiometabolic diseases—including perceived risk, perceived behavioural control and behavioural intention. We identified many prominent theories in health psychology that have been applied in the research landscape (18-22). Examples include Leventhal’s Common Sense Model of Self Regulation (CSM-SR), the Health Belief Model (HBM) and the Theory of Planned Behaviour (TPB). For example, researchers have used CSM-SR to describe how threat representations that include genetic causes may lead individuals to perceive less behavioural control, thereby activating beliefs that behavioural responses may be ineffective in preventing that threat—an idea often termed by researchers as genetic fatalism (1, 12, 13, 20, 23). This provides a framework to understand why individuals sometimes adopt maladaptive responses towards health threats (24). Other theories have similarly been used by researchers as guiding frameworks—with attempts to chart a prediction of health behaviours based on networks of constructs such as perceived risk and behavioural intention (12, 25-30).

However, the evidence generated from this body of work is mixed—and at times contrary to the predictions of the theoretical frameworks behind them (12, 13, 19, 20, 30, 31). Associations between genetic causal beliefs and perceived behavioural control are not always replicated—and higher levels of perceived behavioural control do not necessarily translate into preventative health behaviours (1, 20, 23). In such cases, it is common for any lack of observable changes in participants’ psychological and/or behavioural responses following exposure to genetic risk information to be attributed as a maladaptive reaction, brought about by perceptions of uncontrollability or unpreventability (1, 13). The premise here is that changes in individuals’ cognitive appraisals should logically follow from risk information—and strategies to cope with the information subsequently adopted. Conversely, if participants’ scores on a measure such as perceived risk are not significantly changed post-intervention—the tendency is to conclude that these interventions simply do not “work”.

To untangle these gaps in understanding, we drew upon the concept of “auxiliary assumptions” from theoretical psychology (32, 33). For interventions to “work”—whether in altering cognitive appraisals or influencing behaviour change—the conditions for them to be successful first need to be satisfied (32, 33). For example, if a behaviour change technique has only been tested in older populations, applying it to children may not bring about the same effect. In the latter scenario, it may not mean that the intervention is ineffective per se; rather the conditions for it to be effective—its auxiliary assumptions—have simply not been met. Similarly, an intervention communicating genetic risk information that does not appear to “change” individuals’ perceived risk does not necessarily mean that it is not effective. If factors that are salient or important to an individual’s risk perceptions were never targeted, it is unlikely that they will be informed or shaped by interventions aiming to address this construct—thus making them unlikely to “work” as preventative strategies. Individuals may be drawing upon their own pre-established notions about personal risk—which in turn can be informed by other cognitive, emotional, social and/or environmental resources that are insufficiently accounted for by theoretical models. These may then further interact with individuals’ cognitive appraisals to determine behavioural responses. In such cases, attempting to elicit and/or alter reactions solely at the level of the individual may be insufficient. Instead, there is first a need to consider what the idea of risk means to an individual—and how it is relevant to them—to help ensure that interventions are designed to tap into participants’ understandings and interpretations of risk in the first place. Such an approach can aid in translating the idea of risk into a personalised form that is meaningful to individuals and fits with their current views and/or lifestyles.

### Synthesising argument—beyond the “ascetic subject of compliance”

As such, our analysis crucially indicated a need to focus beyond such self-/individual-oriented perspectives of cognitive appraisals. With the principle narrative placing focus on individual responsibility and personal control, the tendency is for interventions communicating genetic risk information to presuppose their target populations embodying the “ascetic subject of compliance” (34). This concept was first introduced by anthropologist Ian Whitmarsh, who discussed it within the context of global health interventions for chronic diseases. He offered a critique of the biomedical discourse in this field—which necessitates and expects patients to be “disciplined” and “compliant”, taking lifelong responsibility over their long-term treatments (34). In this CIS, we argue that similar assumptions are held in the field of genetic risk communication—presupposing the idea of an individual who strives to comply diligently with professional medical advice; who can self-monitor and adopt recommended lifestyle changes that logically follow on from interventions (34). Such a view neglects the crucial role of various upstream determinants and contextual factors that can influence decision-making processes—and that are themselves risk factors of disease.

We developed a critique, followed by the generation of a synthesising argument, constructed around a set of knowledge gaps that we have observed in the field: (1) Difficulty applying existing theories/models to diverse populations; (2) The role of familial variables and (3) The need for a life course perspective (Figure 3, Table 1). As we illustrate these knowledge gaps over the following sections, we highlight the importance of considering *beyond* the “ascetic subject of compliance”. Genetic risk should be seen as “inherited” alongside wider cultural, social and psychological variables that shape an individual—and interactions can exist between individuals, their micro-contexts (unique family dynamics; e.g. family experience with the condition, family support in relation to healthy lifestyle behaviours) and macro-contexts (upstream determinants; e.g. local socio-cultural context, living in a disadvantaged area). These are aspects that require more mainstream attention, as they can reinforce individuals’ interpretations of health threats and/or the meanings assigned to risk.

**Table 1.**
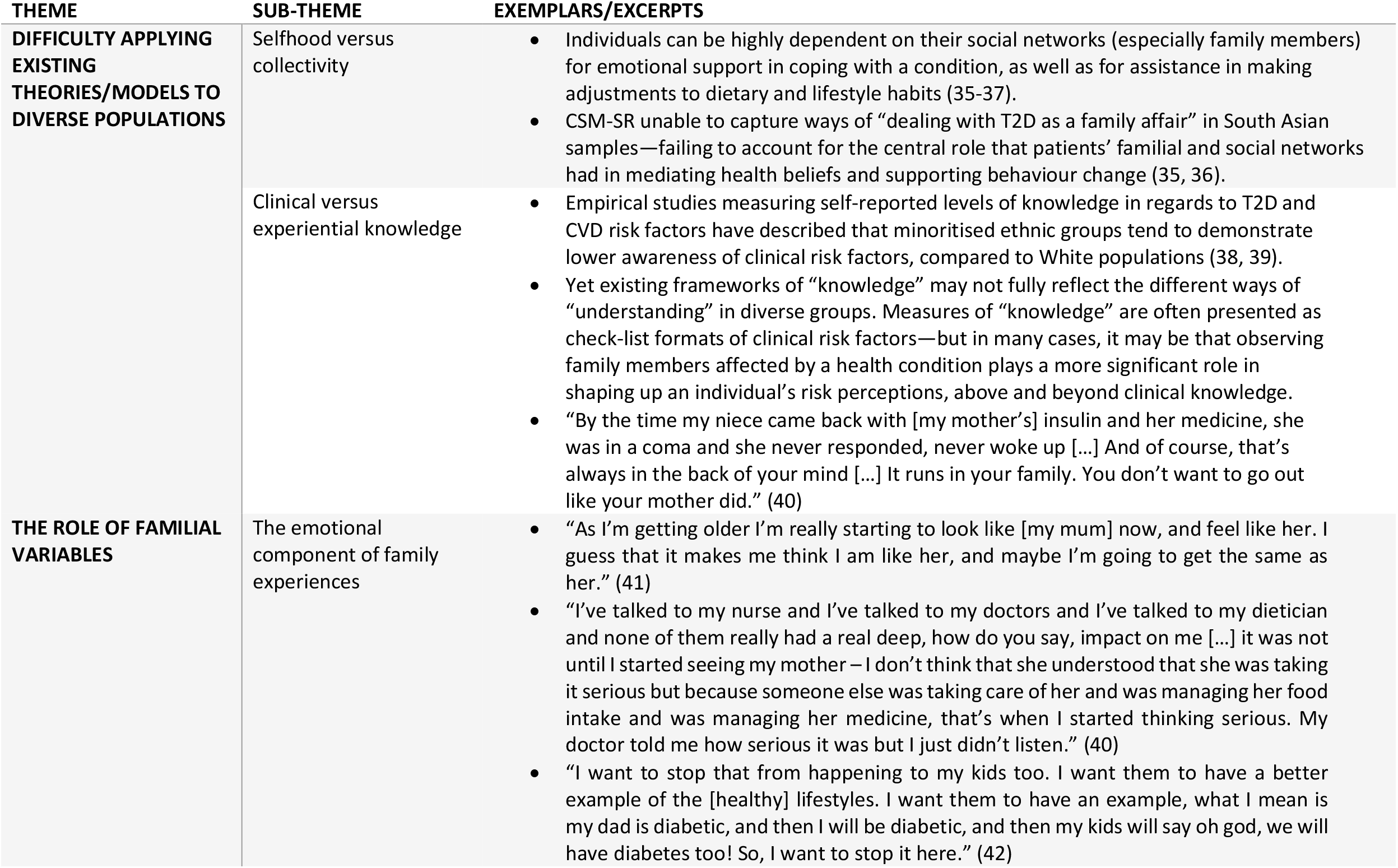
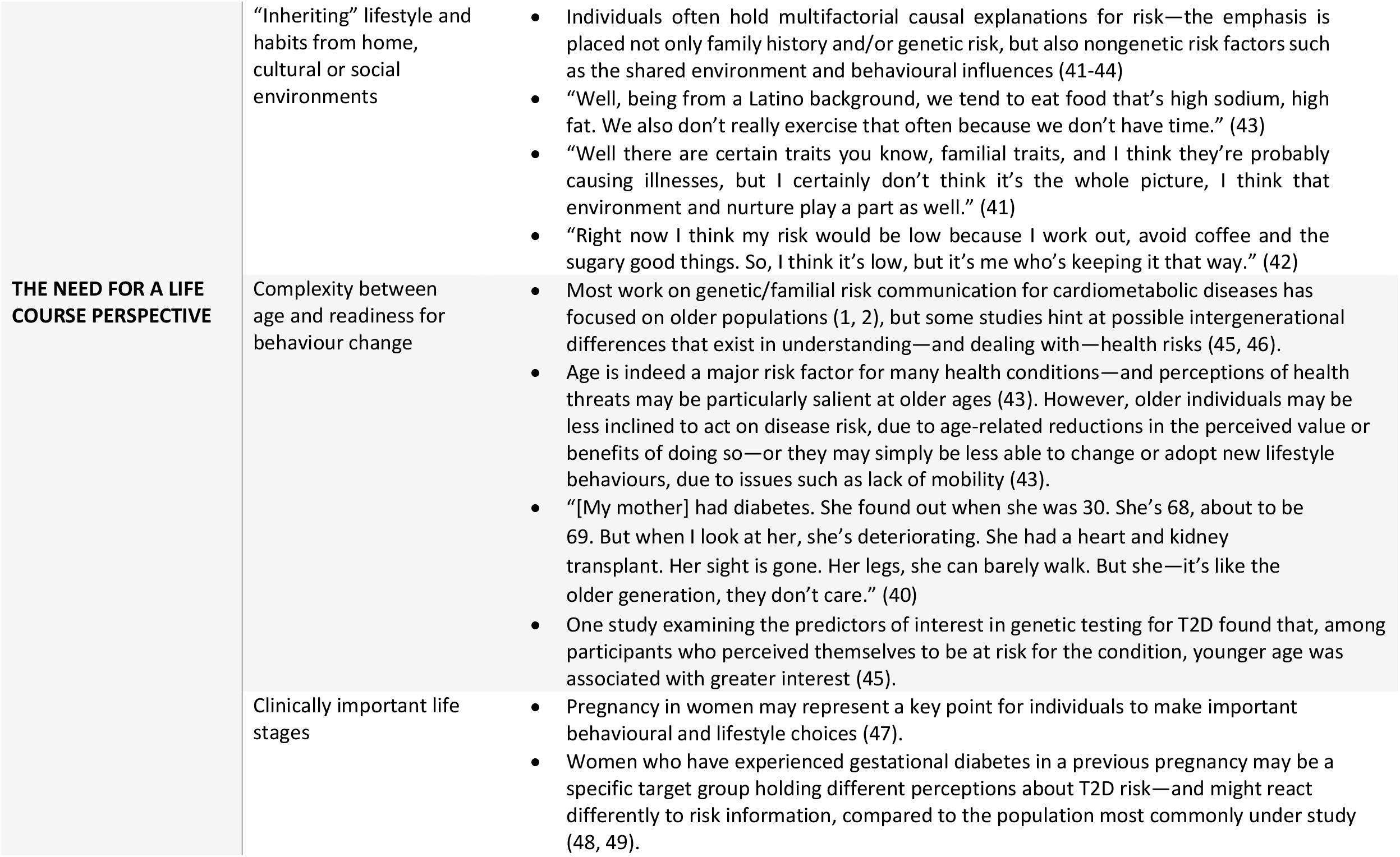
Synthesising argument—beyond the “ascetic subject of compliance”.

**Figure 3.**
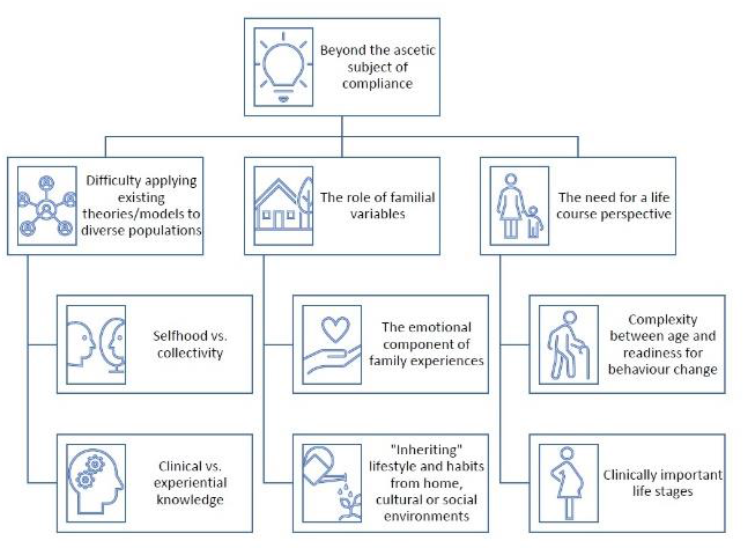
Synthesising argument—beyond the “ascetic subject of compliance”.

### Difficulty applying existing theories/models to diverse populations

Individualistic perspectives of threat and coping representations are largely built on Western ideas of “selfhood” and individuality (34). Accordingly, the first major limitation we identified in the literature was that many theoretical domains or concepts borne out of these perspectives may not hold true for diverse communities. For instance, a mixed-methods study attempting to apply CSM-SR to explore beliefs surrounding T2D self-management in British South Asians found that elements of the model failed to allow a full understanding of illness beliefs in the study sample (35, 36) (Table 1). For many people, how a health condition is understood and experienced is reliant on their immediate network of support, largely consisting of their family. This is perhaps a consequence of—and reinforced by— the central importance of family in many diverse populations within collectivist cultures and/or the salience of the condition due to its high prevalence in certain communities (35-37). Along similar lines, a qualitative study exploring diabetes illness representations in a predominantly Black sample found that participants frequently related their own concerns to family members’ disease-related complications or death (40). Furthermore, participants would often leverage their family experience into a form of motivation to avoid similar health complications (Table 1). This form of experiential knowledge—drawing from cultural and familial experiences and existing beyond traditionally conceptualised clinical risk factors—are aspects that empirical studies taking positivist approaches may not be able to capture or quantify. Such differences in understanding can then inform threat representations in unique ways, translating into variations in coping mechanisms between diverse groups. The emphasis on the role of the family in these processes—particularly in non-White populations—illustrates the limits of current understanding.

### The role of familial variables

The second major limitation included various descriptions about the emotional impact that family experiences can have in underpinning and heightening the sense of risk (41-44, 50). One’s perceived closeness with affected family members—the strength of their emotional bond and relationship, at times even their perceived likeness with the family member—informs the salience and vulnerability associated with a health condition (Table 1). Affective pathways to cognitive appraisals have not been explored extensively, but it is likely that close experiences with a condition, along with the associated emotional impact, can resonate beyond any advice given by healthcare professionals (Table 1). This can further inform individuals’ willingness and readiness to engage in preventative health behaviours—representing a valuable point of intervention (Table 1). Qualitative research also show that participants often discuss risk as being “inherited” via lifestyle and habits from home, cultural or social environments (41-44). Genes are not the only aspect viewed as being transmitted across generations—rather, familial variables such as lifestyle and levels of physical activity (and even dietary habits embedded in broader cultural contexts) are seen as passing down generationally and affecting health (43) (Table 1). This illustrates the complex mental strategies surrounding nature and nurture that can be implicated in individuals making sense of their own risk. Family history, modifiable risk factors such as diet and physical activity, as well as wider environmental factors such as familial, social and cultural contexts are all nested and interact with one another—and it is this interaction that can change the course of an individual’s (actual and perceived) risk.

### The need for a life course perspective

At present, most research on genetic/familial risk communication for cardiometabolic diseases has focused on middle- or older-aged populations (1, 2). The assumption here is that older populations usually report higher levels of interest in seeking out disease risk information via genetic testing, given the saliency of health risks at this life stage. However, this dominant narrative does not explain why most interventions still fail to yield significant behavioural outcomes (12, 13, 19, 30, 31). Some evidence from survey studies indicate that the utility and relevance of genetic testing might actually be stronger in younger age groups (45, 46), suggesting that it may be inadequate to apply a single perspective to interpret the views of individuals at different life stages (Table 1). Additionally, clinically relevant life stages—such as pregnancy in women—are worth further consideration, as they represent key points for individuals to make important behavioural and lifestyle choices (47). Correspondingly, specific clinical populations such as women who have a history of gestational diabetes may hold different perceptions about T2D risk and/or react differently to genetic risk information, compared to the population most commonly under study (48, 49) (Table 1). The perceived value and potential of genetic testing may differ across these diverse groups—and it is even plausible to consider that communicating genetic risk information for health conditions to older adults later in the life course might be less productive than communicating similar information to younger people, a relationship which might explain the lack of observed effect in the literature. As such, it may be worth considering whether there are particular benefits in the application of genetic risk tools for specific age groups and/or clinical populations—especially in light of the proposed utility of genetic risk tools in being able to provide the earliest indication of risk in the first place (3, 8, 9). If genetic information can be conveyed at earlier life stages and in more effective ways—via strategies that target and promote healthy lifestyle practices from younger ages and/or in particular clinical subgroups—there may be greater opportunities to delay or prevent disease onset in high-risk individuals (51).

### Advancing the communication of genetic risk

#### Clarifying clinical outcomes from patients’ perspectives

Our synthesising argument suggests that future efforts aimed at modifying cognitive appraisals and/or behavioural responses via the provision of genetic/familial risk information will have to overlap at various levels—the individual, families, communities, as well as healthcare professionals and/or health systems—to begin tapping into various contextual factors that may play a role in individuals’ understandings and interpretations of risk. Even at the level of the individual, qualitative studies often suggest discrepancies between patient and clinical models of risk—specifically, that patients’ understanding of health conditions are often far more complicated than researchers expect (41, 52, 53). When contextualising familial risk, for example, both healthcare professionals and patients may rely on factors such as counting the number of affected relatives and understanding the age at which these relatives developed the condition (41, 53). However, patients also tended to have a more nuanced view—and the consequences of these discrepancies play out noticeably in a qualitative study directly comparing how family history of coronary heart disease is understood and communicated between patients and clinicians in primary care (53). Clinicians would attribute patients’ risk associated with family history to a “genetic element” that patients would not be able to change, treat it as a numerical adjustment to patients’ clinical risk scores and/or regard it as a “non-modifiable” factor—which can sometimes lead to the effect of family history being overstated (53). Patients, however, seemed keen to explore the multifactorial nature of risk—expressing interest in discussing this with clinicians, weighing up multiple risk factors and making comparisons between those that are “inherited” and what they think they are able to control or modify over time (53).

Such misalignments in understanding can create uncertainties that carry clinical and social implications over the course of a consultation—affecting clinicians’ ability to support patients in making informed decisions about their long-term management of disease risk (53). Importantly, researchers note that these uncertainties can partly be explained by a lack of knowledge around gene-environment interactions (53). Healthcare professionals in primary care hold limited knowledge about genetics—and their approaches are often contingent on existing guidelines for clinical practice (53).

As such, an additive model is often referenced, treating family history as a genetic, independent risk factor—or leaving it unexplained—whilst focusing on primary prevention approaches that prioritise immediate, modifiable risk factors. Yet the idea of risk is often more subjective for patients—who may benefit from more personalised approaches to risk assessment, as opposed to one that prioritises percentages and numbers. Whilst discussions over the multifactorial and non-deterministic nature of risk will no doubt come with their own complexities—the lack thereof often leads patients to express uncertainty over what can really be done in terms of improving their long-term outcomes, despite initial interest in reducing risk (53).

Thus, there can be benefits to equipping healthcare professionals with further knowledge and skill sets that can help clarify some of these uncertainties. Specifically, emphasising environmental and behavioural factors, alongside the possibility of prevention, may provide pathways for positive long-term health outcomes (52). Empirical studies support this notion as well. One study looked at the clinical utility of a composite risk score for atherosclerotic CVD by combining the effects of clinical risk factors and PRSs to estimate patients’ 10-year risk—and returning this information to participants via a web-based interactive tool in a clinical setting (7). This tool allowed participants to explore how altering certain modifiable risk factors within the system (e.g. changing smoking status, lowering cholesterol) can impact on their overall disease risk (7). At approximately 18 months, follow-up results indicated that 15.4% of the participants at high risk signed up for online health coaching; 20.8% consulted a doctor about their disease risk; 12.4% reported weight loss—and 14.2% of smokers reported quitting smoking (7). Objective measurements also showed that participants who reported weight loss and/or consulted doctors had significant reductions in systolic blood pressure and cholesterol (7). The researchers attributed these encouraging results largely to the interactive tool— which allowed the presentation of risk information in a personalised and comprehensive way, providing participants the opportunity to consider how their risk status might change depending on certain modifiable lifestyle and behavioural factors.

#### Using familial variables to leverage genetic risk information

The move beyond “selfhood” in this research area may also need to account for the impact that family systems can have in mediating individuals’ health-related beliefs. One avenue may be to combine family history assessments with genetic risk information—merging these two components to leverage the communication of disease risk. Lay understandings about the hereditary mechanisms of illnesses are often first based on family history (54). Thus, combining a personalised familial risk assessment approach with genetic risk information can help provide a baseline and social context to help individuals make sense of what is typically an objective figure, such as a genetic risk score (55, 56). Research has also suggested opportunities in using family history information to selectively identify patients who can benefit from genetic risk testing (or vice versa) (56). This is especially interesting to consider in light of recent work comparing the interplay of family history information and genome-wide PRSs across 24 common diseases (57). Family history and PRSs have independent and complementary effects in capturing individuals’ risk, highlighting the potential for more comprehensive ways to assess inherited disease risk (57). How these findings can be translated to risk communication in practice will be important to consider—including whether combining family history with genetic risk information in clinical settings can correspond to specific motivators for health behaviours.

Nevertheless, the challenge of bringing about sustained behaviour change remains. A range of multi-level influences are at play in familial contexts—including family food choices, household food insecurity and support for healthy lifestyle behaviours (58, 59). Some issues necessitate broader, more integrative approaches—but there is also potential for family units to present unique pathways to prevention and intervention. Familial systems and environments can be crucial to understand how individuals engage with health behaviours, as their structures and mechanisms allow for family members’ beliefs and behaviours to shape—and be shaped by—one another (60-62). For example, similar eating and/or lifestyle habits tend to be present in shared environments—alongside mutual understandings of any cultural meanings attached to such habits. This presents an environment in which collective practices and goals can be uniquely navigated—facilitating the definition of steps to meaningful health behaviour change.

Here, the unique cultural contexts of minoritised ethnic groups—some of which place distinctive emphasis on responsibilities towards families and/or communities—may present different opportunities and challenges (35, 36, 63-66). One study conducted in the Netherlands found that, compared to Dutch patients, Surinamese South Asian patients tended to report higher levels of concern over their families’ and relatives’ T2D risk (65). This is possibly related to a general sense of awareness about the prevalence of T2D in South Asian communities—but also, the concept of family often means different things for diverse populations. In South Asian communities, families may be large and transgenerational—inclusive of extended family members and even “unofficial” family members, such as close family friends (35, 36, 65). This can contrast to studies considering only family experiences in White European populations—and it is a difference that must be captured, in order to reflect the extent of impact that individuals from different backgrounds might perceive from family environments. The study also found that more Surinamese participants were motivated to convey T2D risk messages with their families—expressing willingness to educate family members about T2D risk and steps that can be taken for primary prevention (65). As such, there may be valuable opportunities here for interventions to try and tap into family systems as a whole—leveraging reciprocal influences within home environments as a resource to encourage the adoption of healthful behaviours that can be integrated into overall family lifestyles (60, 67).

#### Addressing the readiness of the health system

In the UK, clinical risk assessment and management procedures for cardiometabolic diseases occur largely in primary care—a process usually triggered by clinical findings that might indicate undiagnosed health conditions (9). The operational and logistical impact of incorporating genomic information into these settings will require careful assessment and planning across various services and resources in the NHS (8, 9). For example, existing clinical genetic laboratories are organised and coordinated in ways that mainly carry out testing for rare diseases. The position of PRS-based tests for cardiometabolic diseases—which might be used at-scale due to their higher prevalence—remains to be determined (8). At present, there are pilot trials exploring the integration of PRSs for CVD into NHS Health Checks—the national programme in offering free health checks every 5 years to adults between the ages of 40 to 74 (68, 69). A further idea has been proposed to bring forward the age at which patients can receive polygenic risk assessments, but this remains a highly debated issue (9). When considering the “appropriate” age at which individuals can undergo genetic assessments, some would argue for “the earlier the better”—since genetic risk can be quantified at birth and remain relatively stable over time (9). Evidence suggests benefits to starting as early as at 18 years, or even at pre-teen stages, to identify high-risk, pre-symptomatic young adults (8, 9). This will allow preventative action to be taken much earlier, instead of waiting until 40 for their first NHS Health Checks, during which clinical risk factors might already be established (9). PRSs can then be retained in patients’ electronic health records—used iteratively as an ongoing resource to inform future, longitudinal risk assessments (9). However, work is still needed to generate insight into how younger individuals might respond or react to genetic risk information—as well as how interventions aiming to target lifestyle and/or behaviour changes can be effectively implemented for these groups, possibly in coordination with other social and environmental resources.

Perhaps most importantly, healthcare professionals in primary care will require further resources and training to better interpret, communicate and answer questions about PRSs for different groups of patients. There are practical questions, such as how test results should be returned to clinicians—and in what format (e.g. as data and scores requiring further interpretation, or more detailed reports) (9). Ongoing conversations will be required to bring about co-design opportunities and determine clinician preferences (70, 71). Furthermore, whether and how the integration of genetic risk information will impact on clinical decision-making needs to be explored. A study exploring weight-related clinical interactions found that presenting genetic information about obesity to medical students resulted in lower health behaviour screening recommendations and referrals for patients in consultations (72). The possibility of genetic fatalism in clinicians may have unintended consequences—and further work is required to explore their perceptions and attitudes towards these proposed changes, as well as the shifts in responsibilities that they may be expected to take over and/or deliver.

As such, a range of developments—and crucially, funding—are required to ensure that the necessary expertise, resources and infrastructures are in place to support the integration of PRSs into existing health systems in this newly-proposed landscape. There is also a need for further research in several areas that have not been discussed in detail in this CIS. At present, PRSs have mostly been developed in populations of exclusively or majority White European ancestry. While some assessments have demonstrated that they are still able to discriminate between high and low risk groups in other ethnic populations, they do not perform equally well for all traits (5, 8, 9, 73). There is ongoing work aiming to address these limitations to diverse and representative data in GWASs—but time is needed to accumulate evidence. The integration of PRSs and conventional risk calculators also necessitates continuous updates to existing risk prediction models—ensuring that the additional genetic data is accurately embedded and taking into account any updated epidemiological information in diverse populations (9). Furthermore, to reiterate the need for a life course perspective, ongoing developments should aim to determine the age at which tools such as PRSs will likely add the most value. This may require novel research designs to account for the absence of conventional signs of disease in high-risk young patients. Additional endpoints, such as age of onset in premature incidence rates, may need to be considered (9).

## Discussion

### Principal findings

This CIS has discussed the complex evidence available on the communication of genetic/familial risk for cardiometabolic diseases. It includes a total of 188 records. Firstly, we explored how cognitive appraisals have been studied in relation to cardiometabolic disease risk in the literature—highlighting how the tendency to focus on “selfhood” can be a limiting perspective for the field. Delving deeper into these limitations, we argued that assumptions around the “ascetic subject of compliance” appear widely held in the research landscape—which may explain the apparent lack of convincing evidence surrounding current interventions that target individuals’ cognitive appraisals and health behaviours via the provision of genetic risk information. We generated a synthesising argument—beyond the “ascetic subject of compliance”—from a set of knowledge gaps that we have identified in the literature: (1) Difficulty applying existing theories/models to diverse populations; (2) The role of familial variables and (3) The need for a life course perspective. Finally, we discussed how these contextual factors and upstream determinants should be leveraged in future efforts to improve risk communication. Strategies will need to overlap at multiple levels—the individual, families, communities and health systems—in order to fully harness the effects of genetic risk tools and aid in efforts to mitigate cardiometabolic disease risk.

### Implications for research and clinical care

Some researchers have suggested the adoption of broader behavioural science frameworks, such as the Capability, Opportunity, Motivation, Behaviour (COM-B) model, to support the design of interventions communicating genetic/familial risk for cardiometabolic diseases (8). The COM-B model proposes that individuals will need the capability (physical and psychological), opportunity (social and physical) and motivation (reflective and automatic) to engage in risk-reducing behaviours, in the face of health threats. This takes a more comprehensive perspective, compared to most models discussed earlier in this CIS. Yet even so, interventions addressing solely these components may only be sufficient to motivate specific subsets of the population to engage in preventative behaviours under specific conditions—i.e. individuals who have the capabilities, opportunities and motivation to act on risk information (people with adequate financial, social and other resources) (8). The provision of risk information will still need to be combined with other forms of support to achieve the goals of motivating behaviour change more widely. At the macro-level, these may include system-level approaches to help address the social determinants of health—incorporating elements such as training, restructuring or environmental enablement to engage COM-B traits and facilitate constructive behaviour change at significant levels across the population (8). At the micro-level, exploring opportunities to leverage familial factors in risk assessment contexts may help support this avenue of research.

Ultimately, the translation of genetic risk prediction for cardiometabolic diseases from discovery research settings to clinical implementation still requires much work. Ongoing efforts to develop, test and validate the performance of genetic risk tools such as PRSs in diverse populations can help ensure that they are implementation-ready on a population-wide basis. There are also important considerations surrounding the logistical and infrastructural impact of integrating these tools into health systems—alongside potential challenges to build the necessary expertise and workforce to handle the anticipated influx of patients undergoing genetic risk assessments for common diseases. Importantly, establishing whether there may be additional benefit or value for patients from diverse backgrounds and/or age groups to receive genetic risk information—both from a risk assessment and a behavioural perspective—may be key to bring forward these efforts. There is also potential to target specific groups of clinical interest—e.g. women with a history of gestational diabetes, couples interested in family planning etc. However, further evidence on how these subsets of populations might respond or react to genetic risk information is still needed—as well as more work exemplifying how genetic risk information can be effectively presented in ways that can motivate preventative health behaviours, especially in those at high risk.

### Strengths and limitations

In applying a critical perspective to the literature, this CIS was able to build on the findings of conventional systematic reviews in this research area to generate further interpretations. The common strategy employed in conventional reviews—by clearly pre-specifying the study types and methods to be included—is a useful one for the methodical pooling and aggregation of data. However, this approach sometimes restricts the amount and/or type of relevant work that can be synthesised. Potentially valuable information from the surrounding literature may be lost, limiting the ability to offer a full critique of the research landscape (16). In taking a more comprehensive approach of a CIS, we were thus able to expand on previous findings illustrating the lack of convincing evidence surrounding interventions that communicate genetic risk—drawing from notions of auxiliary assumptions, the “ascetic subject of compliance” and a rich body of quantitative and qualitative work. This allowed us to consider beyond the empirical data, gain insight into specific gaps in the literature and ultimately, propose strategies that can be expanded upon in the field. There are, of course, also limitations to this method. Due to the breadth of the literature identified and included in this CIS, the process of developing a synthesising argument has been a subjective process, prone to various biases. However, we maintain that our analysis is demonstrably grounded in—and consistent with—the evidence base. Furthermore, our findings have been corroborated between different members of the multidisciplinary review team to allow for a range of perspectives to be addressed. We acknowledge that a different review team may generate different interpretations of the literature—and thus make no claims to reproducibility and generalisability—however, we believe this to be in line with the purpose of the CIS as a method, to facilitate the production of fresh insights in a research area.

## Conclusions

To our knowledge, this CIS is the first of its kind to be applied to research on the communication of genetic risk for cardiometabolic diseases. It integrates the rich quantitative and qualitative evidence available in the literature, bringing together insights from surrounding fields of behavioural and social sciences to generate a broader conceptualisation of current evidence and gaps. We identified a need for the literature to focus beyond individual-level cognitive appraisals that have been investigated in relation to genetic/familial risk communication. A critique was developed, building on the limitations of assuming an “ascetic subject of compliance”—a view we found to be predominantly held in this research landscape. This was followed by the generation of a synthesising argument—beyond the “ascetic subject of compliance”—constructed around three major gaps that we have observed from the literature: (1) Difficulty applying existing theories/models to diverse populations; (2) The role of familial variables and (3) The need for a life course perspective. We highlighted the importance of addressing various contextual factors and upstream determinants that can influence individuals’ responses at different levels, e.g. through interactions with their family systems, socio-cultural environments, as well as wider health provision. To begin addressing some of these gaps, efforts to improve the communication of genetic risk should consider clarifying clinical outcomes from patients’ perspectives, using familial variables to leverage genetic risk information—and crucially, address the readiness of the health system to accommodate these shifts.

## Data Availability

All data produced in the present work are contained in the manuscript.

## List of abbreviations

CENTRAL: the Cochrane Central Register of Controlled Trials
CIS: Critical interpretive synthesis
COM-B: the Capability, Opportunity, Motivation, Behaviour model
CSM-SR: the Common Sense Model of Self Regulation
CVD: Cardiovascular disease
GWASs: Genome-wide association studies
HBM: the Health Belief Model
PRSs: polygenic risk scores
T2D: Type 2 diabetes
TPB: the Theory of Planned Behaviour

## Declarations

### Ethics approval and consent to participate

Not applicable.

### Consent for publication

Not applicable.

### Availability of data and materials

All data generated in the current study are included in this published article and its additional files.

### Competing interests

The authors declare that they have no competing interests.

### Funding

This work was made possible by funding from the Wellcome Trust for JHL through the doctoral training programme Health Data in Practice: Human-centred Science (Reference: 218584/Z/19/Z).

### Authors’ contributions

JHL, SF and NF conceptualised the review topic. JHL conducted the initial search. All authors finalised the search terms and helped screen eligible records. JHL conducted data extraction and analysis. All authors contributed to the interpretation of data. JHL drafted the paper. SF, NS and NF reviewed and revised the drafts. All authors read and approved the final manuscript.

## Acknowledgements

The authors would like to thank Paula Funnel, the Faculty Liaison Librarian for Medicine & Dentistry at Queen Mary University of London, for assisting with the definition and validation of search terms across various databases.

## Additional files

Additional file 1. Search strategy (Ovid MEDLINE).

Additional file 2. Purposive sampling and quality appraisal process for databases searches.

Additional file 3. Example of data extraction process on NVivo.

Additional file 4. List of records included in this CIS.

## Notes

### Competing Interest Statement

The authors have declared no competing interest.

