## Additional file 1 for "Advancing the communication of genetic risk for cardiometabolic diseases: A critical interpretive synthesis"

Database:

Books@Ovid <November 01, 2021>, Journals@Ovid Full Text <November 02, 2021>, Your Journals@Ovid, Ovid MEDLINE(R) and Epub Ahead of Print, In-Process, In-Data-Review & Other Non-Indexed Citations, Daily and Versions(R) <1946 to November 02, 2021>

Search Strategy:

1. genetic testing/ (41441)
2. genetic services/ (514)
3. genetic counseling/ (14964)
4. genetic predisposition to disease/ (148142)
5. polygenic risk score$.mp. (4767)
6. polygenic risk.mp. (5605)
7. 1 or 2 or 3 or 4 or 5 or 6 (194419)
8. ((gene or genes or genetic* or genotype* or DNA or famil*) adj3 (test* or assess* or risk* or susceptib* or predispos* or disease* or screen* or prognos* or predict* or servic*)).ti,ab,kw. (572368)
9. counseling/ or directive counseling/ (40198)
10. health communication/ (2928)
11. (consult* or assess* or support* or inform* or advis* or advice or counsel* or educat* or shar* or communicat* or teach* or discuss* or decid* or decision*).ti,ab,kw. (11072860)
12. patient education as topic/ (87569)
13. 9 or 10 or 11 or 12 (11110477)
14. 8 and 13 (257334)
15. 7 or 14 (417698)
16. exp obesity/ (234408)
17. exp type 2 diabetes/ (148414)
18. exp cardiovascular disease/ (2546995)
19. 16 or 17 or 18 (2839871)
20. 15 and 19 (40882)
21. ((perceive$ or percep$ or apprai$) adj3 (illness or risk$ or susceptibility or control$)).mp. (156746)
22. control belief$.mp. (3902)
23. controllab$.mp. (55900)
24. 21 or 22 or 23 (209373)
25. 20 and 24 (129)
