## Additional file 2 for "Advancing the communication of genetic risk for cardiometabolic diseases: A critical interpretive synthesis"

| REFERENCE | SOURCE | Maximum variation | Data richness | Match of scope | Decision | Additional notes |
| --- | --- | --- | --- | --- | --- | --- |
| 2010. Abstracts of Diabetes UK Annual Professional Conference. Diabetic Medicine, 27. | EMBASE | Yes | No | Yes | Exclude | See A7 abstract |
| ACHESON, L. S., WANG, C., ZYZANSKI, S. J., LYNN, A., RUFFIN, M. T. T., GRAMLING, R., RUBINSTEIN, W. S., O'NEILL, S. M., NEASE, D. E., JR. & FAMILY HEALTHWARE IMPACT TRIAL GROUP 2010. Family history and perceptions about risk and prevention for chronic diseases in primary care: a report from the family healthware impact trial. Genetics in medicine : official journal of the American College of Medical Genetics, 12, 212-8. | Ovid MEDLINE | No | No | Yes | Include |  |
| AL SHAFAEE, M. A., AL-SHUKAILI, S., RIZVI, S. G. A., AL FARSI, Y., KHAN, M. A., GANGULY, S. S., AFIFI, M. & AL ADAWI, S. 2008. Knowledge and perceptions of diabetes in a semi-urban Omani population. BMC PUBLIC HEALTH, 8. | Web of Science | Yes | No | Yes | Include |  |
| ALBERT, M. A., RAVENELL, J., GLYNN, R. J., KHERA, A., HALEVY, N. & DE LEMOS, J. A. 2008. Cardiovascular risk indicators and perceived race/ethnic discrimination in the Dallas Heart Study. American heart journal, 156, 1103-9. | Ovid MEDLINE | Yes | No | Yes | Exclude | Interesting study but some limitations in study design to address its relevant research questions – focuses on association between race/ethnic discrimination and subclinical cardiovascular disease, but is a cross-sectional analysis |
| ALKERWI, A., PAGNY, S., LAIR, M. L., DELAGARDELLE, C. & BEISSEL, J. 2013. Level of Unawareness and Management of Diabetes, Hypertension, and Dyslipidemia among Adults in Luxembourg: Findings from ORISCAV-LUX Study. PLOS ONE, 8. | Web of Science | No | No | Yes | Include |  |
| ALONSO, R., PEREZ DE ISLA, L., MUNIZ-GRIJALVO, O. & MATA, P. 2020. Barriers to Early Diagnosis and Treatment of Familial Hypercholesterolemia: Current Perspectives on Improving Patient Care. Vascular health and risk management, 16, 11-25. | Ovid MEDLINE | Yes | Yes | No | Exclude | Focuses on barriers to diagnosis for familial hypercholesterolaemia |
| AMIREAULT, S., GODIN, G., VOHL, M. C. & PERUSSE, L. 2008. Moderators of the intention-behaviour and perceived behavioural control-behaviour relationships for leisure-time physical activity. INTERNATIONAL JOURNAL OF BEHAVIORAL NUTRITION AND PHYSICAL ACTIVITY, 5. | Web of Science | Yes | No | Yes | Include |  |
| AMUTA, A. O. 2016. Diabetes family health history among college students. 77, ProQuest Information & Learning. | PsycINFO | Yes | Yes | Yes | Include |  |
| ANDERSON, A. S., CASWELL, S., MACLEOD, M., STEELE, R. J. C., BERG, J., DUNLOP, J., STEAD, M., EADIE, D. & O'CARROLL, R. E. 2017. Health behaviors and their relationship with disease control in people attending genetic clinics with a family history of breast or colorectal cancer. Journal of Genetic Counseling, 26, 40-51. | PsycINFO | No | Yes | No | Exclude |  |
| ARESTEDT, K., AGREN, S., FLEMME, I., MOSER, D. & STROMBERG, A. 2010. Psychometric properties of the Swedish version of the Control Attitudes Scale for patients with cardiac disease and their family members. European Journal of Cardiovascular Nursing, 9, S26-S27. | EMBASE | No | No | Yes | Exclude | Focuses on instrument development |
| ARTMANN, A., GOTTSCHALK, N., JACOBS, V. R., MEINDL, A., KIECHLE, M., RUMMENY, E. J. & HARBECK, N. 2009. Counseling and care: A Breast Cancer Awareness Training to improve participation and adherence to screening recommendations in women with a family history of breast cancer. Journal of Clinical Oncology, 27, 1530. | EMBASE | Yes | No | No | Exclude | Focuses on breast cancer |
| ASRIL, N. M., TABUCHI, K., TSUNEMATSU, M., KOBAYASHI, T. & KAKEHASHI, M. 2020. Predicting Healthy Lifestyle Behaviours Among Patients With Type 2 Diabetes in Rural Bali, Indonesia. Clinical Medicine Insights: Endocrinology and Diabetes, 13. | EMBASE | Yes | No | Yes | Include |  |
| BAIG, K., ZAIDI, T. H., MEHTAB, K., FARID, M., KHALIQ, S., MUKHTAR, W., TARIQ, S. & ZAIDI, F. 2020. Knowledge, Attitude and Practices of Type 2 Diabetic patients attending a tertiary care hospital in Karachi. WORLD FAMILY MEDICINE, 18, 20-28. | Web of Science | Yes | No | No | Exclude | Some limitations in study design – unclear what materials and measures were used/how they were developed? |
| BANERJEE, A. T., MAHAJAN, A., MATHUR-BALENDRA, A., QURESHI, N., TEEKAH, M., YOGARATNAM, S., PRABHAKAR, P., AHMED, S., SHAH, B. R., VELUMMAILUM, R., PRICE, J. A. D., DE SOUZA, R. J. & BAJAJ, H. S. 2022. Impact of the South Asian Adolescent Diabetes Awareness Program (SAADAP) on diabetes knowledge, risk perception and health behaviour. HEALTH EDUCATION JOURNAL. | Web of Science | Yes | No | Yes | Include |  |
| BASILIO, C. D., KWAN, V. S. Y. & TOWERS, M. J. 2016. Culture and Risk Assessments: Why Latino Americans Perceive Greater Risk for Diabetes. CULTURAL DIVERSITY & ETHNIC MINORITY PSYCHOLOGY, 22, 104-113. | Web of Science | Yes | No | Yes | Include |  |
| BATES, B. R., TEMPLETON, A., ACHTER, P. J., HARRIS, T. M. & CONDIT, C. M. 2003. What does "a gene for heart disease" mean? A focus group study of public understandings of genetic risk factors. American journal of medical genetics. Part A, 119A, 156-61. | Ovid MEDLINE | Yes | Yes | Yes | Include |  |
| BATTE, B., SHELDON, J. P., ARSCOTT, P., HUISMANN, D. J., SALBERG, L., DAY, S. M. & YASHAR, B. M. 2015. Family communication in a population at risk for Hypertrophic Cardiomyopathy. Journal of Genetic Counseling, 24, 336-348. | PsycINFO | No | No | No | Exclude |  |
| BEKKE-HANSEN, S., WEINMAN, J., THASTUM, M., THYGESEN, K. & ZACHARIAE, R. 2014. Psycho-social factors are important for the perception of disease in patients with acute coronary disease. Danish medical journal, 61, A4885. | Ovid MEDLINE | No | No | Yes | Include |  |
| BENNICH, B. B., MUNCH, L., OVERGAARD, D., KONRADSEN, H., KNOP, F. K., RODER, M., VILSBOLL, T. & EGEROD, I. 2020. Experience of family function, family involvement, and self-management in adult patients with type 2 diabetes: A thematic analysis. JOURNAL OF ADVANCED NURSING, 76, 621-631. | Web of Science | No | Yes | Yes | Include |  |
| BERGE, J. M., ARIKIAN, A., DOHERTY, W. J. & NEUMARK-SZTAINER, D. 2012. Healthful eating and physical activity in the home environment: Results from multifamily focus groups. Journal of Nutrition Education and Behavior, 44, 123-131. | PsycINFO | Yes | Yes | Yes | Include |  |
| BERNHARDT, B. A., TAMBOR, E. S., FRASER, G., WISSOW, L. S. & GELLER, G. 2003. Parents' and children's attitudes toward the enrollment of minors in genetic susceptibility research: implications for informed consent. American journal of medical genetics. Part A, 116A, 315-23. | Ovid MEDLINE | Yes | Yes | Yes | Include | Focuses on ethics of participation in genetics research but could give useful insights into perceptions about genetic testing for prevention |
| BIN ZAHID, H., NAWAZ, A. & ALI, H. 2019. KNOWLEDGE ASSESSMENT OF DIABETES AND INCIDENCE OF HYPERTENSION IN TYPE 2 DIABETIC PATIENTS. INDO AMERICAN JOURNAL OF PHARMACEUTICAL SCIENCES, 6, 4644-4651. | Web of Science | Yes | No | No | Exclude | Some limitations in study design – unclear justification for dividing participants based on family history, especially given unequal comparisons? |
| BITTENCOURT, M., LAURINAVICIUS, A., PEREIRA, C., CESENA, F., CONCEIÇAO, R. & SANTOS, R. D. 2018. Underdiagnosis, undertreatment and cardiovascular risk misperception among individuals with suspected familial hypercholesterolemia: A brazilian survey. Atherosclerosis Supplements, 32, 48. | EMBASE | Yes | No | Yes | Exclude | Focuses on underdiagnosis of familial hypercholesterolaemia |
| BLUE, G. M., KASPARIAN, N. A., SHOLLER, G. F., KIRK, E. P. & WINLAW, D. S. 2015. Genetic counselling in parents of children with congenital heart disease significantly improves knowledge about causation and enhances psychosocial functioning. International journal of cardiology, 178, 124-30. | Ovid MEDLINE | Yes | No | Yes | Include |  |
| BOELDT, D. L., SCHORK, N. J., TOPOL, E. J. & BLOSS, C. S. 2015. Influence of individual differences in disease perception on consumer response to direct-to-consumer genomic testing. Clinical Genetics, 87, 225-232. | EMBASE | No | No | Yes | Include |  |
| BONNER, C., SPINKS, C., SEMSARIAN, C., BARRATT, A., INGLES, J. & MCCAFFERY, K. 2018. Psychosocial Impact of a Positive Gene Result for Asymptomatic Relatives at Risk of Hypertrophic Cardiomyopathy. Journal of genetic counseling, 27, 1040-1048. | Ovid MEDLINE | Yes | Yes | No | Exclude | Focuses on “silent gene carriers” of cardiomyopathy |
| BOSSUYT, P. M. M., RAAYMAKERS, T. W. M., BONSEL, G. J. & RINKEL, G. J. E. 2005. Screening families for intracranial aneurysms: anxiety, perceived risk, and informed choice. Preventive medicine, 41, 795-9. | Ovid MEDLINE | No | No | No | Exclude | Focuses on intracranial aneurysm and the effects of invitations to family screening |
| BOWLES, B. C. 2010. Genograms as threat appeals: Using the extended parallel process model with familial cardiovascular disease. 70, ProQuest Information & Learning. | PsycINFO | No | No | Yes | Include |  |
| BROCK, J. F. & GORDON, H. 1957. The dietetics of coronary heart disease. South African medical journal, 31, 663-671. | EMBASE | Yes | Yes | No | Exclude |  |
| BROEKHUIZEN, K., VAN POPPEL, M. N. M., KOPPES, L. L. J., BRUG, J. & VAN MECHELEN, W. 2010. A tailored lifestyle intervention to reduce the cardiovascular disease risk of individuals with Familial Hypercholesterolemia (FH): design of the PRO-FIT randomised controlled trial. BMC PUBLIC HEALTH, 10. | Web of Science | No | No | Yes | Exclude | Protocol – no outcomes |
| BROWN, J. B., HARRIS, S. B., WEBSTER-BOGAERT, S., WETMORE, S., FAULDS, C. & STEWART, M. 2002. The role of patient, physician and systemic factors in the management of type 2 diabetes mellitus. FAMILY PRACTICE, 19, 344-349. | Web of Science | No | Yes | Yes | Include |  |
| BROWN, M. C., BELL, R., COLLINS, C., WARING, G., ROBSON, S. C., WAUGH, J. & FINCH, T. 2013. Women's Perception of Future Risk Following Pregnancies Complicated by Preeclampsia. HYPERTENSION IN PREGNANCY, 32, 60-73. | Web of Science | No | No | No | Exclude |  |
| BRUST-RENCK, P. G., REYNA, V. F., WILHELMS, E. A. & LAZAR, A. N. 2016. A fuzzy-trace theory of judgment and decision-making in health care: Explanation, prediction, and application. In: DIEFENBACH, M. A., MILLER-HALEGOUA, S. & BOWEN, D. J. (eds.) Handbook of health decision science. New York, NY: Springer Science + Business Media. | PsycINFO | No | Yes | Yes | Include | Book chapter |
| BUIGUES, C., QUERALT, A., DE VELASCO, J. A., SALVADOR-SANZ, A., JENNINGS, C., WOOD, D. & TRAPERO, I. 2021. Psycho-Social Factors in Patients with Cardiovascular Disease Attending a Family-Centred Prevention and Rehabilitation Programme: EUROACTION Model in Spain. LIFE-BASEL, 11. | Web of Science | No | No | Yes | Include |  |
| BURNS, L., KENNY, U., HEALY, L., CUSHEN, S., O'REILLY, S., RYAN, A. M. & POWER, D. G. 2012. Public perception of cancer risk. Journal of Clinical Oncology, 30. | EMBASE | No | No | No | Exclude |  |
| CAMERON, L. D., SHERMAN, K. A., MARTEAU, T. M. & BROWN, P. M. 2009. Impact of genetic risk information and type of disease on perceived risk, anticipated affect, and expected consequences of genetic tests. Health psychology : official journal of the Division of Health Psychology, American Psychological Association, 28, 307-16. | Ovid MEDLINE | Yes | No | Yes | Include |  |
| CERSOSIMO, E. & MUSI, N. 2011. Improving Treatment in Hispanic/Latino Patients. AMERICAN JOURNAL OF MEDICINE, 124, S16-S21. | Web of Science | Yes | Yes | Yes | Include |  |
| CHARBONNEAU, J., NICOL, D., CHALMERS, D., KATO, K., YAMAMOTO, N., WALSHE, J. & CRITCHLEY, C. 2020. Public reactions to direct-to-consumer genetic health tests: A comparison across the US, UK, Japan and Australia. European Journal of Human Genetics, 28, 339-348. | EMBASE | Yes | No | Yes | Include |  |
| CHEILOUDAKI, E. & ALEXOPOULOS, E. C. 2019. Adherence to treatment in stroke patients. International Journal of Environmental Research and Public Health, 16. | EMBASE | No | No | No | Exclude |  |
| CHIA, J. M. X., GOH, Z. S., SEOW, P. S., SEOW, T. Y. Y., CHOO, J. C. J., FOO, M. W. Y., NEWMAN, S. & GRIVA, K. 2021. Psychosocial Factors, Intentions to Pursue Arteriovenous Dialysis Access, and Access Outcomes: A Cohort Study. American Journal of Kidney Diseases, 77, 931-940. | EMBASE | Yes | No | No | Exclude |  |
| CHO, A. H., KILLEYA-JONES, L. A., O'DANIEL, J. M., KAWAMOTO, K., GALLAGHER, P., HAGA, S., LUCAS, J. E., TRUJILLO, G. M., JOY, S. V. & GINSBURG, G. S. 2012. Effect of genetic testing for risk of type 2 diabetes mellitus on health behaviors and outcomes: study rationale, development and design. BMC health services research, 12, 16. | Ovid MEDLINE | No | No | Yes | Exclude | Protocol – no outcomes |
| CHRISTIAANS, I., VAN LANGEN, I. M., BIRNIE, E., BONSEL, G. J., WILDE, A. A. M. & SMETS, E. M. A. 2009. Quality of life and psychological distress in hypertrophic cardiomyopathy mutation carriers: a cross-sectional cohort study. American journal of medical genetics. Part A, 149A, 602-12. | Ovid MEDLINE | No | No | No | Exclude | Focuses on cardiomyopathy |
| CISLAK, A., SAFRON, M., PRATT, M., GASPAR, T. & LUSZCZYNSKA, A. 2012. Family-related predictors of body weight and weight-related behaviours among children and adolescents: a systematic umbrella review. CHILD CARE HEALTH AND DEVELOPMENT, 38, 321-331. | Web of Science | Yes | Yes | Yes | Include |  |
| CITARELLA, A., KIELER, H., SUNDSTROM, A., LINDER, M., WETTERMARK, B., BERGLIND, I. A. & ANDERSEN, M. 2014. Family history of cardiovascular disease and influence on statin therapy persistence. EUROPEAN JOURNAL OF CLINICAL PHARMACOLOGY, 70, 701-707. | Web of Science | Yes | No | Yes | Include |  |
| CLAASSEN, L., HENNEMAN, L., DE VET, R., KNOL, D., MARTEAU, T. & TIMMERMANS, D. 2010. Fatalistic responses to different types of genetic risk information: Exploring the role of self-malleability. Psychology & Health, 25, 183-196. | PsycINFO | Yes | No | Yes | Include |  |
| CLAASSEN, L., HENNEMAN, L., KINDT, I., MARTEAU, T. M. & TIMMERMANS, D. R. M. 2010. Perceived risk and representations of cardiovascular disease and preventive behaviour in people diagnosed with familial hypercholesterolemia: a cross-sectional questionnaire study. Journal of health psychology, 15, 33-43. | Ovid MEDLINE | No | No | Yes | Include |  |
| CLAASSEN, L., HENNEMAN, L., VAN DER WEIJDEN, T., MARTEAU, T. M. & TIMMERMANS, D. R. M. 2012. Being at risk for cardiovascular disease: perceptions and preventive behavior in people with and without a known genetic predisposition. Psychology, health & medicine, 17, 511-21. | Ovid MEDLINE | No | No | Yes | Include |  |
| COHEN, J. S. & BIESECKER, B. B. 2010. Quality of life in rare genetic conditions: A systematic review of the literature. American Journal of Medical Genetics, Part A, 152, 1136-1156. | EMBASE | Yes | Yes | No | Exclude |  |
| COLLINS, J., RYAN, L. & TRUBY, H. 2014. A systematic review of the factors associated with interest in predictive genetic testing for obesity, type II diabetes and heart disease. Journal of human nutrition and dietetics : the official journal of the British Dietetic Association, 27, 479-88. | Ovid MEDLINE | Yes | Yes | Yes | Include |  |
| CULLEN, K. W. & BUZEK, B. B. 2009. Knowledge about type 2 diabetes risk and prevention of African-American and Hispanic adults and adolescents with family history of type 2 diabetes. The Diabetes educator, 35, 836-42. | Ovid MEDLINE | Yes | Yes | Yes | Include |  |
| CUNNINGHAM, A. T., GENTSCH, A. T., DOTY, A. M. B., MILLS, G., LANOUE, M., CARR, B. G., HOLLANDER, J. E. & RISING, K. L. 2020. "I had no other choice but to catch it too": the roles of family history and experiences with diabetes in illness representations. BMC endocrine disorders, 20, 95. | Ovid MEDLINE | Yes | Yes | Yes | Include |  |
| DAACK-HIRSCH, S., SCHUMACHER, A. C., SHAH, L. & CAMPO, S. 2019. Type 2 diabetes familial risk personalization process profiles: Implications for patient-provider communication. Research in nursing & health, 42, 369-381. | Ovid MEDLINE | Yes | Yes | Yes | Include |  |
| DAACK-HIRSCH, S., SHAH, L. L. & CADY, A. D. 2018. Mental Models of Cause and Inheritance for Type 2 Diabetes Among Unaffected Individuals Who Have a Positive Family History. Qualitative health research, 28, 534-547. | Ovid MEDLINE | Yes | Yes | Yes | Include |  |
| DAACK-HIRSCH, S., SHAH, L. L., JONES, K., ROCHA, B., DOERR, M., GABITZSCH, E. & MEESE, T. 2020. All things considered, my risk for diabetes is medium: A risk personalization process of familial risk for type 2 diabetes. Health expectations : an international journal of public participation in health care and health policy, 23, 169-181. | Ovid MEDLINE | Yes | Yes | Yes | Include |  |
| DAMMAN, O. C., BOGAERTS, N. M. M., VAN DEN HAAK, M. J. & TIMMERMANS, D. R. M. 2017. How lay people understand and make sense of personalized disease risk information. Health Expectations: An International Journal of Public Participation in Health Care & Health Policy, 20, 973-983. | PsycINFO | No | Yes | Yes | Include |  |
| DAR-NIMROD, I., CHEUNG, B. Y., RUBY, M. B. & HEINE, S. J. 2014. Can merely learning about obesity genes affect eating behavior? Appetite, 81, 269-76. | Ovid MEDLINE | No | No | Yes | Include |  |
| DAVIES, L. E. & THIRLAWAY, K. 2013. The influence of genetic explanations of type 2 diabetes on patients' attitudes to prevention, treatment and personal responsibility for health. Public health genomics, 16, 199-207. | Ovid MEDLINE | No | No | Yes | Include |  |
| DE GROOT, M. & WESSEL, J. 2014. Genetic Testing and Type 2 Diabetes Risk Awareness. The Diabetes educator, 40, 427-433. | Ovid MEDLINE | No | No | Yes | Include |  |
| DESALVO, K. B., GREGG, J., KLEINPETER, M., PEDERSEN, B. R., STEPTER, A. & PEABODY, J. 2005. Cardiac risk underestimation in urban, black women. JOURNAL OF GENERAL INTERNAL MEDICINE, 20, 1127-1131. | Web of Science | Yes | No | Yes | Include |  |
| DHALIWAL, H. S., SINGH, R., ABRAHAM, A. M., SHARMA, R., GOYAL, N. K., SOLOMAN, R., BANSAL, P. & GOYAL, A. Perception of Illness and Its Association with Treatment Willingness in Patients with Newly Diagnosed Nonalcoholic Fatty Liver Disease. DIGESTIVE DISEASES AND SCIENCES. | Web of Science | No | No | No | Exclude |  |
| DIGNAN, M. B., YOUNG, L. D., CROUSE, J. R. & KING, J. M. 1995. Factors associated with participation in a preventive cardiology service by patients with coronary heart disease. Southern medical journal, 88, 1057-61. | Ovid MEDLINE | No | No | Yes | Include |  |
| DILORENZO, T. A., SCHNUR, J., MONTGOMERY, G. H., ERBLICH, J., WINKEL, G. & BOVBJERG, D. H. 2006. A model of disease-specific worry in heritable disease: the influence of family history, perceived risk and worry about other illnesses. Journal of behavioral medicine, 29, 37-49. | Ovid MEDLINE | Yes | No | Yes | Include |  |
| DONG, Y. & BRANSCUM, P. 2019. What Motivates Individuals to Get Obesity Related Direct-To-Consumer Genetic Tests? A Reasoned Action Approach. AMERICAN JOURNAL OF HEALTH EDUCATION, 50, 356-365. | Web of Science | Yes | No | Yes | Include |  |
| DORMAN, J. S., VALDEZ, R., LIU, T., WANG, C., RUBINSTEIN, W. S., O'NEILL, S. M., ACHESON, L. S., RUFFIN, M. T. T. & KHOURY, M. J. 2012. Health beliefs among individuals at increased familial risk for type 2 diabetes: implications for prevention. Diabetes research and clinical practice, 96, 156-62. | Ovid MEDLINE | No | No | Yes | Include |  |
| DRAPKIN, R. G., WING, R. R. & SHIFFMAN, S. 1995. Responses to hypothetical high risk situations: Do they predict weight loss in a behavioral treatment program or the context of dietary lapses? Health Psychology, 14, 427-434. | PsycINFO | No | Yes | Yes | Include |  |
| EDELSTEIN, J. & LINN, M. W. 1985. The influence of the family on control of diabetes. Social Science & Medicine, 21, 541-544. | PsycINFO | No | No | Yes | Include |  |
| ERBLICH, J., BOVBJERG, D. H., NORMAN, C., VALDIMARSDOTTIR, H. B. & MONTGOMERY, G. H. 2000. It won't happen to me: Lower perception of heart disease risk among women with family histories of breast cancer. PREVENTIVE MEDICINE, 31, 714-721. | Web of Science | Yes | No | Yes | Include |  |
| ETCHEGARY, H., ENRIGHT, G., AUDAS, R., PULLMAN, D., YOUNG, T.-L. & HODGKINSON, K. 2016. Perceived economic burden associated with an inherited cardiac condition: a qualitative inquiry with families affected by arrhythmogenic right ventricular cardiomyopathy. Genetics in medicine : official journal of the American College of Medical Genetics, 18, 584-92. | Ovid MEDLINE | No | Yes | No | Exclude | Focuses on economic burden associated with cardiomyopathy |
| ETELSON, D., BRAND, D. A., PATRICK, P. A. & SHIRALI, A. 2003. Childhood obesity: Do parents recognize this health risk? OBESITY RESEARCH, 11, 1362-1368. | Web of Science | No | No | Yes | Include |  |
| FALCONER, C., SKOW, A., BLACK, J., SOVIO, U., SAXENA, S., CROKER, H., KESSEL, A., VINER, R. & KINRA, S. 2012. Does BMI FEEDBACK change parental perceptions about The health risk associated with their child's BMI? Obesity Facts, 5, 240. | EMBASE | Yes | No | No | Exclude |  |
| FALLAIZE, R., MACREADY, A. L., BUTLER, L. T., ELLIS, J. A. & LOVEGROVE, J. A. 2013. An insight into the public acceptance of nutrigenomic-based personalised nutrition. NUTRITION RESEARCH REVIEWS, 26, 39-48. | Web of Science | Yes | Yes | No | Exclude |  |
| FARMER, A. J., LEVY, J. C. & TURNER, R. C. 1999. Knowledge of risk of developing diabetes mellitus among siblings of Type 2 diabetic patients. Diabetic medicine, 16, 233‐237. | Cochrane Central Register of Controlled Trials (CENTRAL) | No | No | Yes | Include |  |
| FERRANTI, E. P. 2014. Dietary quality and cardiometabolic risk after gestational diabetes. 74, ProQuest Information & Learning. | PsycINFO | Yes | No | Yes | Include |  |
| FISHER, E., ACHILLES, S. & TÖNNIES, H. 2014. Predictive genetic testing, risk communication, and risk perception: An international expert meeting in Berlin, Germany. Journal of Community Genetics, 5, 1-5. | EMBASE | Yes | Yes | Yes | Include |  |
| FLAHERTY, G., GIBSON, I., JONES, J., CONNOLLY, S. & CROWLEY, J. 2011. Durability of lifestyle change and cardiovascular disease (CVD) risk factor reductions-1 year outcomes from a community based CVD prevention programme for high risk patients in Ireland. European Heart Journal, 32, 225. | EMBASE | No | No | Yes | Exclude | Study not complete – no outcomes |
| FOLLING, I. S., SOLBJOR, M., MIDTHJELL, K., KULSENG, B. & HELVIK, A.-S. 2016. Exploring lifestyle and risk in preventing type 2 diabetes-a nested qualitative study of older participants in a lifestyle intervention program (VEND-RISK). BMC public health, 16, 876. | Ovid MEDLINE | No | Yes | Yes | Include |  |
| FRICH, J. C., OSE, L., MALTERUD, K. & FUGELLI, P. 2006. Perceived Vulnerability to Heart Disease in Patients with Familial Hypercholesterolemia: A Qualitative Interview Study. Annals of Family Medicine, 4, 198-204. | PsycINFO | Yes | Yes | Yes | Include |  |
| FRIEDRICH, O., KUNSCHITZ, E., PONGRATZ, L., WIELÄNDER, S., SCHÖPPL, C. & SIPÖTZ, J. 2021. Classification of illness attributions in patients with coronary artery disease. Psychology & health, 1-16. | EMBASE | No | No | Yes | Include |  |
| FRIJLING, B. D., LOBO, C. M., KEUS, I. M., JENKS, K. M., AKKERMANS, R. P., HULSCHER, M. E. J. L., PRINS, A., VAN DER WOUDEN, J. C. & GROL, R. P. T. M. 2004. Perceptions of cardiovascular risk among patients with hypertension or diabetes. Patient education and counseling, 52, 47-53. | Ovid MEDLINE | No | No | Yes | Include |  |
| FROSCH, D. L., MELLO, P. & LERMAN, C. 2005. Behavioral consequences of testing for obesity risk. Cancer epidemiology, biomarkers & prevention : a publication of the American Association for Cancer Research, cosponsored by the American Society of Preventive Oncology, 14, 1485-9. | Ovid MEDLINE | No | No | Yes | Include |  |
| GALLAGHER, P., KING, H. A., HAGA, S. B., ORLANDO, L. A., JOY, S. V., TRUJILLO, G. M., SCOTT, W. M., BEMBE, M., CREIGHTON, D. L., CHO, A. H., GINSBURG, G. S. & VORDERSTRASSE, A. 2015. Patient beliefs and behaviors about genomic risk for type 2 diabetes: Implications for prevention. Journal of Health Communication, 20, 728-735. | PsycINFO | No | No | Yes | Include |  |
| GIBSON, E. G., GAGE, J. C., CASTLE, P. E. & SCARINCI, I. C. 2019. Perceived Susceptibility to Cervical Cancer among African American Women in the Mississippi Delta: Does Adherence to Screening Matter? WOMENS HEALTH ISSUES, 29, 38-47. | Web of Science | Yes | No | No | Exclude |  |
| GODIN, G., BELANGER-GRAVEL, A., AMIREAULT, S., VOHL, M.-C. & PERUSSE, L. 2011. The effect of mere-measurement of cognitions on physical activity behavior: a randomized controlled trial among overweight and obese individuals. The international journal of behavioral nutrition and physical activity, 8, 2. | Ovid MEDLINE | No | No | No | Exclude | Some limitations in study design – testing mere-measurement effects of questionnaires but unclear why control group was also given a questionnaire (dietary instead of physical activity – but what is the comparison?); also not following up on objective behavioural outcomes |
| GODINO, J. G., VAN SLUIJS, E. M. F., MARTEAU, T. M., SUTTON, S., SHARP, S. J. & GRIFFIN, S. J. 2012. Effect of communicating genetic and phenotypic risk for type 2 diabetes in combination with lifestyle advice on objectively measured physical activity: protocol of a randomised controlled trial. BMC PUBLIC HEALTH, 12. | Web of Science | No | No | Yes | Exclude | Protocol – no outcomes |
| GODINO, J. G., VAN SLUIJS, E. M. F., MARTEAU, T. M., SUTTON, S., SHARP, S. J. & GRIFFIN, S. J. 2016. Lifestyle Advice Combined with Personalized Estimates of Genetic or Phenotypic Risk of Type 2 Diabetes, and Objectively Measured Physical Activity: A Randomized Controlled Trial. PLoS medicine, 13, e1002185. | Ovid MEDLINE | No | No | Yes | Include |  |
| GOODWIN, C. L. 2018. A randomized controlled trial of heart disease risk education on delay discounting, perceived disease risk, health behavior, and health behavior intentions among men and women with and without a family history of cardiovascular disease. 79, ProQuest Information & Learning. | PsycINFO | Yes | No | Yes | Include |  |
| GRABOWSKI, D. & ANDERSEN, T. H. 2020. Barriers to intra-familial prevention of type 2 diabetes: A qualitative study on horizons of significance and social imaginaries. Chronic Illness, 16, 119-130. | EMBASE | Yes | Yes | Yes | Include |  |
| GRAUMAN, Å., VELDWIJK, J., JAMES, S., HANSSON, M. & BYBERG, L. 2021. Good general health and lack of family history influence the underestimation of cardiovascular risk: a cross-sectional study. European journal of cardiovascular nursing : journal of the Working Group on Cardiovascular Nursing of the European Society of Cardiology. | EMBASE | No | No | Yes | Include |  |
| GRAY, W. N., JANICKE, D. M., WISTEDT, K. M. & DUMONT-DRISCOLL, M. C. 2010. Factors associated with parental use of restrictive feeding practices to control their children's food intake. APPETITE, 55, 332-337. | Web of Science | Yes | No | No | Exclude |  |
| GREENLEE, H., CREW, K., FERGUSON, K., MCKINLEY, P., RUNDLE, A., OGEDEGBE, G., MATTA, J., SANDOVAL, R. & HERSHMAN, D. L. 2009. Facilitators and barriers to recruitment for an exercise and dietary intervention study in minority breast cancer survivors. Cancer Research, 69. | EMBASE | Yes | No | No | Exclude |  |
| GROSS, C. P., FILARDO, G., SINGH, H. S., FREEDMAN, A. N. & FARRELL, M. H. 2006. The relation between projected breast cancer risk, perceived cancer risk, and mammography use: Results from the national health interview survey. Journal of General Internal Medicine, 21, 158-164. | EMBASE | Yes | No | No | Exclude |  |
| GUBRIUM, A., LECKENBY, D., HARVEY, M. W., MARCUS, B. H., ROSAL, M. C. & CHASAN-TABER, L. 2019. Perspectives of health educators and interviewers in a randomized controlled trial of a postpartum diabetes prevention program for Latinas: a qualitative assessment. BMC HEALTH SERVICES RESEARCH, 19. | Web of Science | Yes | Yes | Yes | Include |  |
| GÜEMES-HIDALGO, M. & MUÑOZ-CALVO, M. T. 2015. Obesity in childhood and adolescence. Pediatria Integral, 19, 412-427. | EMBASE | Yes | Yes | Yes | Exclude | Full text unavailable |
| HAGA, S. B., BARRY, W. T., MILLS, R., SVETKEY, L., SUCHINDRAN, S., WILLARD, H. F. & GINSBURG, G. S. 2014. Impact of delivery models on understanding genomic risk for type 2 diabetes. Public Health Genomics, 17, 95-104. | EMBASE | Yes | No | Yes | Include |  |
| HALL, R., SAUKKO, P. M., EVANS, P. H., QURESHI, N. & HUMPHRIES, S. E. 2007. Assessing family history of heart disease in primary care consultations: A qualitative study. Family Practice, 24, 435-442. | PsycINFO | No | Yes | Yes | Include |  |
| HARRINGTON, J. & MORGAN, M. 2016. Understanding kidney transplant patients' treatment choices: The interaction of emotion with medical and social influences on risk preferences. Social Science & Medicine, 155, 43-50. | PsycINFO | No | Yes | No | Exclude |  |
| HAUKKALA, A., VORNANEN, M., HALMESVAARA, O., MARTTILA, M., KÄÄRIÄINEN, H. & PEROLA, M. 2019. Risk perceptions for Type 2 diabetes and coronary heart disease after receiving risk information-participants of P5 FinHealth study. European Journal of Human Genetics, 27, 1803. | EMBASE | No | No | Yes | Exclude | Full text unavailable |
| HAYES, J. F., FOWLER, L. A., BALANTEKIN, K. N., ROTMAN, S. A., ALTMAN, M. & WILFLEY, D. E. 2021. Child and Family Predictors of Relative Weight Change in a Low-Income, School-Based Weight Management Intervention. FAMILIES SYSTEMS & HEALTH, 39, 316-326. | Web of Science | Yes | No | Yes | Include |  |
| HEIDEMAN, W. H., DE WIT, M., MIDDELKOOP, B. J. C., NIERKENS, V., STRONKS, K., VERHOEFF, A. P. & SNOEK, F. J. 2012. DiAlert: a prevention program for overweight first degree relatives of type 2 diabetes patients: results of a pilot study to test feasibility and acceptability. Trials, 13, 178. | Ovid MEDLINE | No | Yes | Yes | Include |  |
| HELBIG, K. L., BERNHARDT, B. A., CONWAY, L. J., VALVERDE, K. D., HELBIG, I. & SPERLING, M. R. 2010. Genetic risk perception and reproductive decision making among people with epilepsy. Epilepsia, 51, 1874-1877. | EMBASE | No | No | No | Exclude |  |
| HELLWIG, L. D., BIESECKER, B. B., LEWIS, K. L., BIESECKER, L. G., JAMES, C. A. & KLEIN, W. M. P. 2018. Ability of Patients to Distinguish Among Cardiac Genomic Variant Subclassifications. Circulation. Genomic and precision medicine, 11, e001975. | Ovid MEDLINE | No | No | No | Exclude | Some limitations in study design and procedures – includes participants who have already been diagnosed with CVD |
| HESHKA, J., PALLESCHI, C., WILSON, B., BREHAUT, J., RUTBERG, J., ETCHEGARY, H., LANGLOIS, N., RODGER, M. & WELLS, P. S. 2008. Cognitive and behavioural effects of genetic testing for thrombophilia. Journal of Genetic Counseling, 17, 288-296. | EMBASE | No | No | No | Exclude |  |
| HICKEY, K. T., SCIACCA, R. R., BIVIANO, A. B., WHANG, W., DIZON, J. M., GARAN, H. & CHUNG, W. K. 2014. The effect of cardiac genetic testing on psychological well-being and illness perceptions. Heart & lung : the journal of critical care, 43, 127-32. | Ovid MEDLINE | No | No | Yes | Exclude | Full text unavailable |
| HICKEY, K. T., TAYLOR, J. Y., SCIACCA, R. R., ABOELELA, S., GONZALEZ, P., CASTILLO, C., HAUSER, N. & FRULLA, A. 2014. Cardiac genetic testing: a single-center pilot study of a Dominican population. Hispanic health care international : the official journal of the National Association of Hispanic Nurses, 12, 183-8. | Ovid MEDLINE | Yes | No | Yes | Include |  |
| HILBERT, A. 2013. Genetic determinism in weight bias reduction. Obesity Facts, 6, 49. | EMBASE | Yes | No | Yes | Include |  |
| HILBERT, A., DIERK, J.-M., CONRADT, M., SCHLUMBERGER, P., HINNEY, A., HEBEBRAND, J. & RIEF, W. 2009. Causal attributions of obese men and women in genetic testing: implications of genetic/biological attributions. Psychology & health, 24, 749-61. | Ovid MEDLINE | No | No | Yes | Include |  |
| HO, J., LEE, A., KAMINSKY, L. & WIRRELL, E. 2008. Self-concept, attitude toward illness and family functioning in adolescents with type 1 diabetes. PAEDIATRICS & CHILD HEALTH, 13, 600-604. | Web of Science | Yes | No | No | Exclude |  |
| HOEDEMAEKERS, E., JASPERS, J. P. C. & VAN TINTELEN, J. P. 2007. The influence of coping styles and perceived control on emotional distress in persons at risk for a hereditary heart disease. American Journal of Medical Genetics, Part A, 143, 1997-2005. | EMBASE | No | No | Yes | Include |  |
| HOEEG, D., CHRISTENSEN, U. & GRABOWSKI, D. 2020. Intra-familial health polarisation: how diverse health concerns become barriers to health behaviour change in families with preschool children and emerging obesity. Sociology of health & illness, 42, 1243-1258. | Ovid MEDLINE | Yes | Yes | Yes | Include |  |
| HOLLIS, J. F., SEXTON, G., CONNOR, S. L., CALVIN, L., PEREIRA, C. & MATARAZZO, J. D. 1984. The family heart dietary intervention program: community response and characteristics of joining and nonjoining families. Preventive medicine, 13, 276-85. | Ovid MEDLINE | No | No | No | Exclude | Full text unavailable |
| HONDA, K. & NEUGUT, A. I. 2004. Associations between perceived cancer risk and established risk factors in a national community sample. Cancer detection and prevention, 28, 1-7. | Ovid MEDLINE | Yes | No | No | Exclude | Focuses on cancer risk |
| HOPKINS, R. J., YOUNG, R. P., HAY, B. A. & GAMBLE, G. D. 2012. Lung cancer risk testing enhances NRT uptake and quit rates in randomly recruited smokers offered a gene-based risk test. American Journal of Respiratory and Critical Care Medicine, 185. | EMBASE | No | No | No | Exclude |  |
| HORNE, J., MADILL, J. & GILLILAND, J. 2017. Incorporating the 'Theory of Planned Behavior' into personalized healthcare behavior change research: A call to action. Personalized Medicine, 14, 521-529. | EMBASE | No | No | Yes | Exclude | Full text unavailable |
| HOVICK, S. R., WILKINSON, A. V., ASHIDA, S., DE HEER, H. D. & KOEHLY, L. M. 2014. The impact of personalized risk feedback on Mexican Americans' perceived risk for heart disease and diabetes. Health education research, 29, 222-34. | Ovid MEDLINE | Yes | No | Yes | Include |  |
| HULKOWER, R., DAVIS, N., SCHECHTER, C. & WALKER, E. 2015. Risk perception of obesity and fast food behavior among obese adults in primary care. Endocrine Reviews, 36. | EMBASE | Yes | No | Yes | Include |  |
| HUNT, A. V., HILTON, D. C. K., VERRALL, C. E., BARLOW-STEWART, K. K., FLEMING, J., WINLAW, D. S. & BLUE, G. M. 2020. "Why and how did this happen?": development and evaluation of an information resource for parents of children with CHD. Cardiology in the young, 30, 346-352. | Ovid MEDLINE | No | No | Yes | Include |  |
| IMES, C. C. 2013. Family history and cardiovascular disease risk in at-risk young adults: A pilot intervention study. 74, ProQuest Information & Learning. | PsycINFO | Yes | Yes | Yes | Include |  |
| IMES, C. C., DOUGHERTY, C. M., LEWIS, F. M. & AUSTIN, M. A. 2016. Outcomes of a Pilot Intervention Study for Young Adults at Risk for Cardiovascular Disease Based on Their Family History. The Journal of cardiovascular nursing, 31, 433-40. | Ovid MEDLINE | Yes | No | Yes | Include |  |
| IMES, C. C., NOVOSEL, L. M. & BURKE, L. E. 2016. Heart Disease Risk and Self-efficacy in Overweight and Obese Adults. JNP-JOURNAL FOR NURSE PRACTITIONERS, 12, 710-716. | Web of Science | Yes | No | Yes | Include |  |
| JAFAR-MOHAMMADI, B. & MCCARTHY, M. I. 2008. Genetics of type 2 diabetes mellitus and obesity - A review. Annals of Medicine, 40, 2-10. | Scopus | No | Yes | Yes | Include |  |
| JAMES, K. M., COWL, C. T., TILBURT, J. C., SINICROPE, P. S., ROBINSON, M. E., FRIMANNSDOTTIR, K. R., TIEDJE, K. & KOENIG, B. A. 2011. Impact of direct-to-consumer predictive genomic testing on risk perception and worry among patients receiving routine care in a preventive health clinic. Mayo Clinic proceedings, 86, 933-40. | Ovid MEDLINE | No | No | Yes | Include |  |
| JEONG, S.-H. 2007. Effects of news about genetics and obesity on controllability attribution and helping behavior. Health communication, 22, 221-8. | Ovid MEDLINE | Yes | No | Yes | Include | Focuses on public views of condition |
| JOSLYN, M. R. & HAIDER-MARKEL, D. P. 2019. Perceived causes of obesity, emotions, and attitudes about Discrimination Policy. Social Science & Medicine, 223, 97-103. | PsycINFO | No | No | Yes | Include | Focuses on public views of obesity |
| JOY, S. V., LIPKUS, I. M. & CHO, A. 2010. Patient interest and response to genetic testing for risk of type 2 diabetes in a primary care setting. Diabetes. | EMBASE | No | No | Yes | Exclude | Full text unavailable |
| JURKOVITZ, C., HYLTON, T. N. & MCCLELLAN, W. M. 2005. Prevalence of family history of kidney disease and perception of risk for kidney disease: a population-based study. American journal of kidney diseases : the official journal of the National Kidney Foundation, 46, 11-7. | Ovid MEDLINE | Yes | No | No | Exclude | Focuses on kidney disease |
| KAPTEIN, A. A., VAN KORLAAR, I. M., CAMERON, L. D., VOSSEN, C. Y., VAN DER MEER, F. J. M. & ROSENDAAL, F. R. 2007. Using the common-sense model to predict risk perception and disease-related worry in individuals at increased risk for venous thrombosis. Health psychology : official journal of the Division of Health Psychology, American Psychological Association, 26, 807-12. | Ovid MEDLINE | No | No | No | Exclude | Focuses on thrombosis |
| KASHANI, M., ELIASSON, A., VERNALIS, M., COSTA, L. & TERHAAR, M. 2013. Improving assessment of cardiovascular disease risk by using family history: an integrative literature review. The Journal of cardiovascular nursing, 28, E18-27. | Ovid MEDLINE | No | No | Yes | Include |  |
| KAYANIYIL, S., ARDERN, C. I., WINSTANLEY, J., PARSONS, C., BRISTER, S., OH, P., STEWART, D. E. & GRACE, S. L. 2009. Degree and correlates of cardiac knowledge and awareness among cardiac inpatients. Patient Education and Counseling, 75, 99-107. | PsycINFO | Yes | No | Yes | Include |  |
| KERR, J. A. C. 1985. Adherence and self-care. Heart and Lung: Journal of Acute and Critical Care, 14, 24-31. | EMBASE | No | Yes | No | Exclude |  |
| KHALID, Y., MALINA, O., ROFIAH, A., LATINAH, M., THAHIRAHTUL, A. Z., ZARIDAH, M. S. & TAN, M. H. 1994. Disease and risk factor perception among patients with coronary artery disease in Kuala Terengganu. The Medical journal of Malaysia, 49, 205-8. | Ovid MEDLINE | Yes | No | Yes | Include |  |
| KIM, J., CHOI, S., KIM, C. J., OH, Y. & SHINN, S. H. 2002. Perception of risk of developing diabetes in offspring of type 2 diabetic patients. The Korean journal of internal medicine, 17, 14-8. | Ovid MEDLINE | Yes | No | Yes | Include |  |
| KINNEAR, F. J., WAINWRIGHT, E., PERRY, R., LITHANDER, F. E., BAYLY, G., HUNTLEY, A., COX, J., SHIELD, J. P. & SEARLE, A. 2019. Enablers and barriers to treatment adherence in heterozygous familial hypercholesterolaemia: a qualitative evidence synthesis. BMJ open, 9, e030290. | Ovid MEDLINE | No | Yes | No | Exclude | Focuses on familial hypercholesterolaemia |
| KLEIN WOOLTHUIS, E. P., DE GRAUW, W. J. C., CARDOL, M., VAN WEEL, C., METSEMAKERS, J. F. M. & BIERMANS, M. C. J. 2013. Patients' and partners' illness perceptions in screen-detected versus clinically diagnosed type 2 diabetes: partners matter! Family practice, 30, 418-25. | Ovid MEDLINE | No | No | No | Exclude | Focuses on route to diagnosis of T2D, rather than genetic/familial risk communication |
| KNOWLES, J., ZARAFSHAR, S., PAVLOVIC, A., GOLDSTEIN, B., KIERNAN, M., TSAI, S., MCCONNELL, M., ABSHER, D., ASHLEY, E., IOANNIDIS, J. & ASSIMES, T. 2016. Impact of a genetic risk score for coronary artery disease in reducing cardiovascular risk: A pilot randomized controlled study. Circulation, 134. | EMBASE | Yes | No | Yes | Include |  |
| KORTHALS, M. & KOMDUUR, R. 2010. Uncertainties of Nutrigenomics and Their Ethical Meaning. JOURNAL OF AGRICULTURAL & ENVIRONMENTAL ETHICS, 23, 435-454. | Web of Science | No | Yes | No | Exclude |  |
| KRISTOFFERSEN, A. E., SIROIS, F. M., STUB, T. & HANSEN, A. H. 2017. Prevalence and predictors of complementary and alternative medicine use among people with coronary heart disease or at risk for this in the sixth Tromso study: a comparative analysis using protection motivation theory. BMC complementary and alternative medicine, 17, 324. | Ovid MEDLINE | No | No | No | Exclude | Focuses on use of alternative medicine in people at risk of/with CAD |
| KUNSCHITZ, E., FRIEDRICH, O. & SIPÖTZ, J. 2018. Causal attribution in patients with Coronary Heart Disease. Wiener Klinische Wochenschrift, 130, S44-S45. | EMBASE | No | No | Yes | Exclude | Full text unavailable |
| L FALCONER, C., SKOW, A., BLACK, J., CROKER, H., SOVIO, U., KESSEL, A., SAXENA, S., VINER, R. & KINRA, S. 2012. The majority of parents of overweight and very overweight children underestimate their child's weight status and weightrelated health risk. Archives of Disease in Childhood, 97, A181. | EMBASE | Yes | No | Yes | Exclude | Focuses on parents' perception of children's weight |
| LASCO, G., MENDOZA, J., RENEDO, A., SEGUIN, M. L., PALAFOX, B., PALILEO-VILLANUEVA, L. M., AMIT, A. M. L., DANS, A. L., BALABANOVA, D. & MCKEE, M. 2020. Nasa dugo('It's in the blood'): lay conceptions of hypertension in the Philippines. BMJ GLOBAL HEALTH, 5. | Web of Science | Yes | Yes | Yes | Include |  |
| LAWAL, T. A., LEWIS, K. L., JOHNSTON, J. J., HEIDLEBAUGH, A. R., NG, D., GASTON-JOHANSSON, F. G., KLEIN, W. M. P., BIESECKER, B. B. & BIESECKER, L. G. 2018. Disclosure of cardiac variants of uncertain significance results in an exome cohort. Clinical genetics, 93, 1022-1029. | Ovid MEDLINE | No | Yes | No | Exclude | Some limitations in study design and procedures – focuses on genetic literacy but unclear how participants’ understanding of the test results have been captured (no standardised questionnaires, but the authors also do not present examples of participants’ open-ended responses?) |
| LEINWEBER, K. A., COLUMBO, J. A., KANG, R., TROOBOFF, S. W. & GOODNEY, P. P. 2019. A Review of Decision Aids for Patients Considering More Than One Type of Invasive Treatment. Journal of Surgical Research, 235, 350-366. | EMBASE | No | No | No | Exclude |  |
| LEROUX, D. 2006. Genetic risk of breast cancer. Reproduction Humaine et Hormones, 19, 89-104. | EMBASE | No | Yes | No | Exclude |  |
| LEWIS, N. M. & BAKER, M. K. 1994. YOUNG-WOMEN WITH OR WITHOUT A FAMILY HISTORY OF CARDIOVASCULAR-DISEASE HAVE SIMILAR DIETARY INTAKES AND ANTHROPOMETRIC MEASUREMENTS. NUTRITION RESEARCH, 14, 1003-1012. | Web of Science | Yes | No | Yes | Exclude | Full text unavailable |
| LIN, J., MARCUM, C. S., MYERS, M. F. & KOEHLY, L. M. 2017. Put the family back in family health history: A multiple-informant approach. American Journal of Preventive Medicine, 52, 640-644. | PsycINFO | Yes | No | Yes | Include |  |
| LIN, J., MARCUM, C. S., WILKINSON, A. V. & KOEHLY, L. M. 2018. Developing Shared Appraisals of Diabetes Risk Through Family Health History Feedback: The Case of Mexican-Heritage Families. Annals of behavioral medicine : a publication of the Society of Behavioral Medicine, 52, 262-271. | Ovid MEDLINE | Yes | No | Yes | Include |  |
| LINDSAY, A. C., ARRUDA, C. A. M., MACHADO, M. M. T., DE ANDRADE, G. P. & GREANEY, M. L. 2018. Exploring Brazilian Immigrant Mothers' Beliefs, Attitudes, and Practices Related to Their Preschool-Age Children's Sleep and Bedtime Routines: A Qualitative Study Conducted in the United States. INTERNATIONAL JOURNAL OF ENVIRONMENTAL RESEARCH AND PUBLIC HEALTH, 15. | Web of Science | Yes | Yes | No | Exclude |  |
| LINDSAY, A. C., WALLINGTON, S. F., LEES, F. D. & GREANEY, M. L. 2018. Exploring How the Home Environment Influences Eating and Physical Activity Habits of Low-Income, Latino Children of Predominantly Immigrant Families: A Qualitative Study. INTERNATIONAL JOURNAL OF ENVIRONMENTAL RESEARCH AND PUBLIC HEALTH, 15. | Web of Science | Yes | Yes | Yes | Include |  |
| LIPPA, N. C. & SANDERSON, S. C. 2012. Impact of information about obesity genomics on the stigmatization of overweight individuals: An experimental study. Obesity, 20, 2367-2376. | PsycINFO | Yes | No | Yes | Include | Focuses on public views of condition |
| LOUZADA, M. L., TALJAARD, M., LANGLOIS, N. J., KAHN, S. R., RODGER, M. A., ANDERSON, D. R., KOVACS, M. J. & WELLS, P. S. 2011. Psychological impact of thrombophilia testing in asymptomatic family members. Thrombosis research, 128, 530-5. | Ovid MEDLINE | No | No | No | Exclude | Focuses on thrombophilia |
| MACCIOCCA, I., UKOUMUNNE, O., DAVIS, A., WEINTRAUB, R., CONNELL, V., INGLES, J., DELATYCKI, M., WAKE, S., SEMSARIAN, C., YEATES, L., MCKENNA, W., MARTEAU, T., WINSHIP, I. & COLLINS, V. 2010. Understanding, risk perception and psychological outcomes three months after predictive gene testing for hypertrophic cardiomyopathy and long QT syndrome. Twin Research and Human Genetics, 13, 651. | EMBASE | Yes | No | No | Exclude | Focuses on cardiomyopathy and long QT syndrome |
| MACKIE, T. I., TSE, L. L., DE FERRANTI, S. D., RYAN, H. R. & LESLIE, L. K. 2015. Treatment decision making for adolescents with familial hypercholesterolemia: Role of family history and past experiences. JOURNAL OF CLINICAL LIPIDOLOGY, 9, 583-593. | Web of Science | Yes | Yes | No | Exclude | Focuses on familial hypercholesterolaemia |
| MANUEL, A. & BRUNGER, F. 2015. “Awakening to” a new meaning of being at-risk for arrhythmogenic right ventricular cardiomyopathy: a grounded theory study. Journal of Community Genetics, 6, 167-175. | EMBASE | No | Yes | No | Exclude |  |
| MARATHE, J., OGDEN, K. & WOODROFFE, J. 2017. Exploring the lived experience of patients with inherited cardiac conditions. Heart Lung and Circulation, 26, S305. | EMBASE | No | Yes | Yes | Exclude | Focuses on cardiomyopathy |
| MARKOWITZ, S. M., PARK, E. R., DELAHANTY, L. M., O'BRIEN, K. E. & GRANT, R. W. 2011. Perceived impact of diabetes genetic risk testing among patients at high phenotypic risk for type 2 diabetes. Diabetes care, 34, 568-73. | Ovid MEDLINE | No | Yes | Yes | Include |  |
| MARRERO, W. J., LAVIERI, M. S. & SUSSMAN, J. B. 2021. Optimal cholesterol treatment plans and genetic testing strategies for cardiovascular diseases. Health care management science, 24, 1-25. | Ovid MEDLINE | No | No | No | Exclude | Focuses on health trajectory modelling |
| MARRERO, W. J., LAVIERI, M. S., SUSSMAN, J. B. & IEEE 2019. A SIMULATION MODEL TO EVALUATE THE IMPLICATIONS OF GENETIC TESTING IN CHOLESTEROL TREATMENT PLANS. 2019 WINTER SIMULATION CONFERENCE (WSC). | Web of Science | No | No | No | Exclude |  |
| MARTEAU, T. M., KINMONTH, A. L., PYKE, S. & THOMPSON, S. G. 1995. READINESS FOR LIFE-STYLE ADVICE - SELF-ASSESSMENTS OF CORONARY RISK PRIOR TO SCREENING IN THE BRITISH FAMILY HEART-STUDY. BRITISH JOURNAL OF GENERAL PRACTICE, 45, 5-8. | Web of Science | No | No | Yes | Include |  |
| MARTEAU, T. M. & WEINMAN, J. 2006. Self-regulation and the behavioural response to DNA risk information: A theoretical analysis and framework for future research. Social Science and Medicine, 62, 1360-1368. | EMBASE | No | Yes | Yes | Include |  |
| MARTINEZ, A. M. S., FRANCO, H. F. C., DE LEON, D. B. E., FRANCO, G. E. M., DE LA GARZA, F. J. G. & ROCHA, G. M. N. 2020. Estimating and differentiating maternal feeding practices in a country ranked first in childhood obesity. PUBLIC HEALTH NUTRITION, 23, 620-630. | Web of Science | Yes | No | No | Exclude |  |
| MATLOFF, E. T., MOYER, A., SHANNON, K. M., NIENDORF, K. B. & COL, N. F. 2006. Healthy women with a family history of breast cancer: Impact of a tailored genetic counseling intervention on risk perception, knowledge, and menopausal therapy decision making. Journal of Women's Health, 15, 843-856. | EMBASE | No | No | No | Exclude |  |
| MAZZA, E., LA MILIA, D. I., GALLETTI, C., GAMBIOLI, S., MOSCATO, U., DAMIANI, G. & LAURENTI, P. 2017. [Risk perception and biological parameters related to obesity in a sample population of the city of Rome, Italy: nurses' contribution to supporting health-related decisions]. Percezione del rischio e parametri biologici correlati all'obesita in una popolazione campione della citta di Roma. Il contributo dell'infermiere a supporto delle decisioni per la salute., 73, 215-234. | Ovid MEDLINE | No | No | No | Exclude | Full text unavailable |
| MCVAY, M. A., BEADLES, C., WU, R., GRUBBER, J., COFFMAN, C. J., YANCY, W. S., REINER, I. L. & VOILS, C. I. 2015. Effects of provision of type 2 diabetes genetic risk feedback on patient perceptions of diabetes control and diet and physical activity self-efficacy. Patient education and counseling, 98, 1600‐1607. | Cochrane Central Register of Controlled Trials (CENTRAL) | Yes | No | Yes | Include |  |
| MELIN, J., MAZIARZ, M., ARONSSON, C. A., LUNDGREN, M. & LARSSON, H. E. 2020. Parental anxiety after 5 years of participation in a longitudinal study of children at high risk of type 1 diabetes. PEDIATRIC DIABETES, 21, 878-889. | Web of Science | Yes | No | No | Exclude |  |
| MEMETOVIC, J., ROBINSON, T., MACDONALD, G., LEESE, J., KOEHN, C. & LI, L. 2016. Examining perceived risk factors of arthritis: Findings from a public opinion survey. Journal of Rheumatology, 43, 1182-1183. | EMBASE | No | No | No | Exclude |  |
| MEULENKAMP, T. M., TIBBEN, A., MOLLEMA, E. D., VAN LANGEN, I. M., WIEGMAN, A., DE WERT, G. M., DE BEAUFORT, I. D., WILDE, A. A. M. & SMETS, E. M. A. 2008. Predictive genetic testing for cardiovascular diseases: impact on carrier children. American journal of medical genetics. Part A, 146A, 3136-46. | Ovid MEDLINE | Yes | Yes | Yes | Include |  |
| MIDDLEMASS, J. B., YAZDANI, M. F., KAI, J., STANDEN, P. J. & QURESHI, N. 2014. Introducing genetic testing for cardiovascular disease in primary care: a qualitative study. The British journal of general practice : the journal of the Royal College of General Practitioners, 64, e282-9. | Ovid MEDLINE | No | Yes | Yes | Include |  |
| MORAN, M. T., MAZZOCCO, V. E., FISCUS, W. G. & KOZA, E. P. 1989. Coronary heart disease risk assessment. American Journal of Preventive Medicine, 5, 330-336. | PsycINFO | No | No | Yes | Exclude | Some limitations in study design – unsure why recruiting people who have already been diagnosed with CVD? |
| MORRILL, K. E. 2021. Informing the development of a culturally-sensitive, genotype-informed intervention for treatment of nonalcoholic fatty liver disease in Mexican-origin women. 82, ProQuest Information & Learning. | PsycINFO | Yes | Yes | No | Exclude |  |
| MUNOZ, L. R., ETNYRE, A., ADAMS, M., HERBERS, S., WITTE, A., HORLEN, C., BAYNTON, S., ESTRADA, R. & JONES, M. E. 2010. Awareness of heart disease among female college students. Journal of women's health (2002), 19, 2253-9. | Ovid MEDLINE | Yes | No | Yes | Include |  |
| MURPHY, B., WORCESTER, M., HIGGINS, R., LE GRANDE, M., LARRITT, P. & GOBLE, A. 2005. Causal attributions for coronary heart disease among female cardiac patients. Journal of Cardiopulmonary Rehabilitation, 25, 135-145. | Scopus | No | No | Yes | Include |  |
| NELSON, H. D., HUFFMAN, L. H., FU, R. & HARRIS, E. L. 2005. Genetic risk assessment and BRCA mutation testing for breast and ovarian cancer susceptibility: Systematic evidence review for the U.S. Preventive Services Task Force. Annals of Internal Medicine, 143, 362-379+I-47. | EMBASE | No | Yes | No | Exclude |  |
| NICHOLS, G. J. 1995. Testing a culturally consistent behavioral outcomes strategy for cardiovascular disease risk reduction and prevention in low income African American women. 56, ProQuest Information & Learning. | PsycINFO | No | No | No | Exclude | Focuses on cardiomyopathy |
| NIEUWHOF, K., BIRNIE, E., VAN DEN BERG, M. P., DE BOER, R. A., VAN HAELST, P. L., VAN TINTELEN, J. P. & VAN LANGEN, I. M. 2017. Follow-up care by a genetic counsellor for relatives at risk for cardiomyopathies is cost-saving and well-appreciated: a randomised comparison. European journal of human genetics : EJHG, 25, 169-175. | Ovid MEDLINE | Yes | No | Yes | Include |  |
| NISHIGAKI, M., KOBAYASHI, K., KATO, N., SEKI, N., YOKOMURA, T., YOKOYAMA, M. & KAZUMA, K. 2009. Preventive advice given by patients with type 2 diabetes to their offspring. The British journal of general practice : the journal of the Royal College of General Practitioners, 59, 37-42. | Ovid MEDLINE | No | No | Yes | Include |  |
| NISHIGAKI, M., TOKUNAGA-NAKAWATASE, Y., NISHIDA, J. & KAZUMA, K. 2014. The effect of genetic counseling for adult offspring of patients with type 2 diabetes on attitudes toward diabetes and its heredity: a randomized controlled trial. Journal of genetic counseling, 23, 762-9. | Ovid MEDLINE | Yes | No | No | Exclude | Some limitations in study design – different tutors delivering experimental conditions; also unsure if study design is best suited for the purposes of its research questions |
| O'BRIEN, K. S., PUHL, R. M., LATNER, J. D., MIR, A. S. & HUNTER, J. A. 2010. Reducing anti-fat prejudice in preservice health students: a randomized trial. Obesity (Silver Spring, Md.), 18, 2138-44. | Ovid MEDLINE | No | No | No | Exclude | Focuses on cardiomyopathy |
| O'DONOVAN, C. E., SKINNER, J. R. & BROADBENT, E. 2020. Perceptions of Risk of Cardiac Arrest in Individuals Living With a Cardiac Inherited Disease: Are the Doctor and the Patient on the Same Page? Heart Lung and Circulation, 29, 851-858. | EMBASE | Yes | No | Yes | Include |  |
| OGDEN, J., DALKOU, M., KOUSANTONI, M., VENTURA, S. S. & REYNOLDS, R. 2017. Body weight, the home environment, and eating behaviour across three generations of women: A quasi‐longitudinal study in four Mediterranean and non‐Mediterranean countries. Australian Psychologist, 52, 442-452. | PsycINFO | Yes | Yes | Yes | Include |  |
| OLIVERI, S., FERRARI, F., MANFRINATI, A. & PRAVETTONI, G. 2018. A systematic review of the psychological implications of genetic testing: A comparative analysis among cardiovascular, neurodegenerative and cancer diseases. Frontiers in Genetics, 9. | EMBASE | Yes | Yes | Yes | Include |  |
| ORMONDROYD, E., OATES, S., PARKER, M., BLAIR, E. & WATKINS, H. 2014. Pre-symptomatic genetic testing for inherited cardiac conditions: a qualitative exploration of psychosocial and ethical implications. European journal of human genetics : EJHG, 22, 88-93. | Ovid MEDLINE | Yes | Yes | No | Exclude | Focuses on hypertrophic cardiomyopathy and long QT syndrome |
| OROM, H., SCHOFIELD, E., KIVINIEMI, M. T., WATERS, E. A. & HAY, J. L. 2021. Agency beliefs are associated with lower health information avoidance. Health Education Journal, 80, 272-286. | EMBASE | No | No | Yes | Include |  |
| OSUJI, N. A., OJO, O. S., MALOMO, S. O., SOGUNLE, P. T., EGUNJOBI, A. O. & ODEBUNMI, O. O. 2018. Relationship between glycemic control and perceived family support among people with type 2 diabetes mellitus seen in a rich kinship network in Southwest Nigeria. FAMILY MEDICINE AND COMMUNITY HEALTH, 6, 168-177. | Web of Science | Yes | No | Yes | Include |  |
| OTTEN, E., BIRNIE, E., RANCHOR, A. V., VAN TINTELEN, J. P. & VAN LANGEN, I. M. 2015. A group approach to genetic counselling of cardiomyopathy patients: satisfaction and psychological outcomes sufficient for further implementation. European journal of human genetics : EJHG, 23, 1462-7. | Ovid MEDLINE | No | No | No | Exclude | Focuses on cardiomyopathy |
| PAPPA, E., KONTODIMOPOULOS, N., PAPADOPOULOS, A. A., PALLIKARONA, G., NIAKAS, D. & TOUNTAS, Y. 2009. Factors Affecting Use of Preventive Tests for Cardiovascular Risk among Greeks. INTERNATIONAL JOURNAL OF ENVIRONMENTAL RESEARCH AND PUBLIC HEALTH, 6, 2712-2724. | Web of Science | Yes | No | Yes | Include |  |
| PEARLSON, G. D. & FOLLEY, B. S. 2008. Endophenotypes, dimensions, risks: Is psychosis analogous to common inherited medical illnesses? CLINICAL EEG AND NEUROSCIENCE, 39, 73-77. | Web of Science | No | No | No | Exclude |  |
| PELEG, O., HADAR, E. & COHEN, A. 2020. Individuals with type 2 diabetes: An exploratory study of their experience of family relationships and coping with the illness. The Diabetes Educator, 46, 83-93. | PsycINFO | Yes | Yes | Yes | Include |  |
| PERSKY, S., BOUHLAL, S., GOLDRING, M. R. & MCBRIDE, C. M. 2017. Beliefs about genetic influences on eating behaviors: Characteristics and associations with weight management confidence. EATING BEHAVIORS, 26, 93-98. | Web of Science | Yes | No | Yes | Include |  |
| PERSKY, S. & ECCLESTON, C. P. 2011. Impact of Genetic Causal Information on Medical Students' Clinical Encounters with an Obese Virtual Patient: Health Promotion and Social Stigma. ANNALS OF BEHAVIORAL MEDICINE, 41, 363-372. | Web of Science | No | No | Yes | Include |  |
| PERSKY, S., GOLDRING, M. R., EL-TOUKHY, S., FERRER, R. A. & HOLLISTER, B. 2019. Parental Defensiveness about Multifactorial Genomic and Environmental Causes of Children's Obesity Risk. Childhood obesity (Print), 15, 289-297. | Ovid MEDLINE | No | No | Yes | Include |  |
| PERSKY, S. & YAREMYCH, H. E. 2020. Parents' genetic attributions for children's eating behaviors: Relationships with beliefs, emotions, and food choice behavior. APPETITE, 155. | Web of Science | Yes | No | Yes | Include |  |
| PESCH, M. H., WENTZ, E. E., ROSENBLUM, K. L., APPUGLIESE, D. P., MILLER, A. L. & LUMENG, J. C. 2015. "You've got to settle down!": Mothers' perceptions of physical activity in their young children. BMC PEDIATRICS, 15. | Web of Science | No | Yes | No | Exclude |  |
| PETR, E. J., AYERS, C. R., PANDEY, A., DE LEMOS, J. A., POWELL-WILEY, T. M., KHERA, A., LLOYD-JONES, D. M. & BERRY, J. D. 2014. Perceived Lifetime Risk for Cardiovascular Disease (from the Dallas Heart Study). AMERICAN JOURNAL OF CARDIOLOGY, 114, 53-58. | Web of Science | No | No | Yes | Include |  |
| PETRIČEK, G., VULETĆ MAVRINAC, G. & VRCIĆ-KEGLEVIĆ, M. 2009. Health locus of control assessment in diabetes mellitus type 2 patients. Acta Medica Croatica, 63, 135-143. | Scopus | No | No | Yes | Exclude | Full text unavailable |
| PICCININO, L., GRIFFEY, S., GALLIVAN, J., LOTENBERG, L. D. & TUNCER, D. 2015. Recent Trends in Diabetes Knowledge, Perceptions, and Behaviors: Implications for National Diabetes Education. Health education & behavior : the official publication of the Society for Public Health Education, 42, 687-96. | Ovid MEDLINE | Yes | Yes | Yes | Include |  |
| PIERCE, M., HARDING, D., RIDOUT, D., KEEN, H. & BRADLEY, C. 2001. Risk and prevention of type II diabetes: Offspring's views. British Journal of General Practice, 51, 194-199. | Scopus | Yes | No | Yes | Include |  |
| PIERCE, M., HAYWORTH, J., WARBURTON, F., KEEN, H. & BRADLEY, C. 1999. Diabetes mellitus in the family: Perceptions of offspring's risk. Diabetic Medicine, 16, 431-436. | Scopus | Yes | No | Yes | Include |  |
| PIERCE, M., RIDOUT, D., HARDING, D., KEEN, H. & BRADLEY, C. 2000. More good than harm: a randomised controlled trial of the effect of education about familial risk of diabetes on psychological outcomes. The British journal of general practice : the journal of the Royal College of General Practitioners, 50, 867-71. | Ovid MEDLINE | No | Yes | Yes | Include |  |
| PODURI, A. & GRISSO, J. A. 1998. Cardiovascular risk factors in economically disadvantaged women: A study of prevalence and awareness. Journal of the National Medical Association, 90, 531-536. | PsycINFO | Yes | No | Yes | Include |  |
| POLACSEK, M., ORR, J., O'BRIEN, L. M., ROGERS, V. W., FANBURG, J. & GORTMAKER, S. L. 2014. Sustainability of Key Maine Youth Overweight Collaborative Improvements: A Follow-Up Study. CHILDHOOD OBESITY, 10, 326-333. | Web of Science | Yes | No | Yes | Include |  |
| POLLEY, B. A., JAKICIC, J. M., VENDITTI, E. M., BARR, S. & WING, R. R. 1997. The effects of health beliefs on weight loss in individuals at high risk for NIDDM. Diabetes Care, 20, 1533-1538. | Scopus | No | No | Yes | Include |  |
| PREDHAM, S., HATHAWAY, J., HULAIT, G., ARBOUR, L. & LEHMAN, A. 2017. Patient Recall, Interpretation, and Perspective of an Inconclusive Long QT Syndrome Genetic Test Result. Journal of genetic counseling, 26, 150-158. | Ovid MEDLINE | No | Yes | No | Exclude | Focuses on long QT syndrome |
| RAZALI, S., ISMAIL, Z., ABDULLAH, N. & NAWAWI, H. M. 2019. Illness Perception, Level of Education and Presence of Cardiovascular Disease among Patients with Familial Hypercholesterolaemia. ENVIRONMENT-BEHAVIOUR PROCEEDINGS JOURNAL. | Web of Science | Yes | No | Yes | Include |  |
| REGO, S., DAGAN‐ROSENFELD, O., BIVONA, S. A., SNYDER, M. P. & ORMOND, K. E. 2019. Much ado about nothing: A qualitative study of the experiences of an average‐risk population receiving results of exome sequencing. Journal of Genetic Counseling, 28, 428-437. | PsycINFO | No | Yes | Yes | Include |  |
| REID, G., WALTER, F. & EMERY, J. 2011. Assessing the psychosocial impact of family history screening in the australian primary care setting. Familial Cancer, 10, S86. | EMBASE | Yes | Yes | Yes | Include |  |
| REYNA, V. F. 2008. A theory of medical decision making and health: Fuzzy trace theory. Medical Decision Making, 28, 850-865. | EMBASE | No | Yes | Yes | Include |  |
| RINKEL, G. J. E. 2009. Screening and management of unruptured cerebral aneurysms. Journal of Neurology, 256, S4. | EMBASE | No | No | No | Exclude |  |
| ROBINS, J. L. W., MCCAIN, N. L. & ELSWICK, R. K. 2012. Exploring the Complexity of Cardiometabolic Risk in Women. BIOLOGICAL RESEARCH FOR NURSING, 14, 160-170. | Web of Science | No | No | No | Exclude | Some limitations in study design – small sample size for study goals and purposes |
| ROBINSON, C. L., JOUNI, H., KRUISSELBRINK, T. M., AUSTIN, E. E., CHRISTENSEN, K. D., GREEN, R. C. & KULLO, I. J. 2016. Disclosing genetic risk for coronary heart disease: effects on perceived personal control and genetic counseling satisfaction. Clinical genetics, 89, 251-7. | Ovid MEDLINE | No | No | Yes | Include |  |
| ROBINSON, C. L., JOUNI, H., KRUISSELBRINK, T. M., CHRISTENSEN, K. D., GREEN, R. C. & KULLO, I. J. 2014. The effect of disclosing genetic risk for coronary heart disease on perceived personal control and genetic counseling satisfaction: The MI-genes study. Circulation, 130. | EMBASE | No | No | Yes | Include |  |
| ROBINSON, D. & ALLAWAY, S. 1998. Health risk appraisal in the UK--some preliminary results. Methods of information in medicine, 37, 143-6. | Ovid MEDLINE | No | No | No | Exclude | Full text unavailable |
| RODRIGUES, A. & CARRINGTON, M. 2017. Relationship between cardio-metabolic disease risk and health beliefs, perceptions and behaviours: A regional perspective. Heart Lung and Circulation, 26, S231. | EMBASE | No | No | Yes | Include |  |
| ROGERS, N. T., WATERLOW, N. R., BRINDLE, H., ENRIA, L., EGGO, R. M., LEES, S. & ROBERTS, C. H. 2020. Behavioral Change Towards Reduced Intensity Physical Activity Is Disproportionately Prevalent Among Adults With Serious Health Issues or Self-Perception of High Risk During the UK COVID-19 Lockdown. FRONTIERS IN PUBLIC HEALTH, 8. | Web of Science | No | No | No | Exclude |  |
| SAMUEL-HARRIS, S. N. 2021. Perceptions of health and family history associations in African American men at risk for cardiovascular disease. 82, ProQuest Information & Learning. | PsycINFO | Yes | No | No | Exclude | Full text unavailable |
| SANDERSON, S. C., PERSKY, S. & MICHIE, S. 2010. Psychological and behavioral responses to genetic test results indicating increased risk of obesity: does the causal pathway from gene to obesity matter? Public health genomics, 13, 34-47. | Ovid MEDLINE | Yes | No | Yes | Include |  |
| SANTOS, R. D., PEREIRA, C., CESENA, F., LAURINAVICIUS, A. G., TABONE, V. & BITTENCOURT, M. S. 2021. Cardiovascular Risk Misperception and Low Awareness of Familial Hypercholesterolemia in Individuals with Severe Hypercholesterolemia. Percepcao Inadequada do Risco Cardiovascular e Baixo Conhecimento sobre Hipercolesterolemia Familiar em Individuos com Hipercolesterolemia Grave., 116, 706-712. | Ovid MEDLINE | Yes | No | No | Exclude | Focuses on familial hypercholesterolaemia |
| SAUKKO, P. M., RICHARDS, S. H., SHEPHERD, M. H. & CAMPBELL, J. L. 2006. Are genetic tests exceptional? Lessons from a qualitative study on thrombophilia. Social Science and Medicine, 63, 1947-1959. | EMBASE | No | Yes | No | Exclude |  |
| SCALZI, L. V., BALLOU, S. P., PARK, J. Y., REDLINE, S. & KIRCHNER, H. L. 2008. Cardiovascular disease risk awareness in systemic lupus erythematosus patients. Arthritis and rheumatism, 58, 1458-64. | Ovid MEDLINE | No | No | Yes | Include |  |
| SCHNEIDER, K. I. & SCHMIDTKE, J. 2014. Patient compliance based on genetic medicine: A literature review. Journal of Community Genetics, 5, 31-48. | EMBASE | No | Yes | Yes | Include |  |
| SCOLLAN-KOLIOPOULOS, M. 2005. Type 2 diabetes illness representation, self-care, and multigenerational legacies of diabetes: Three reports. 66, ProQuest Information & Learning. | PsycINFO | No | No | Yes | Include |  |
| SEABORN, C., SUTHER, S., LEE, T., KIROS, G. E., BECKER, A., CAMPBELL, E. & COLLINS-ROBINSON, J. 2016. Utilizing Genomics through Family Health History with the Theory of Planned Behavior: Prediction of Type 2 Diabetes Risk Factors and Preventive Behavior in an African American Population in Florida. PUBLIC HEALTH GENOMICS, 19, 69-80. | Web of Science | Yes | No | Yes | Include |  |
| SEABORN, C. A. E. 2016. Introducing genomics via family history utilizing the theory of planned behavior: An assessment of type 2 diabetes mellitus risk factors and its influence on healthy behavior in an African American population in Florida. 76, ProQuest Information & Learning. | PsycINFO | Yes | No | Yes | Include |  |
| SEGAL, M. E., POLANSKY, M. & SANKAR, P. 2007. Predictors of uptake of obesity genetic testing among affected adults. HUMAN GENETICS, 120, 641-652. | Web of Science | Yes | Yes | Yes | Include |  |
| SEGAL, M. E., SANKAR, P. & REED, D. R. 2004. Research issues in genetic testing of adolescents for obesity. NUTRITION REVIEWS, 62, 307-320. | Web of Science | Yes | Yes | Yes | Include | Focuses on ethics |
| SENIOR, V. & MARTEAU, T. M. 2007. Causal attributions for raised cholesterol and perceptions of effective risk-reduction: Self-regulation strategies for an increased risk of coronary heart disease. Psychology & Health, 22, 699-717. | PsycINFO | No | No | Yes | Include |  |
| SENIOR, V., WEINMAN, J. & MARTEAU, T. M. 2002. The influence of perceived control over causes and responses to health threats: A vignette study. British Journal of Health Psychology, 7, 203-211. | EMBASE | Yes | No | Yes | Include | Focuses on public views of conditions |
| SHAH, L. L., PERKHOUNKOVA, Y. & DAACK-HIRSCH, S. 2016. Evaluation of the Perception of Risk Factors for Type 2 Diabetes Instrument in an At-Risk, Nondiabetic Population. JOURNAL OF NURSING MEASUREMENT, 24, E83-E100. | Web of Science | No | No | Yes | Exclude | Focuses on instrument development |
| SHEN, Y., WANG, T. T., GAO, M., HU, K., ZHU, X. R., ZHANG, X., WANG, F. B., HE, C. & SUN, X. Y. 2020. [Effectiveness evaluation of health belief model-based health education intervention for patients with hypertension in community settings]. Zhonghua yu fang yi xue za zhi [Chinese journal of preventive medicine], 54, 155-159. | Ovid MEDLINE | Yes | No | No | Exclude | Full text unavailable |
| SHEPHERD, M., SPARKES, A. C. & HATTERSLEY, A. T. 2001. Genetic testing in maturity onset diabetes of the young (MODY): A new challenge for the diabetic clinic. Practical Diabetes International, 18, 16-21. | EMBASE | Yes | Yes | No | Exclude | Focuses on MODY |
| SHERMAN, K., CAMERON, L., BROWN, P. & MARTEAU, T. 2009. Effect of worry and monitoring processing style on cognitive and affective responses to genetic risk information. Psycho-Oncology, 18, S139-S140. | EMBASE | No | No | Yes | Include |  |
| SHERMAN, K., SHAW, L.-K., CHAMPION, K., CALDEIRA, F. & MCCASKILL, M. 2015. The effect of disease risk probability and disease type on interest in clinic-based versus direct-to-consumer genetic testing services. Journal of behavioral medicine, 38, 706-14. | Ovid MEDLINE | Yes | No | Yes | Include |  |
| SHILOH, S., DEHEER, H. D., PELEG, S., HENSLEY ALFORD, S., SKAPINSKY, K., ROBERTS, J. S. & HADLEY, D. W. 2015. The impact of multiplex genetic testing on disease risk perceptions. Clinical Genetics, 87, 117-123. | EMBASE | Yes | No | Yes | Include |  |
| SHILOH, S., WADE, C. H., ROBERTS, J. S., ALFORD, S. H. & BIESECKER, B. B. 2013. Associations between risk perceptions and worry about common diseases: a between- and within-subjects examination. Psychology & health, 28, 434-49. | Ovid MEDLINE | Yes | No | Yes | Include |  |
| SILVA, L., CONDON, L., KAI, J., WENG, S., VEDHARA, K. & QURESHI, N. 2019. Patient's experiences of genomic testing for Familial Hypercholesterolaemia: What it might reveal about the adoption of healthy lifestyle behaviours. European Journal of Human Genetics, 27, 1798. | EMBASE | No | Yes | Yes | Exclude | Full text unavailable |
| SILVERMAN, K. R., OHMAN-STRICKLAND, P. A. & CHRISTIAN, A. H. 2017. Perceptions of Cancer Risk: Differences by Weight Status. Journal of cancer education : the official journal of the American Association for Cancer Education, 32, 357-363. | Ovid MEDLINE | Yes | No | No | Exclude | Focuses on cancer risk |
| SMITH, L. B., LYNCH, K. F., DRISCOLL, K. A., JOHNSON, S. B. & GRP, T. S. 2021. Parental monitoring for type 1 diabetes in genetically at-risk young children: The TEDDY study. PEDIATRIC DIABETES, 22, 717-728. | Web of Science | Yes | No | No | Exclude |  |
| SOHAL, P. S. 2008. Prevention and management of diabetes in South Asians. Canadian Journal of Diabetes, 32, 206-210. | Scopus | Yes | Yes | Yes | Exclude | Full text unavailable |
| STACEY, D., LÉGARÉ, F., COL, N. F., BENNETT, C. L., BARRY, M. J., EDEN, K. B., HOLMES-ROVNER, M., LLEWELLYN-THOMAS, H., LYDDIATT, A., THOMSON, R., TREVENA, L. & WU, J. H. C. 2014. Decision aids for people facing health treatment or screening decisions. Cochrane Database of Systematic Reviews, 2014. | EMBASE | No | Yes | No | Exclude |  |
| STEWART, S., FERRY, A., STRACHAN, F., JIN, K., NEUBECK, L. & MILLS, N. 2020. Cardiovascular risk communication strategies in primary prevention. A mixed methods systematic review. European Journal of Cardiovascular Nursing, 19, S14-S15. | EMBASE | No | Yes | Yes | Include | Full text unavailable |
| STOL, D. M., HOLLANDER, M., DAMMAN, O. C., NIELEN, M. M. J., BADENBROEK, I. F., SCHELLEVIS, F. G. & DE WIT, N. J. 2020. Mismatch between self-perceived and calculated cardiometabolic disease risk among participants in a prevention program for cardiometabolic disease: a cross-sectional study. BMC public health, 20, 740. | Ovid MEDLINE | No | No | Yes | Include |  |
| STUTTGEN, K., PACYNA, J., KULLO, I. & SHARP, R. 2020. Neutral, negative, or negligible? Changes in patient perceptions of disease risk following receipt of a negative genomic screening result. Journal of Personalized Medicine, 10. | EMBASE | No | No | Yes | Include |  |
| SUBAS, T., LUITEN, R., HANSON‐KAHN, A., WHEELER, M. & CALESHU, C. 2019. Evolving decisions: Perspectives of active and athletic individuals with inherited heart disease who exercise against recommendations. Journal of Genetic Counseling, 28, 119-129. | PsycINFO | No | Yes | No | Exclude |  |
| SWARTLING, U., LYNCH, K., SMITH, L., JOHNSON, S. B. & GRP, T. S. 2016. Parental Estimation of Their Child's Increased Type 1 Diabetes Risk During the First 2 Years of Participation in an International Observational Study: Results From the TEDDY study. JOURNAL OF EMPIRICAL RESEARCH ON HUMAN RESEARCH ETHICS, 11, 106-114. | Web of Science | Yes | No | No | Exclude |  |
| SWEET, K., GORDON, E. S., STURM, A. C., SCHMIDLEN, T. J., MANICKAM, K., TOL, A. E., KELLER, M. A., STACK, C. B., FELIPE GARCÍA-ESPAÑA, J., BELLAFANTE, M., TAYAL, N., EMBI, P., BINKLEY, P., HERSHBERGER, R. E., SADEE, W., CHRISTMAN, M. & MARSH, C. 2014. Design and implementation of a randomized controlled trial of genomic counseling for patients with chronic disease. Journal of Personalized Medicine, 4, 1-19. | EMBASE | No | No | Yes | Include |  |
| TAMRAGOURI, R. N., MARTIN, R. W., CLEAVENGER, R. L. & SIEBER, W. K. 1986. Cardiovascular risk factors and health knowledge among freshman college students with a family history of cardiovascular disease. Journal of American College Health, 34, 267-270. | PsycINFO | Yes | No | Yes | Exclude | Some limitations in study design – unequal comparisons between groups? |
| TANG, J. W., CAMERON, K. A., PUMARINO, J., PEACEMAN, A. & ACKERMANN, R. T. 2013. Perceived risk for type 2 diabetes among women with a history of gestational diabetes. Journal of General Internal Medicine, 28, S144. | EMBASE | Yes | No | Yes | Include |  |
| TAYLOR, J. Y., SUN, Y. V., BARCELONA DE MENDOZA, V., IFATUNJI, M., RAFFERTY, J., FOX, E. R., MUSANI, S. K., SIMS, M. & JACKSON, J. S. 2017. The combined effects of genetic risk and perceived discrimination on blood pressure among African Americans in the Jackson Heart Study. Medicine, 96, e8369. | Ovid MEDLINE | Yes | No | No | Exclude | Focuses on SNP x perceived race/ethnic discrimination interactions in influencing clinically relevant traits |
| TURNWALD, B. P., GOYER, J. P., BOLES, D. Z., SILDER, A., DELP, S. L. & CRUM, A. J. 2019. Learning one's genetic risk changes physiology independent of actual genetic risk. Nature human behaviour, 3, 48-56. | Ovid MEDLINE | No | No | Yes | Include |  |
| TURRINI, M. & BOURGAIN, C. 2021. Appraising screening, making risk in/visible. The medical debate over Non-Rare Thrombophilia (NRT) testing before prescribing the pill. Sociology of health & illness, 43, 1627-1642. | EMBASE | No | Yes | No | Exclude |  |
| VAN ESCH, S. C. M., CORNEL, M. C., GEELHOED-DUIJVESTIJN, P. H. L. M. & SNOEK, F. J. 2012. Family communication as strategy in diabetes prevention: an observational study in families with Dutch and Surinamese South-Asian ancestry. Patient education and counseling, 87, 23-9. | Ovid MEDLINE | Yes | No | Yes | Include |  |
| VAN ESCH, S. C. M., NIJKAMP, M. D., CORNEL, M. C. & SNOEK, F. J. 2012. Patients' intentions to inform relatives about Type 2 diabetes risk: the role of worry in the process of family risk disclosure. Diabetic medicine : a journal of the British Diabetic Association, 29, e461-7. | Ovid MEDLINE | Yes | No | Yes | Include |  |
| VAN ESCH, S. C. M., NIJKAMP, M. D., CORNEL, M. C. & SNOEK, F. J. 2014. Illness representations of type 2 diabetes patients are associated with perceptions of diabetes threat in relatives. Journal of health psychology, 19, 358-68. | Ovid MEDLINE | Yes | No | Yes | Include |  |
| VAN KORLAAR, I. M., VOSSEN, C. Y., ROSENDAAL, F. R., BOVILL, E. G., NAUD, S., CAMERON, L. D. & KAPTEIN, A. A. 2005. Attitudes toward genetic testing for thrombophilia in asymptomatic members of a large family with heritable protein C deficiency. Journal of thrombosis and haemostasis : JTH, 3, 2437-44. | Ovid MEDLINE | Yes | No | No | Exclude | Focuses on thrombophilia |
| VAN MAARLE, M. C., STOUTHARD, M. E. A. & BONSEL, G. J. 2003. Risk perception of participants in a family-based genetic screening program on familial hypercholesterolemia. American journal of medical genetics. Part A, 116A, 136-43. | Ovid MEDLINE | No | No | No | Exclude | Focuses on familial hypercholesterolaemia |
| VASSY, J. L., O'BRIEN, K. E., WAXLER, J. L., PARK, E. R., DELAHANTY, L. M., FLOREZ, J. C., MEIGS, J. B. & GRANT, R. W. 2012. Impact of Literacy and Numeracy on Motivation for Behavior Change After Diabetes Genetic Risk Testing. MEDICAL DECISION MAKING, 32, 606-615. | Web of Science | Yes | No | Yes | Include |  |
| VAUGHN, A. E., HALES, D. P., NESHTERUK, C. D. & WARD, D. S. 2019. HomeSTEAD's physical activity and screen media practices and beliefs survey: Instrument development and integrated conceptual model. PLOS ONE, 14. | Web of Science | No | No | No | Exclude | Focuses on instrument development |
| VOILS, C. I., COFFMAN, C. J., GRUBBER, J. M., EDELMAN, D., SADEGHPOUR, A., MACIEJEWSKI, M. L., BOLTON, J., CHO, A., GINSBURG, G. S. & YANCY, W. S., JR. 2015. Does Type 2 Diabetes Genetic Testing and Counseling Reduce Modifiable Risk Factors? A Randomized Controlled Trial of Veterans. Journal of general internal medicine, 30, 1591-8. | Ovid MEDLINE | Yes | No | Yes | Include |  |
| VORDERSTRASSE, A. A., GINSBURG, G. S., KRAUS, W. E., MALDONADO, M. C. J. & WOLEVER, R. Q. 2013. Health coaching and genomics-potential avenues to elicit behavior change in those at risk for chronic disease: Protocol for personalized medicine effectiveness study in air force primary care. Global Advances In Health and Medicine, 2, 26-38. | EMBASE | Yes | No | Yes | Exclude | Protocol – no outcomes |
| VORNANEN, M., AKTAN-COLLAN, K., HALLOWELL, N., KONTTINEN, H. & HAUKKALA, A. 2019. Lay Perspectives on Receiving Different Types of Genomic Secondary Findings: a Qualitative Vignette Study. Journal of genetic counseling, 28, 343-354. | Ovid MEDLINE | Yes | Yes | Yes | Include | Genomic secondary findings |
| VORNANEN, M., KONTTINEN, H., KAARIAINEN, H., MANNISTO, S., SALOMAA, V., PEROLA, M. & HAUKKALA, A. 2016. Family history and perceived risk of diabetes, cardiovascular disease, cancer, and depression. Preventive medicine, 90, 177-83. | Ovid MEDLINE | Yes | No | Yes | Include |  |
| VOSSEN, C. Y., ROSENDAAL, F. R., BOVILL, E. G., NAUD, S., CAMERON, L. D., KAPTEIN, A. A. & VAN KORLAAR, I. M. 2005. Attitudes toward genetic testing for thrombophilia in asymptomatic members of a large family with heritable protein C deficiency. Journal of Thrombosis and Haemostasis, 3, 2437-2444. | EMBASE | Yes | No | No | Exclude |  |
| VU, A. V., TURK, N., DURU, O. K., MANGIONE, C., PANCHAL, H., AMAYA, S. A., NORRIS, K. C. & MOIN, T. 2020. The impact of type 2 diabetes mellitus risk perception on adoption of preventive strategies in women with a history of gestational diabetes. Diabetes, 69. | EMBASE | Yes | No | Yes | Include |  |
| WALTER, F. M. & EMERY, J. 2005. 'Coming down the line'-- patients' understanding of their family history of common chronic disease. Annals of family medicine, 3, 405-14. | Ovid MEDLINE | Yes | Yes | Yes | Include |  |
| WALTER, F. M., EMERY, J., BRAITHWAITE, D. & MARTEAU, T. M. 2004. Lay understanding of familial risk of common chronic diseases: A systematic review and synthesis of qualitative research. Annals of Family Medicine, 2, 583-594. | EMBASE | Yes | Yes | Yes | Include |  |
| WANG, C., GONZALEZ, R. & MERAJVER, S. D. 2004. Assessment of genetic testing and related counseling services: current research and future directions. SOCIAL SCIENCE & MEDICINE, 58, 1427-1442. | Web of Science | No | Yes | Yes | Include |  |
| WANG, C., SEN, A., RUFFIN, M. T., NEASE, D. E., GRAMLING, R., ACHESON, L. S., O'NEILL, S. M. & RUBINSTEIN, W. S. 2012. Family history assessment: Impact on disease risk perceptions. American Journal of Preventive Medicine, 43, 392-398. | EMBASE | No | No | Yes | Include |  |
| WATERS, E. A., ACKERMAN, N. & WHEELER, C. S. 2019. Cognitive and Affective Responses to Mass-media Based Genetic Risk Information in a Socio-demographically Diverse Sample of Smokers. JOURNAL OF HEALTH COMMUNICATION, 24, 700-710. | Web of Science | Yes | No | Yes | Include | Focuses on public views of condition |
| WAXLER, J. L., O'BRIEN, K. E., DELAHANTY, L. M., MEIGS, J. B., FLOREZ, J. C., PARK, E. R., POBER, B. R. & GRANT, R. W. 2012. Genetic counseling as a tool for type 2 diabetes prevention: a genetic counseling framework for common polygenetic disorders. Journal of genetic counseling, 21, 684-91. | Ovid MEDLINE | No | No | Yes | Include |  |
| WEBB, M. S. & CAREY, M. P. 2009. Psychosocial factors associated with weight control expectancies in treatment-seeking African American smokers. Journal of the National Medical Association, 101, 793-799. | PsycINFO | Yes | No | Yes | Exclude | Full text unavailable |
| WEBBER, C. J., O'HEA, E. C., ABAR, B., BOCK, B. & BOUDREAUX, E. D. 2020. Ecological momentary assessment and first smoking cessation lapse after an acute cardiac event: A pilot study. Journal of health psychology, 25, 1076-1081. | Ovid MEDLINE | No | No | No | Exclude | Focuses on ecological momentary assessment of smoking lapse |
| WEINER, K. & DURRINGTON, P. N. 2008. Patients' understandings and experiences of familial hypercholesterolemia. Community genetics, 11, 273-82. | Ovid MEDLINE | Yes | Yes | No | Exclude | Focuses on familial hypercholesterolaemia |
| WENZEL, L. & GLANZ, K. 2004. Behavioral Aspects of Genetic Risk for Disease: Cancer Genetics as a Prototype for Complex Issues in Health Psychology. In: BOLL, T. J., FRANK, R. G., BAUM, A. & WALLANDER, J. L. (eds.) Handbook of clinical health psychology: Volume 3. Models and perspectives in health psychology. Washington, DC: American Psychological Association. | PsycINFO | No | Yes | Yes | Include |  |
| WESSEL, J., GUPTA, J. & DE GROOT, M. 2013. Factors motivating individuals to consider genetic testing for type 2 diabetes risk prediction. Diabetes, 62, A196. | EMBASE | Yes | No | Yes | Exclude | Full text unavailable |
| WESSEL, J., GUPTA, J. & DE GROOT, M. 2016. Factors Motivating Individuals to Consider Genetic Testing for Type 2 Diabetes Risk Prediction. PloS one, 11, e0147071. | Ovid MEDLINE | No | No | Yes | Include |  |
| WHITFORD, D. L. & AL-SABBAGH, M. 2010. Cultural variations in attitudes towards family risk of diabetes. Diabetes Research and Clinical Practice, 90, 173-181. | Scopus | Yes | No | Yes | Exclude | Full text unavailable |
| WIJDENES, M., HENNEMAN, L., QURESHI, N., KOSTENSE, P. J., CORNEL, M. C. & TIMMERMANS, D. R. M. 2013. Using web-based familial risk information for diabetes prevention: a randomized controlled trial. BMC public health, 13, 485. | Ovid MEDLINE | No | No | Yes | Include |  |
| WIJDENES-PIJL, M., DONDORP, W. J., TIMMERMANS, D. R. M., CORNEL, M. C. & HENNEMAN, L. 2011. Lay perceptions of predictive testing for diabetes based on DNA test results versus family history assessment: a focus group study. BMC PUBLIC HEALTH, 11. | Web of Science | No | Yes | Yes | Include |  |
| WILSON, C. J., DE LA HAYE, K., COVENEY, J., HUGHES, D. L., HUTCHINSON, A., MILLER, C., PRICHARD, I., WARD, P. & KOEHLY, L. M. 2016. Protocol for a randomized controlled trial testing the impact of feedback on familial risk of chronic diseases on family-level intentions to participate in preventive lifestyle behaviors. BMC PUBLIC HEALTH, 16. | Web of Science | No | No | Yes | Exclude | Protocol – no outcomes |
| WRIGHT, A. J., SUTTON, S. R., HANKINS, M., WHITWELL, S. C. L., MACFARLANE, A. & MARTEAU, T. M. 2012. Why does genetic causal information alter perceived treatment effectiveness? An analogue study. British journal of health psychology, 17, 294-313. | Ovid MEDLINE | No | No | Yes | Include |  |
| WRIGHT, C. E., HARVIE, M., HOWELL, A., EVANS, D. G., HULBERT-WILLIAMS, N. & DONNELLY, L. S. 2015. Beliefs about weight and breast cancer: an interview study with high risk women following a 12 month weight loss intervention. HEREDITARY CANCER IN CLINICAL PRACTICE, 13. | Web of Science | No | Yes | No | Exclude | Focuses on cancer |
| WU, R. R., HIMMEL, T., MYERS, R. A., HAUSER, E., VORDERSTRASSE, A., GINSBURG, G. & ORLANDO, L. A. 2015. Effect of family history and genetic risk counselling for type 2 diabetes on perceptions of risk and control: secondary analysis of a randomized controlled trial. Journal of general internal medicine, 30, S55‐. | Cochrane Central Register of Controlled Trials (CENTRAL) | No | No | Yes | Exclude | Full text unavailable |
| WU, R. R., MYERS, R. A., HAUSER, E. R., VORDERSTRASSE, A., CHO, A., GINSBURG, G. S. & ORLANDO, L. A. 2017. Impact of Genetic Testing and Family Health History Based Risk Counseling on Behavior Change and Cognitive Precursors for Type 2 Diabetes. Journal of genetic counseling, 26, 133-140. | Ovid MEDLINE | No | No | Yes | Include |  |
| XU, E. J., BOYER, L. P. L., JAAR, B. G., EPHRAIM, P. L., GIMENEZ, L., CHENG, A., CHRISPIN, J., WEIR, M. R., RAJ, D., GUALLAR, E. & SHAFI, T. 2021. Patients’ and family members’ perspectives on arrhythmias and sudden death in dialysis: the HeartLink focus groups pilot study. BMC Nephrology, 22. | EMBASE | Yes | Yes | No | Exclude |  |
| YANG, K., BANIAK, L. M., IMES, C. C., CHOI, J. & CHASENS, E. R. 2018. Perceived Versus Actual Risk of Type 2 Diabetes by Race and Ethnicity. DIABETES EDUCATOR, 44, 269-277. | Web of Science | Yes | No | Yes | Include |  |
| YOUNG-HYMAN, D., HERMAN, L. J., SCOTT, D. L. & SCHLUNDT, D. G. 2000. Care giver perception of children's obesity-related health risk: A study of African American families. OBESITY RESEARCH, 8, 241-248. | Web of Science | Yes | No | Yes | Include |  |
| ZLOT, A. I., BLAND, M. P., SILVEY, K., EPSTEIN, B., MIELKE, B. & LEMAN, R. F. 2009. Influence of Family History of Diabetes on Health Care Provider Practice and Patient Behavior Among Nondiabetic Oregonians. PREVENTING CHRONIC DISEASE, 6. | Web of Science | Yes | No | Yes | Include |  |
| ZLOT, A. I., VALDEZ, R., HAN, Y., SILVEY, K. & LEMAN, R. F. 2010. Influence of family history of cardiovascular disease on clinicians' preventive recommendations and subsequent adherence of patients without cardiovascular disease. Public health genomics, 13, 457-66. | Ovid MEDLINE | No | No | Yes | Include |  |
