## Supplementary figures and images for "Advancing the communication of genetic risk for cardiometabolic diseases: A critical interpretive synthesis"

### Additional file 3

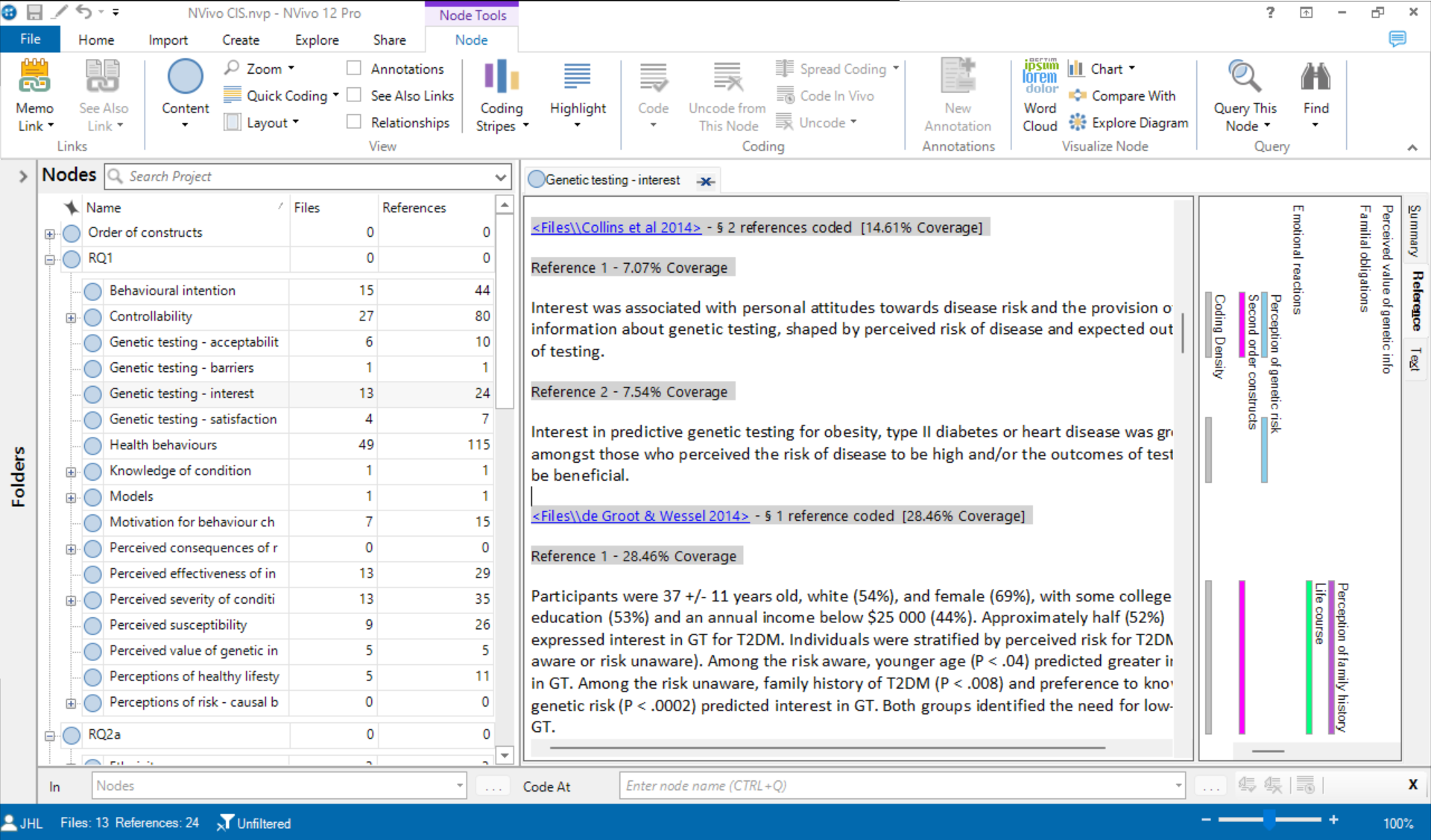
