## Additional file 4 for "Advancing the communication of genetic risk for cardiometabolic diseases: A critical interpretive synthesis"

| Reference | Source | Geographical region | Type of study | Population | Aims and objectives |
| --- | --- | --- | --- | --- | --- |
| ACHESON, L. S., WANG, C., ZYZANSKI, S. J., LYNN, A., RUFFIN, M. T. T., GRAMLING, R., RUBINSTEIN, W. S., O’NEILL, S. M., NEASE, D. E., JR. & FAMILY HEALTHWARE IMPACT TRIAL GROUP 2010. Family history and perceptions about risk and prevention for chronic diseases in primary care: a report from the family healthware impact trial. *Genetics in medicine : official journal of the American College of Medical Genetics,* 12, 212-8. | Ovid MEDLINE | Northern America (US) | Quantitative (cross-sectional analysis of the cluster-randomised, controlled Family Healthware Impact Trial, or FHITr) | 2,330 patients recruited to FHITr from primary care practices (aged 35 to 65 years) | To determine whether family medical history as a risk factor for six common diseases (CHD, stroke, diabetes, and breast, ovarian, and colon cancers) is related to patients’ perceptions of risk, worry, and control over getting these diseases. |
| AL SHAFAEE, M. A., AL-SHUKAILI, S., RIZVI, S. G. A., AL FARSI, Y., KHAN, M. A., GANGULY, S. S., AFIFI, M. & AL ADAWI, S. 2008. Knowledge and perceptions of diabetes in a semi-urban Omani population. *BMC PUBLIC HEALTH,* 8. | Web of Science | Middle East (Oman) | Quantitative (interview questionnaires) | 563 adult residents in two semi-urban localities in the Sultanate of Oman (aged 20 years and above) | To evaluate the knowledge and perception of diabetes in a sample of the Omani general population, and the associations between the elements of knowledge and perception, and socio-demographic factors. |
| ALKERWI, A., PAGNY, S., LAIR, M. L., DELAGARDELLE, C. & BEISSEL, J. 2013. Level of Unawareness and Management of Diabetes, Hypertension, and Dyslipidemia among Adults in Luxembourg: Findings from ORISCAV-LUX Study. *PLOS ONE,* 8. | Web of Science | Europe (Luxembourg) | Quantitative (cross-sectional analysis of the ORISCAV-LUX survey, a population-based cardiovascular risk factors survey) | 1,432 adult participants recruited for ORISCAV-LUX, weighted to produce nationally representative estimates of the total population residing in Luxembourg (aged 18 to 69 years) | To 1) assess the population level of unawareness diabetes, hypertension, and dyslipidaemia; 2) identify the potential determinants of lack of awareness; 3) evaluate the level of management for the three pathologies and 4) provide information about the 10-year risk prediction of CHD among the unaware (untreated) groups. |
| AMIREAULT, S., GODIN, G., VOHL, M. C. & PERUSSE, L. 2008. Moderators of the intention-behaviour and perceived behavioural control-behaviour relationships for leisure-time physical activity. *INTERNATIONAL JOURNAL OF BEHAVIORAL NUTRITION AND PHYSICAL ACTIVITY,* 5. | Web of Science | Northern America (Canada) | Quantitative (cross-sectional survey, conducted as part of a larger study on genetic susceptibility to obesity) | 300 volunteers recruited in the Quebec City metropolitan area via local newspapers and radio advertisements between May 2004 to March 2007 (aged 18 to 55 years) | To identify psychosocial, socio-demographic, as well as biological moderators of the intention-behaviour and perceived behavioural control-behaviour relationships for leisure-time physical activity, using the Theory of Planned Behaviour (TPB). |
| AMUTA, A. O. 2016. *Diabetes family health history among college students.* 77, ProQuest Information & Learning. | PsycINFO | Northern America (US) | Theses and dissertations | 909 undergraduate students enrolled full time or part-time in four colleges/universities across Texas (aged 18 years and above) | To 1) determine what the extant scientific literature reports on the association between T2D family history status and T2D related-preventive behaviours; 2) examine differences in T2D knowledge (behavioural and genetic) among college students with and without a family history of T2D, and assess the influence of demographic characteristics (age, biological sex, BMI, race and marital status) and 3) use Structural Equation Modelling procedures to assess the relative impact of behavioural intention, attitude, perceptions (risk and severity), family history and demographic factors (age, biological sex, BMI, race and marital status) on T2D related-preventive behaviours among college students. |
| ANDERSSON, P., SJOBERG, R. L., OHRVIK, J. & LEPPERT, J. 2009. The effects of family history and personal experiences of illness on the inclination to change health-related behaviour. *Cent Eur J Public Health,* 17, 3-7. | Other – reference chaining | Europe (Sweden and Poland) | Quantitative (cross-sectional survey) | 2,054 participants recruited from a primary health care screening programme (aged 50) | To examine how a personal experience of illness and a family history of cardiovascular disease (CVD), adjusted for sex, level of education and nationality, affect risk behaviour. |
| ASRIL, N. M., TABUCHI, K., TSUNEMATSU, M., KOBAYASHI, T. & KAKEHASHI, M. 2020. Predicting Healthy Lifestyle Behaviours Among Patients With Type 2 Diabetes in Rural Bali, Indonesia. *Clinical Medicine Insights: Endocrinology and Diabetes,* 13. | EMBASE | Asia (Indonesia) | Quantitative (cross-sectional survey) | 203 patients with T2D recruited from community health centres in the rural areas of Bali province (mean age 54.6 years) | To identify factors explaining the healthy lifestyle behaviours of patients with T2D, using the extended Health Belief Model (HBM). |
| BANERJEE, A. T., MAHAJAN, A., MATHUR-BALENDRA, A., QURESHI, N., TEEKAH, M., YOGARATNAM, S., PRABHAKAR, P., AHMED, S., SHAH, B. R., VELUMMAILUM, R., PRICE, J. A. D., DE SOUZA, R. J. & BAJAJ, H. S. 2022. Impact of the South Asian Adolescent Diabetes Awareness Program (SAADAP) on diabetes knowledge, risk perception and health behaviour. *HEALTH EDUCATION JOURNAL*. | Web of Science | Northern America (Canada) | Quantitative (pilot intervention with a pre-test, post-test design for South Asian youth in Canada with a family history of T2D – the South Asian Adolescent Diabetes Awareness Program, or SAADAP) | 68 adolescents with a family history of T2D, recruited from a clinical-community setting in Canada (aged 13 to 17 years) | To investigate changes in 1) diabetes knowledge and associated risk factors; 2) risk perception and 3) health behaviours among adolescents participating in SAADAP. |
| BASILIO, C. D., KWAN, V. S. Y. & TOWERS, M. J. 2016. Culture and Risk Assessments: Why Latino Americans Perceive Greater Risk for Diabetes. *CULTURAL DIVERSITY & ETHNIC MINORITY PSYCHOLOGY,* 22, 104-113. | Web of Science | Northern America (US) | Quantitative (cross-sectional survey) | 306 undergraduate students in Arizona, self-identifying as either Latino American or European American (mean age 19.26 years) | To 1) examine ethnic group differences in perceived vulnerability to disease between European Americans and Latino Americans; 2) examine potential psychological mechanisms that account for ethnic differences in perceived vulnerability to disease and 3) examine the relationship between ethnic identification and perceived vulnerability to disease among Latino Americans. |
| BATES, B. R., TEMPLETON, A., ACHTER, P. J., HARRIS, T. M. & CONDIT, C. M. 2003. What does “a gene for heart disease” mean? A focus group study of public understandings of genetic risk factors. *American journal of medical genetics. Part A,* 119A, 156-61. | Ovid MEDLINE | Northern America (US) | Qualitative (focus group study) | 108 participants (predominantly African-Americans) recruited from urban, suburban, and rural communities in Georgia between July to October 2001 (mean age 32.6 years) | To access public perceptions of potentially deterministic phrasing of genetic risk factors and establish interpretations of the phrase, “a gene for heart disease.” |
| BEKKE-HANSEN, S., WEINMAN, J., THASTUM, M., THYGESEN, K. & ZACHARIAE, R. 2014. Psycho-social factors are important for the perception of disease in patients with acute coronary disease. *Danish medical journal,* 61, A4885. | Ovid MEDLINE | Europe (Denmark) | Quantitative (cross-sectional study, conducted as part of a larger prospective questionnaire study) | 97 hospitalised patients with acute coronary syndrome (ACS) recruited consecutively from Aarhus University Hospital, Denmark in 2008/09 (aged 18 years and above) | To examine the role of socio-demographic, illness-related and psycho-social factors (Multidimensional Scale of Perceived Social Support, General Self-Efficacy Scale and Life Orientation Test-Revised) in the perceived consequences, controllability and causes (Revised Illness Perception Questionnaire) of illness, among patients with ACS. |
| BENNICH, B. B., MUNCH, L., OVERGAARD, D., KONRADSEN, H., KNOP, F. K., RODER, M., VILSBOLL, T. & EGEROD, I. 2020. Experience of family function, family involvement, and self-management in adult patients with type 2 diabetes: A thematic analysis. *JOURNAL OF ADVANCED NURSING,* 76, 621-631. | Web of Science | Europe (Denmark) | Qualitative (semi-structured interview study) | 20 adult patients with T2D recruited from the outpatient clinic of a university hospital in Denmark (mean age 69 years) | To describe patients’ experience of family function and its importance in diabetes-related self-management. |
| BERGE, J. M., ARIKIAN, A., DOHERTY, W. J. & NEUMARK-SZTAINER, D. 2012. Healthful eating and physical activity in the home environment: Results from multifamily focus groups. *Journal of Nutrition Education and Behavior,* 44, 123-131. | PsycINFO | Northern America (US) | Qualitative (multifamily focus group study) | 103 participants representing 26 family units, primarily black and white low- to middle-income families living in urban Minnesota (aged 8 to 61 years) | To explore perceptions of risk and protective factors for child and adolescent obesity in the home environment. |
| BERNHARDT, B. A., TAMBOR, E. S., FRASER, G., WISSOW, L. S. & GELLER, G. 2003. Parents’ and children’s attitudes toward the enrollment of minors in genetic susceptibility research: implications for informed consent. *American journal of medical genetics. Part A,* 116A, 315-23. | Ovid MEDLINE | Northern America (US) | Qualitative (semi-structured interview study) | 74 participants representing 37 family units (aged 10 years and above) | To assess parents’ and children’s reactions to participation in disease susceptibility research involving genetic testing—and their perceptions of risks and benefits of participating. |
| BLUE, G. M., KASPARIAN, N. A., SHOLLER, G. F., KIRK, E. P. & WINLAW, D. S. 2015. Genetic counselling in parents of children with congenital heart disease significantly improves knowledge about causation and enhances psychosocial functioning. *International journal of cardiology,* 178, 124-30. | Ovid MEDLINE | Oceania (Australia) | Quantitative (cross-sectional survey) | 55 parents of children with CHD undergoing elective cardiac surgery at The Children’s Hospital at Westmead (mean age 34.1 years) | To assess the efficacy of individualised genetic counselling sessions in improving knowledge of causation and psychosocial functioning in parents of children with CHD. |
| BOELDT, D. L., SCHORK, N. J., TOPOL, E. J. & BLOSS, C. S. 2015. Influence of individual differences in disease perception on consumer response to direct-to-consumer genomic testing. *Clinical Genetics,* 87, 225-232. | EMBASE | Northern America (US) | Quantitative (cross-sectional analysis of the Scripps Genomic Health Initiative, a longitudinal cohort study examining the psychological and behavioural impact of direct-to-consumer testing for common diseases) | 2,037 participants recruited from health and technology companies (mean age 46.7 years) | To evaluate consumer perceptions of direct-to-consumer personalised genomic risk assessments and assess the extent to which consumer characteristics may be associated with attitudes toward testing. |
| BOWLES, B. C. 2010. *Genograms as threat appeals: Using the extended parallel process model with familial cardiovascular disease.* 70, ProQuest Information & Learning. | PsycINFO | Northern America (US) | Theses and dissertations | 171 participants with at least one first-degree relative with CVD (aged 18 years and above) | To introduce the health genogram (based on the Extended Parallel Process Model) as a threat appeal in motivating intent to perform health promotion and disease prevention behaviours in participants with a family history of CVD. |
| BROCKMAN, D. G., PETRONIO, L., DRON, J. S., KWON, B. C., VOSBURG, T., NIP, L., TANG, A., O’REILLY, M., LENNON, N., WONG, B., NG, K., HUANG, K. H., FAHED, A. C. & KHERA, A. V. 2021. Design and user experience testing of a polygenic score report: a qualitative study of prospective users. *BMC Med Genomics,* 14, 238. | Other – expert subject knowledge in review team | Northern America (US) | Qualitative (interview study on user experience) | 10 participants recruited from a national online recruitment platform (mean age 50.3 years) | To review the landscape of PRS reporting and describe a generalisable approach for development of a PRS disclosure tool for CAD. |
| BROWN, J. B., HARRIS, S. B., WEBSTER-BOGAERT, S., WETMORE, S., FAULDS, C. & STEWART, M. 2002. The role of patient, physician and systemic factors in the management of type 2 diabetes mellitus. *FAMILY PRACTICE,* 19, 344-349. | Web of Science | Northern America (Canada) | Qualitative (semi-structured focus group interview study) | 30 family physicians participating in a simultaneous quantitative study on the management of patients with T2D | To explore family physicians’ issues and perceptions regarding the barriers to and facilitators of the management of patients with T2D. |
| BRUST-RENCK, P. G., REYNA, V. F., WILHELMS, E. A. & LAZAR, A. N. 2016. A fuzzy-trace theory of judgment and decision-making in health care: Explanation, prediction, and application. *In:* DIEFENBACH, M. A., MILLER-HALEGOUA, S. & BOWEN, D. J. (eds.) *Handbook of health decision science.* New York, NY: Springer Science + Business Media. | PsycINFO | N/A | Commentaries and opinion pieces | N/A | To discuss how an evidence-based theory of human behaviour and decision-making—Fuzzy-Trace Theory (FTT)—can be used to better understand and improve public health and medicine. |
| BUIGUES, C., QUERALT, A., DE VELASCO, J. A., SALVADOR-SANZ, A., JENNINGS, C., WOOD, D. & TRAPERO, I. 2021. Psycho-Social Factors in Patients with Cardiovascular Disease Attending a Family-Centred Prevention and Rehabilitation Programme: EUROACTION Model in Spain. *LIFE-BASEL,* 11. | Web of Science | Europe (Spain) | Quantitative (analysis of the cluster randomised, controlled trial EUROACTION) | 165 participants in intervention group and 210 participants in usual care (mean age 59.87 and 64.53 years respectively) | To highlight the effects of an interdisciplinary, family-centred cardiovascular prevention and rehabilitation programme on modifying illness perceptions to achieve better outcomes and therefore improve the state of anxiety and depression of patients, as well as improving not only their lifestyle but also that of their partners. |
| CAMERON, L. D., SHERMAN, K. A., MARTEAU, T. M. & BROWN, P. M. 2009. Impact of genetic risk information and type of disease on perceived risk, anticipated affect, and expected consequences of genetic tests. *Health psychology : official journal of the Division of Health Psychology, American Psychological Association,* 28, 307-16. | Ovid MEDLINE | Europe and Oceania (UK, Australia, New Zealand) | Quantitative (randomised online experiment) | 752 participants recruited through university networks (mean age 26 years) | To assess how increments in absolute risk of disease (diabetes, heart disease, colon cancer, or lung cancer) influence risk perceptions, interest, and expected consequences of genetic tests for diseases of varying severity. |
| CERSOSIMO, E. & MUSI, N. 2011. Improving Treatment in Hispanic/Latino Patients. *AMERICAN JOURNAL OF MEDICINE,* 124, S16-S21. | Web of Science | N/A | Commentaries and opinion pieces | N/A | To propose policies that can reduce disparities between Hispanics/Latinos and non-Hispanic whites regarding health insurance coverage and access to healthcare. |
| CHARBONNEAU, J., NICOL, D., CHALMERS, D., KATO, K., YAMAMOTO, N., WALSHE, J. & CRITCHLEY, C. 2020. Public reactions to direct-to-consumer genetic health tests: A comparison across the US, UK, Japan and Australia. *European Journal of Human Genetics,* 28, 339-348. | EMBASE | Northern America, Europe, Asia and Oceania (US, UK, Japan and Australia) | Quantitative (cross-sectional study) | 4,032 participants sourced by Qualtrics in four countries (mean age 46.51 years) | To investigate general public views of direct-to-consumer genetic testing across four countries, each at different stages of market development. |
| CISLAK, A., SAFRON, M., PRATT, M., GASPAR, T. & LUSZCZYNSKA, A. 2012. Family-related predictors of body weight and weight-related behaviours among children and adolescents: a systematic umbrella review. *CHILD CARE HEALTH AND DEVELOPMENT,* 38, 321-331. | Web of Science | N/A | Systematic reviews | N/A | To analyse the relationships between family variables and child/adolescent body weight, diet and physical activity. |
| CITARELLA, A., KIELER, H., SUNDSTROM, A., LINDER, M., WETTERMARK, B., BERGLIND, I. A. & ANDERSEN, M. 2014. Family history of cardiovascular disease and influence on statin therapy persistence. *EUROPEAN JOURNAL OF CLINICAL PHARMACOLOGY,* 70, 701-707. | Web of Science | Europe (Sweden) | Quantitative (population-based cohort study) | 86,002 patients identified in the Swedish register on dispensed drugs, hospitalisation and cause of death and the Multi-generation Register (aged 20 to 72 years) | To investigate whether family history of CVD influences the discontinuation of statin treatment. |
| CLAASSEN, L., HENNEMAN, L., DE VET, R., KNOL, D., MARTEAU, T. & TIMMERMANS, D. 2010. Fatalistic responses to different types of genetic risk information: Exploring the role of self-malleability. *Psychology & Health,* 25, 183-196. | PsycINFO | Europe (the Netherlands) | Quantitative (cross-sectional study) | 94 students from The Vrije Universiteit Amsterdam (mean age 21.5 years) | To 1) construct a scale that captures the extent to which people view themselves as able to change self-attributes, the Malleability of Self questionnaire and 2) examine responses to scenario vignettes describing different types of health risk information and to assess the predictive validity of Malleability of Self in explaining these responses. |
| CLAASSEN, L., HENNEMAN, L., KINDT, I., MARTEAU, T. M. & TIMMERMANS, D. R. M. 2010. Perceived risk and representations of cardiovascular disease and preventive behaviour in people diagnosed with familial hypercholesterolemia: a cross-sectional questionnaire study. *Journal of health psychology,* 15, 33-43. | Ovid MEDLINE | Europe (the Netherlands) | Quantitative (cross-sectional study) | 81 participants screen positive for Familial Hypercholesterolemia in nationwide family cascade screening programme | To assess the perceived risk and representations of CVD and preventive behaviours of people diagnosed with Familial Hypercholesterolemia by DNA testing. |
| CLAASSEN, L., HENNEMAN, L., VAN DER WEIJDEN, T., MARTEAU, T. M. & TIMMERMANS, D. R. M. 2012. Being at risk for cardiovascular disease: perceptions and preventive behavior in people with and without a known genetic predisposition. *Psychology, health & medicine,* 17, 511-21. | Ovid MEDLINE | Europe (the Netherlands) | Quantitative (cross-sectional study) | 51 participants with a genetic predisposition to CVD and 49 participants without a genetic predisposition to CVD (mean age 54 and 55 years respectively) | To compare and explain differences in perceptions of CVD risk and preventive behaviours in people with and without a known genetic predisposition to CVD. |
| COLLINS, J., RYAN, L. & TRUBY, H. 2014. A systematic review of the factors associated with interest in predictive genetic testing for obesity, type II diabetes and heart disease. *Journal of human nutrition and dietetics : the official journal of the British Dietetic Association,* 27, 479-88. | Ovid MEDLINE | N/A | Systematic reviews | N/A | To identify the factors associated with an interest in having predictive genetic testing for obesity, type II diabetes and heart disease amongst unaffected adults. |
| COLLINS, R. E., WRIGHT, A. J. & MARTEAU, T. M. 2011. Impact of communicating personalized genetic risk information on perceived control over the risk: a systematic review. *Genet Med,* 13, 273-7. | Other – expert subject knowledge in review team | N/A | Systematic reviews | N/A | To assess the strength of evidence surrounding the feedback of personalised genetic risk information leading to fatalism, i.e., a lack of perceived control over risk. |
| CULLEN, K. W. & BUZEK, B. B. 2009. Knowledge about type 2 diabetes risk and prevention of African-American and Hispanic adults and adolescents with family history of type 2 diabetes. *The Diabetes educator,* 35, 836-42. | Ovid MEDLINE | Northern America (US) | Qualitative (interview study) | 39 parents and 21 ninth and tenth grade adolescents in Houston, Texas reporting a family history of T2D | To assess T2D knowledge, perceptions, risk factor awareness, and prevention practices among African American and Hispanic families with a history of diabetes. |
| CUNNINGHAM, A. T., GENTSCH, A. T., DOTY, A. M. B., MILLS, G., LANOUE, M., CARR, B. G., HOLLANDER, J. E. & RISING, K. L. 2020. “I had no other choice but to catch it too”: the roles of family history and experiences with diabetes in illness representations. *BMC endocrine disorders,* 20, 95. | Ovid MEDLINE | Northern America (US) | Qualitative (secondary analysis of interview data) | 89 patients with T1D or T2D seeking care in an urban health system (mean age 54.6 years) | To explore the perceptions of diabetes family history and experiences on the illness representations of individuals with diabetes. |
| DAACK-HIRSCH, S., SCHUMACHER, A. C., SHAH, L. & CAMPO, S. 2019. Type 2 diabetes familial risk personalization process profiles: Implications for patient-provider communication. *Research in nursing & health,* 42, 369-381. | Ovid MEDLINE | Northern America (US) | Mixed-methods (secondary analysis of survey data and interview data) | 109 participants with a positive family history of T2D (mean age 28.8 years) | To identify possible subtypes of the familial risk perception (FRP) model and explore how people with a positive family history for T2D personalise and process their familial risk to form perceptions about their own risk. |
| DAACK-HIRSCH, S., SHAH, L. L. & CADY, A. D. 2018. Mental Models of Cause and Inheritance for Type 2 Diabetes Among Unaffected Individuals Who Have a Positive Family History. *Qualitative health research,* 28, 534-547. | Ovid MEDLINE | Northern America (US) | Qualitative (secondary analysis of interview data) | 111 participants with moderate-to-high familial risk for T2D (mean age 29 years) | To elicit causal and inheritance explanations for T2D, using the FRP model as a framework, in participants with a family history. |
| DAACK-HIRSCH, S., SHAH, L. L., JONES, K., ROCHA, B., DOERR, M., GABITZSCH, E. & MEESE, T. 2020. All things considered, my risk for diabetes is medium: A risk personalization process of familial risk for type 2 diabetes. *Health expectations : an international journal of public participation in health care and health policy,* 23, 169-181. | Ovid MEDLINE | Northern America (US) | Mixed-methods (secondary analysis of survey data and clinical risk assessment) | 111 participants with moderate-to-high familial risk for T2D (mean age 29 years) | To characterize two key concepts, salience and vulnerability, within the FRP model among unaffected individuals, at increased familial risk for T2D. |
| DAMMAN, O. C., BOGAERTS, N. M. M., VAN DEN HAAK, M. J. & TIMMERMANS, D. R. M. 2017. How lay people understand and make sense of personalized disease risk information. *Health Expectations: An International Journal of Public Participation in Health Care & Health Policy,* 20, 973-983. | PsycINFO | Europe (the Netherlands) | Mixed-methods (analysis of eye-tracking data and interview data) | 16 participants without medical history of T2D (aged 45 to 65 years) | To examine how lay people understand the result derived from an online cardiometabolic risk calculator. |
| DAR-NIMROD, I., CHEUNG, B. Y., RUBY, M. B. & HEINE, S. J. 2014. Can merely learning about obesity genes affect eating behavior? *Appetite,* 81, 269-76. | Ovid MEDLINE | Northern America (Canada) | Quantitative (experimental study) | 143 undergraduate students (mean age 20.5 years) | To assess the potential behavioural implications of a perceived genetic aetiology for obesity, using the TPB as framework. |
| DAVIES, L. E. & THIRLAWAY, K. 2013. The influence of genetic explanations of type 2 diabetes on patients’ attitudes to prevention, treatment and personal responsibility for health. *Public health genomics,* 16, 199-207. | Ovid MEDLINE | Europe (UK) | Quantitative (experimental study) | 200 participants recruited from a primary care setting (mean age 59.2 years) | To investigate the role of perceived aetiology in the beliefs and attitudes that may influence personal responsibility and perceived efficacy of preventative behaviours and treatment of T2D. |
| DE GROOT, M. & WESSEL, J. 2014. Genetic Testing and Type 2 Diabetes Risk Awareness. *The Diabetes educator,* 40, 427-433. | Ovid MEDLINE | Northern America (US) | Quantitative (cross-sectional survey) | 265 participants representing diverse groups in Indiana (mean age 37 years) | To examine the motivational, attitudinal, and behavioural predictors of interest in genetic testing in those with and without awareness of their risk for T2D. |
| DESALVO, K. B., GREGG, J., KLEINPETER, M., PEDERSEN, B. R., STEPTER, A. & PEABODY, J. 2005. Cardiac risk underestimation in urban, black women. *JOURNAL OF GENERAL INTERNAL MEDICINE,* 20, 1127-1131. | Web of Science | Northern America (US) | Mixed-methods (interview study and clinical risk assessment) | 128 black women recruited from an urban continuity clinic in metropolitan  New Orleans (mean age 56 years) | To investigate the personal characteristics associated with underestimating cardiovascular disease in black women. |
| DIGNAN, M. B., YOUNG, L. D., CROUSE, J. R. & KING, J. M. 1995. Factors associated with participation in a preventive cardiology service by patients with coronary heart disease. *Southern medical journal,* 88, 1057-61. | Ovid MEDLINE | Northern America (US) | Quantitative (cross-sectional survey) | 62 patients with CHD (mean age 51.1 years) | To explore determinants of attending a preventive cardiology consultation service, using the HBM as framework. |
| DILORENZO, T. A., SCHNUR, J., MONTGOMERY, G. H., ERBLICH, J., WINKEL, G. & BOVBJERG, D. H. 2006. A model of disease-specific worry in heritable disease: the influence of family history, perceived risk and worry about other illnesses. *Journal of behavioral medicine,* 29, 37-49. | Ovid MEDLINE | Northern America (US) | Quantitative (cross-sectional survey) | 434 participants recruited near the entrance to the cafeteria of a large urban medical centre (mean age 37.9 years) | To examine a theoretical model of disease-specific worry which includes family history of disease, disease-specific perceived risk, perceived risk for other diseases, and worry about other diseases using structural equation modelling. |
| DONG, Y. & BRANSCUM, P. 2019. What Motivates Individuals to Get Obesity Related Direct-To-Consumer Genetic Tests? A Reasoned Action Approach. *AMERICAN JOURNAL OF HEALTH EDUCATION,* 50, 356-365. | Web of Science | Northern America (US) | Quantitative (cross-sectional survey) | 288 undergraduate and postgraduate students (mean age 24.57 years) | To 1) better understand what motivates individuals to take an obesity-related direct-to-consumer genetic test, using the Reasoned Action Approach as a theoretical framework and 2) evaluate differences in the behavioural intentions and other behavioural antecedents based on participants’ awareness of genetic testing. |
| DORMAN, J. S., VALDEZ, R., LIU, T., WANG, C., RUBINSTEIN, W. S., O’NEILL, S. M., ACHESON, L. S., RUFFIN, M. T. T. & KHOURY, M. J. 2012. Health beliefs among individuals at increased familial risk for type 2 diabetes: implications for prevention. *Diabetes research and clinical practice,* 96, 156-62. | Ovid MEDLINE | Northern America (US) | Quantitative (cross-sectional analysis of FHITr) | 2,081 patients recruited to FHITr from primary care practices (mean age 50.82 years) | To evaluate perceived risk, control, worry, and severity about diabetes CHD and stroke among individuals at increased familial risk of diabetes. |
| DRAPKIN, R. G., WING, R. R. & SHIFFMAN, S. 1995. Responses to hypothetical high risk situations: Do they predict weight loss in a behavioral treatment program or the context of dietary lapses? *Health Psychology,* 14, 427-434. | PsycINFO | Northern America (US) | Quantitative (experimental study) | 93 participants with T2D (mean age 52 years) | To determine whether baseline performance on a hypothetical high risk task would predict subsequent weight loss, the type of situations in which  participants would actually lapse, or both. |
| EDELSTEIN, J. & LINN, M. W. 1985. The influence of the family on control of diabetes. *Social Science & Medicine,* 21, 541-544. | PsycINFO | Northern America (US) | Quantitative (cross-sectional survey) | 97 male patients with diabetes (mean age 55 years) | To determine the role of family environment in the metabolic control of adult men with diabetes. |
| ERBLICH, J., BOVBJERG, D. H., NORMAN, C., VALDIMARSDOTTIR, H. B. & MONTGOMERY, G. H. 2000. It won’t happen to me: Lower perception of heart disease risk among women with family histories of breast cancer. *PREVENTIVE MEDICINE,* 31, 714-721. | Web of Science | Northern America (US) | Quantitative (cross-sectional survey) | 177 healthy women with and without a family history of breast cancer (mean age 41.7 years) | To examine the possibility that women with family histories of breast cancer may be particularly susceptible to overestimating their risks of breast cancer while minimizing their risks of CVD. |
| ETELSON, D., BRAND, D. A., PATRICK, P. A. & SHIRALI, A. 2003. Childhood obesity: Do parents recognize this health risk? *OBESITY RESEARCH,* 11, 1362-1368. | Web of Science | Northern America (US) | Quantitative (cross-sectional survey) | 83 parents of children aged between 4 to 8 years | To examine parents’ understanding of excess weight as a health risk, knowledge of healthy eating habits, and recognition of obesity in their children. |
| FARMER, A. J., LEVY, J. C. & TURNER, R. C. 1999. Knowledge of risk of developing diabetes mellitus among siblings of Type 2 diabetic patients. *Diabetic medicine,* 16, 233‐237. | Cochrane Central Register of Controlled Trials (CENTRAL) | Europe (UK) | Quantitative (cross-sectional survey) | 481 participants identified through a progressive search of disease registers in all GPs in the areas of the Oxford and Northampton Health Authorities (median age 60 years) | To investigate the extent to which siblings of patients with T2D perceived themselves likely to develop T2D when offered screening tests. |
| FERRANTI, E. P. 2014. *Dietary quality and cardiometabolic risk after gestational diabetes.* 74, ProQuest Information & Learning. | PsycINFO | Northern America (US) | Quantitative (cross-sectional survey) | 75 women with a history of gestational diabetes (mean age 35.5 years) | To examine individual, family, and social-level influences of diet quality in women with a history of gestational diabetes—guided by a synthesis of the HBM and the ecological framework for eating behaviour. |
| FISHER, E., ACHILLES, S. & TÖNNIES, H. 2014. Predictive genetic testing, risk communication, and risk perception: An international expert meeting in Berlin, Germany. *Journal of Community Genetics,* 5, 1-5. | EMBASE | N/A | Conference papers and proceedings | N/A | To discuss 1) the current status of risk prediction models for common diseases (breast cancer, T2D) and the evidence-based evaluation of genetic tests, in particular, pre-symptomatic or probabilistic genetic tests for health care purposes; 2) the communication of genetic risks to, and perception of genetic risks by healthy persons and 3) the psychological and motivational impacts on persons who underwent genetic testing. |
| FOLLING, I. S., SOLBJOR, M., MIDTHJELL, K., KULSENG, B. & HELVIK, A.-S. 2016. Exploring lifestyle and risk in preventing type 2 diabetes-a nested qualitative study of older participants in a lifestyle intervention program (VEND-RISK). *BMC public health,* 16, 876. | Ovid MEDLINE | Europe (Norway and Finland) | Qualitative (secondary analysis of semi-structured interviews) | 26 participants previously recruited onto the Nord-Trøndelag Health Study 3 (HUNT3), the HUNT DE-PLAN Study and the VEND-RISK Study (mean age 68 years) | To explore how older adults perceived their own lifestyle and being at increased risk for T2D while they participated in a lifestyle intervention programme. |
| FRENCH, D. P., BANAFA, R., WILLIAMS, S., TAYLOR, C. & BROWN, L. J. E. 2021. How Does the Understanding, Experience, and Enactment of Self-Regulation Behaviour Change Techniques Vary with Age? A Thematic Analysis. *Appl Psychol Health Well Being,* 13, 239-260. | Other – expert subject knowledge in review team | Europe (UK) | Qualitative (semi-structured interviews study) | 12 participants involved in a feasibility study of a walking intervention delivered by nurses or healthcare assistants in seven GP surgeries (mean age 58 years) | To investigate participants’ understanding, experiences and enactment of self-regulatory behaviour change techniques, and to consider how these differ according to age. |
| FRICH, J. C., OSE, L., MALTERUD, K. & FUGELLI, P. 2006. Perceived Vulnerability to Heart Disease in Patients with Familial Hypercholesterolemia: A Qualitative Interview Study. *Annals of Family Medicine,* 4, 198-204. | PsycINFO | Europe (Norway) | Qualitative (semi-structured interview study) | 40 patients with familial hypercholesterolemia recruited through a lipid clinic (mean age 31 years) | To explore how patients with a diagnosis of heterozygous familial hypercholesterolemia understand and perceive their vulnerability to CHD. |
| FRIEDRICH, O., KUNSCHITZ, E., PONGRATZ, L., WIELÄNDER, S., SCHÖPPL, C. & SIPÖTZ, J. 2021. Classification of illness attributions in patients with coronary artery disease. *Psychology & health*, 1-16. | EMBASE | Europe (Vienna) | Quantitative (cross-sectional survey) | 459 patients with angiographically verified CAD (median age 66 years) | To examine patient-reported causal attributions in patients with CAD and classify them according to attribution theory. |
| FRIJLING, B. D., LOBO, C. M., KEUS, I. M., JENKS, K. M., AKKERMANS, R. P., HULSCHER, M. E. J. L., PRINS, A., VAN DER WOUDEN, J. C. & GROL, R. P. T. M. 2004. Perceptions of cardiovascular risk among patients with hypertension or diabetes. *Patient education and counseling,* 52, 47-53. | Ovid MEDLINE | Europe (the Netherlands) | Quantitative (survey embedded in trial) | 1,557 patients with hypertension or diabetes but no known atherosclerotic disease (mean age 62.5 years) | To examine risk perceptions among patients at moderate to high cardiovascular risk. |
| FROSCH, D. L., MELLO, P. & LERMAN, C. 2005. Behavioral consequences of testing for obesity risk. *Cancer epidemiology, biomarkers & prevention : a publication of the American Association for Cancer Research, cosponsored by the American Society of Preventive Oncology,* 14, 1485-9. | Ovid MEDLINE | Northern America (US) | Quantitative (experimental study) | 249 undergraduate students from the University of Pennsylvania (mean age 20.5 years) | To examine potential behavioural consequences of genetic testing for susceptibility to obesity. |
| GALLAGHER, P., KING, H. A., HAGA, S. B., ORLANDO, L. A., JOY, S. V., TRUJILLO, G. M., SCOTT, W. M., BEMBE, M., CREIGHTON, D. L., CHO, A. H., GINSBURG, G. S. & VORDERSTRASSE, A. 2015. Patient beliefs and behaviors about genomic risk for type 2 diabetes: Implications for prevention. *Journal of Health Communication,* 20, 728-735. | PsycINFO | Northern America (US) | Quantitative (cross-sectional survey) | 409 patients recruited in the clinical laboratory waiting areas of two primary care outpatient clinics (mean age 50 years) | To examine illness representations in a clinical sample who are at risk for T2D and interested in genetic testing, using the CSM-SR as a framework. |
| GODINO, J. G., VAN SLUIJS, E. M. F., MARTEAU, T. M., SUTTON, S., SHARP, S. J. & GRIFFIN, S. J. 2016. Lifestyle Advice Combined with Personalized Estimates of Genetic or Phenotypic Risk of Type 2 Diabetes, and Objectively Measured Physical Activity: A Randomized Controlled Trial. *PLoS medicine,* 13, e1002185. | Ovid MEDLINE | Europe (UK) | Quantitative (RCT) | 569 healthy middle-aged adults recruited from the Fenland Study, an ongoing population-based, observational study (mean age 48.7 years) | To examine the effect of communicating an estimate of genetic or phenotypic risk of T2D in a parallel-group, open RCT. |
| GOODWIN, C. L. 2018. *A randomized controlled trial of heart disease risk education on delay discounting, perceived disease risk, health behavior, and health behavior intentions among men and women with and without a family history of cardiovascular disease.* 79, ProQuest Information & Learning. | PsycINFO | Northern America (US) | Theses and dissertations | 108 participants recruited from undergraduate students at The Ohio State University, as well as in the community (mean age 23.68 years) | To examine rates of delay discounting among young adults with a family history of early-onset CVD (i.e., adults with higher genetic risk for CVD); and without a family history of CVD. |
| GRABOWSKI, D. & ANDERSEN, T. H. 2020. Barriers to intra-familial prevention of type 2 diabetes: A qualitative study on horizons of significance and social imaginaries. *Chronic Illness,* 16, 119-130. | EMBASE | Europe (Denmark) | Qualitative (problem assessment and ideation workshops with families) | 57 participants with T2D and their relatives (aged 15 years and above) | To explore barriers to prevention in families with at least one adult with T2D. |
| GRAUMAN, Å., VELDWIJK, J., JAMES, S., HANSSON, M. & BYBERG, L. 2021. Good general health and lack of family history influence the underestimation of cardiovascular risk: a cross-sectional study. *European journal of cardiovascular nursing : journal of the Working Group on Cardiovascular Nursing of the European Society of Cardiology*. | EMBASE | Europe (Sweden) | Quantitative (cross-sectional analysis of the Swedish Cardio Pulmonary BioImage Study, or SCAPIS) | 526 participants enrolled in SCAPIS (aged 50 to 64 years) | To investigate whether general health and family history of myocardial infarction are associated with underestimation of perceived cardiovascular risk, and if the participants’ calculated risk modifies that association. |
| GREENHALGH, T., CLINCH, M., AFSAR, N., CHOUDHURY, Y., SUDRA, R., CAMPBELL-RICHARDS, D., CLAYDON, A., HITMAN, G. A., HANSON, P. & FINER, S. 2015. Socio-cultural influences on the behaviour of South Asian women with diabetes in pregnancy: qualitative study using a multi-level theoretical approach. *BMC Med,* 13, 120. | Other – expert subject knowledge in review team | Europe (UK) | Qualitative (interview study) | 45 women of Bangladeshi, Indian, Sri Lankan, or Pakistani origin with a history of diabetes in pregnancy, recruited from diabetes and antenatal services in two deprived London boroughs (aged 21 to 45 years) | To understand the multiple influences on behaviour (hence risks to metabolic health) of South Asian mothers and their unborn child, theorise how these influences interact and build over time, and inform the design of culturally congruent, multi-level interventions. |
| GUBRIUM, A., LECKENBY, D., HARVEY, M. W., MARCUS, B. H., ROSAL, M. C. & CHASAN-TABER, L. 2019. Perspectives of health educators and interviewers in a randomized controlled trial of a postpartum diabetes prevention program for Latinas: a qualitative assessment. *BMC HEALTH SERVICES RESEARCH,* 19. | Web of Science | Northern America (US) | Qualitative (focus group study) | 8 staff members (5 health educators and 3 health interviewers) | To describe the experiences of project staff in two RCTs of a postpartum lifestyle intervention to reduce risk factors for T2D in Latinas. |
| HAGA, S. B., BARRY, W. T., MILLS, R., SVETKEY, L., SUCHINDRAN, S., WILLARD, H. F. & GINSBURG, G. S. 2014. Impact of delivery models on understanding genomic risk for type 2 diabetes. *Public Health Genomics,* 17, 95-104. | EMBASE | Northern America (US) | Quantitative (randomised clinical trial) | 300 participants without T2D, recruited from Duke University and surrounding areas (aged 18 years and above) | To address the effectiveness of in-person versus online delivery models for understanding genomic risk and adoption of healthy behaviours. |
| HALL, R., SAUKKO, P. M., EVANS, P. H., QURESHI, N. & HUMPHRIES, S. E. 2007. Assessing family history of heart disease in primary care consultations: A qualitative study. *Family Practice,* 24, 435-442. | PsycINFO | Europe (UK) | Qualitative (semi-structured interviews) | 21 primary care patients and 7 primary care clinicians (2 practice nurses, 5 GPs) recruited from 4 practices in South West England | To examine how clinicians and patients understand and communicate family history in the context of CHD risk assessment in primary care. |
| HAYES, J. F., FOWLER, L. A., BALANTEKIN, K. N., ROTMAN, S. A., ALTMAN, M. & WILFLEY, D. E. 2021. Child and Family Predictors of Relative Weight Change in a Low-Income, School-Based Weight Management Intervention. *FAMILIES SYSTEMS & HEALTH,* 39, 316-326. | Web of Science | Northern America (US) | Quantitative (intervention study) | 145 children and families from an urban elementary school in the Midwest (children mean age 6.5 years) | To examines predictors of weight outcomes following the socioecological model in a school-based weight management intervention implemented in an elementary school serving primarily low-income, Black youth. |
| HEIDEMAN, W. H., DE WIT, M., MIDDELKOOP, B. J. C., NIERKENS, V., STRONKS, K., VERHOEFF, A. P. & SNOEK, F. J. 2012. DiAlert: a prevention program for overweight first degree relatives of type 2 diabetes patients: results of a pilot study to test feasibility and acceptability. *Trials,* 13, 178. | Ovid MEDLINE | Europe (the Netherlands) | Quantitative (pilot trial) | 21 participants with first- or second-degree relatives with T2D (mean age 47.9 years) | To assess fidelity, feasibility and acceptability prior to starting DiAlert, a targeted group-based intervention aimed to promote intrinsic motivation and action planning for lifestyle changes and weight loss in relatives of patients with T2D. |
| HICKEY, K. T., TAYLOR, J. Y., SCIACCA, R. R., ABOELELA, S., GONZALEZ, P., CASTILLO, C., HAUSER, N. & FRULLA, A. 2014. Cardiac genetic testing: a single-center pilot study of a Dominican population. *Hispanic health care international : the official journal of the National Association of Hispanic Nurses,* 12, 183-8. | Ovid MEDLINE | Northern America (US) | Mixed-methods (survey and interviews) | 31 participants recruited from the cardiovascular services and ICD clinic at New York-Presbyterian Hospital, Columbia University Medical Centre (mean age 42 years) | To evaluate the physiological well-being and perceived cardiac risk among Dominicans who underwent genetic testing. |
| HILBERT, A. 2013. Genetic determinism in weight bias reduction. *Obesity Facts,* 6, 49. | EMBASE | Europe (Germany) | Quantitative (intervention study) | 128 university students (mean age 23.01 years) and 128 individuals recruited from the community (mean age 35.31 years) | To 1) examine associations between genetic causal attributions, belief in genetic determinism, and stigmatizing attitudes (N = 432); 2) develop and pilot a brief, interactive stigma reduction intervention educating about gene x environment interactions in the aetiology of obesity within an RCT with N = 128 university students and 3) evaluate this intervention in an RCT in the general population (N = 128). |
| HILBERT, A., DIERK, J.-M., CONRADT, M., SCHLUMBERGER, P., HINNEY, A., HEBEBRAND, J. & RIEF, W. 2009. Causal attributions of obese men and women in genetic testing: implications of genetic/biological attributions. *Psychology & health,* 24, 749-61. | Ovid MEDLINE | Europe (Germany) | Quantitative (secondary analysis of intervention study) | 421 individuals with obesity, recruited from the community for a larger study on genetic testing and counselling of obesity (mean age 45.8 years) | To investigate genetic/biological attributions of obesity, their associations with a predisposition to obesity and their cross-sectional and longitudinal implications for weight regulation in individuals with obesity presenting for genetic testing and counselling. |
| HOEDEMAEKERS, E., JASPERS, J. P. C. & VAN TINTELEN, J. P. 2007. The influence of coping styles and perceived control on emotional distress in persons at risk for a hereditary heart disease. *American Journal of Medical Genetics, Part A,* 143, 1997-2005. | EMBASE | Europe (the Netherlands) | Quantitative (trial) | 114 participants at risk for different hereditary cardiac diseases (aged 18 years and above) | To investigate the influence of two coping styles (monitoring and blunting) and perceived control (health locus of control and mastery) on emotional stress in persons at risk of a hereditary cardiac disease. |
| HOEEG, D., CHRISTENSEN, U. & GRABOWSKI, D. 2020. Intra-familial health polarisation: how diverse health concerns become barriers to health behaviour change in families with preschool children and emerging obesity. *Sociology of health & illness,* 42, 1243-1258. | Ovid MEDLINE | Europe (Denmark) | Qualitative (workshops with families and professionals) | 12 families and 34 professionals recruited from a rural municipality in Denmark | To understand the role of family dynamics in families’ everyday health practices in a disadvantaged area. |
| HOLLANDS, G. J., FRENCH, D. P., GRIFFIN, S. J., PREVOST, A. T., SUTTON, S., KING, S. & MARTEAU, T. M. 2016. The impact of communicating genetic risks of disease on risk-reducing health behaviour: systematic review with meta-analysis. *BMJ,* 352, i1102. | Other – expert subject knowledge in review team | N/A | Systematic reviews | N/A | To assess the impact of communicating DNA based disease risk estimates on risk-reducing health behaviours and motivation to engage in such behaviours. |
| HOVICK, S. R., WILKINSON, A. V., ASHIDA, S., DE HEER, H. D. & KOEHLY, L. M. 2014. The impact of personalized risk feedback on Mexican Americans’ perceived risk for heart disease and diabetes. *Health education research,* 29, 222-34. | Ovid MEDLINE | Northern America (US) | Quantitative (cluster RCT) | 497 adults (comprising 162 households) recruited from The University of Texas MD Anderson Cancer Center’s Mexican American Cohort Study (mean age 41.2 years) | To examine Mexican Americans’ risk perceptions for heart disease and diabetes at baseline and following receipt of risk feedback based on family health history. |
| HULKOWER, R., DAVIS, N., SCHECHTER, C. & WALKER, E. 2015. Risk perception of obesity and fast food behavior among obese adults in primary care. *Endocrine Reviews,* 36. | EMBASE | Northern America (US) | Quantitative (cross-sectional survey) | 145 participants recruited from municipal hospital clinics in Bronx NYC, a largely Latino and Black population (mean age 49 years) | To determine the associations between BMI, calories purchased in fast food restaurants and risk perception among overweight and obese adults. |
| HUNT, A. V., HILTON, D. C. K., VERRALL, C. E., BARLOW-STEWART, K. K., FLEMING, J., WINLAW, D. S. & BLUE, G. M. 2020. “Why and how did this happen?”: development and evaluation of an information resource for parents of children with CHD. *Cardiology in the young,* 30, 346-352. | Ovid MEDLINE | Oceania (Australia) | Quantitative (cross-sectional survey) | 52 parents of children attending preadmission clinic for surgery (mean age 35.22 years) | To develop a brochure to determine whether an information resource could improve parents’ knowledge about CHD causation and inheritance and increase psychosocial functioning. |
| IMES, C. C. 2013. *Family history and cardiovascular disease risk in at-risk young adults: A pilot intervention study.* 74, ProQuest Information & Learning. | PsycINFO | Northern America (US) | Theses and dissertations | 15 asymptomatic, young adults with a family history of CVD (mean age 20.8 years) | To examine the short-term impact of a theoretically driven, educational intervention on perceived CVD risk and behavioural intention to change health-related behaviour to reduce CVD risk in asymptomatic young adults with a known family history of CVD. |
| IMES, C. C., DOUGHERTY, C. M., LEWIS, F. M. & AUSTIN, M. A. 2016. Outcomes of a Pilot Intervention Study for Young Adults at Risk for Cardiovascular Disease Based on Their Family History. *The Journal of cardiovascular nursing,* 31, 433-40. | Ovid MEDLINE | Northern America (US) | Quantitative (pilot feasibility study) | 15 asymptomatic, young adults with a family history of CVD (mean age 20.8 years) | To examine the short-term impact of a theoretically driven, educational intervention on perceived CVD risk and behavioural intention to change health-related behaviour to reduce CVD risk in asymptomatic young adults with a known family history of CVD. |
| IMES, C. C. & LEWIS, F. M. 2014. Family history of cardiovascular disease, perceived cardiovascular disease risk, and health-related behavior: a review of the literature. *J Cardiovasc Nurs,* 29, 108-29. | Other – reference chaining | N/A | Systematic reviews | N/A | To review and summarize the published research on the relationship between an family history of CVD, an individual's perceived risk, and health-related behaviour to make recommendations for clinical practice and future research. |
| IMES, C. C., NOVOSEL, L. M. & BURKE, L. E. 2016. Heart Disease Risk and Self-efficacy in Overweight and Obese Adults. *JNP-JOURNAL FOR NURSE PRACTITIONERS,* 12, 710-716. | Web of Science | Northern America (US) | Quantitative (cross-sectional survey) | 151 respondents recruited from a weight loss research registry (mean age 51.3 years) | To examine the associations between sociodemographic characteristics, CHD risk factors including family history, perceived CHD risk, and self-efficacy to eat a heart healthy low-fat, low-salt diet, in a sample of overweight and obese adults. |
| JAFAR-MOHAMMADI, B. & MCCARTHY, M. I. 2008. Genetics of type 2 diabetes mellitus and obesity - A review. *Annals of Medicine,* 40, 2-10. | Scopus | N/A | Systematic reviews | N/A | To review the genetics of T2D and obesity. |
| JAMES, K. M., COWL, C. T., TILBURT, J. C., SINICROPE, P. S., ROBINSON, M. E., FRIMANNSDOTTIR, K. R., TIEDJE, K. & KOENIG, B. A. 2011. Impact of direct-to-consumer predictive genomic testing on risk perception and worry among patients receiving routine care in a preventive health clinic. *Mayo Clinic proceedings,* 86, 933-40. | Ovid MEDLINE | Northern America (US) | Mixed-methods (trial and follow-up interviews) | 150 patients attending a preventive medicine clinic (aged 70 years and below) | To assess the impact of direct-to-consumer predictive genomic risk information on perceived risk and worry in the context of routine clinical care. |
| JEONG, S.-H. 2007. Effects of news about genetics and obesity on controllability attribution and helping behavior. *Health communication,* 22, 221-8. | Ovid MEDLINE | Asia (Korea) | Quantitative (cross-sectional survey) | 95 undergraduate students recruited from a psychology course at a large private university (mean age 20.3 years) | To test the effects of news stories that offer gene-based explanations of obesity compared to behaviour-based and complex (combining genetic and behavioural) explanations. |
| JOSLYN, M. R. & HAIDER-MARKEL, D. P. 2019. Perceived causes of obesity, emotions, and attitudes about Discrimination Policy. *Social Science & Medicine,* 223, 97-103. | PsycINFO | Northern America (US) | Quantitative (cross-sectional survey) | 1,596 adults for Study 1 and 2,250 adults for Study 2 | To examine perceived causes of obesity, emotional responses, and related policy implications. |
| KANE, M., THORNTON, J. & BIBBY, J. 2022. Building public understanding of health and health inequalities. *The Health Foundation*. | Other – reference chaining | N/A | Reports and policy documents | N/A | To explore the reasons behind public attitudes towards health and health inequalities. |
| KASHANI, M., ELIASSON, A., VERNALIS, M., COSTA, L. & TERHAAR, M. 2013. Improving assessment of cardiovascular disease risk by using family history: an integrative literature review. *The Journal of cardiovascular nursing,* 28, E18-27. | Ovid MEDLINE | N/A | Systematic reviews | N/A | To examine the state of evidence for the use of family history as a predictor in CVD risk stratification. |
| KAYANIYIL, S., ARDERN, C. I., WINSTANLEY, J., PARSONS, C., BRISTER, S., OH, P., STEWART, D. E. & GRACE, S. L. 2009. Degree and correlates of cardiac knowledge and awareness among cardiac inpatients. *Patient Education and Counseling,* 75, 99-107. | PsycINFO | Northern America (Canada) | Quantitative (cross-sectional survey) | 1,308 CHD inpatients (mean age 65.2 years) | To investigate the degree of CHD awareness as well as symptom, risk factor, and treatment knowledge in a broad sample of cardiac inpatients, and to examine its sociodemographic, clinical and psychosocial correlates. |
| KHALID, Y., MALINA, O., ROFIAH, A., LATINAH, M., THAHIRAHTUL, A. Z., ZARIDAH, M. S. & TAN, M. H. 1994. Disease and risk factor perception among patients with coronary artery disease in Kuala Terengganu. *The Medical journal of Malaysia,* 49, 205-8. | Ovid MEDLINE | Asia (Malaysia) | Quantitative (cross-sectional survey) | 100 patients with CHD attending the Physician Clinic, Kuala Terengganu General Hospital (mean age 60.3 years) | To explore CHD patients’ awareness of coronary risk factors and their perceptions of their disease. |
| KIM, J., CHOI, S., KIM, C. J., OH, Y. & SHINN, S. H. 2002. Perception of risk of developing diabetes in offspring of type 2 diabetic patients. *The Korean journal of internal medicine,* 17, 14-8. | Ovid MEDLINE | Asia (Korea) | Quantitative (cross-sectional survey) | 101 men without T2D, but with one or both parents having T2D (median age 22 years) | To investigate how the male offspring of patients with T2D assess their likelihood of developing diabetes. |
| KNOWLES, J., ZARAFSHAR, S., PAVLOVIC, A., GOLDSTEIN, B., KIERNAN, M., TSAI, S., MCCONNELL, M., ABSHER, D., ASHLEY, E., IOANNIDIS, J. & ASSIMES, T. 2016. Impact of a genetic risk score for coronary artery disease in reducing cardiovascular risk: A pilot randomized controlled study. *Circulation,* 134. | EMBASE | Northern America (US) | Quantitative (RCT) | 84 participants with moderate CAD risk (mean age 57 years in intervention arm and 58 years in control arm) | To test whether providing a genetic risk score of CAD would improve adherence to risk-reducing strategies. |
| LASCO, G., MENDOZA, J., RENEDO, A., SEGUIN, M. L., PALAFOX, B., PALILEO-VILLANUEVA, L. M., AMIT, A. M. L., DANS, A. L., BALABANOVA, D. & MCKEE, M. 2020. Nasa dugo(‘It’s in the blood’): lay conceptions of hypertension in the Philippines. *BMJ GLOBAL HEALTH,* 5. | Web of Science | Asia (the Philippines) | Qualitative (semi-structured interviews) | 33 patients with hypertension from low-income communities in Valenzuela City (urban) and Quezon Province (rural) (aged 35 to 70 years) | To determine what adult patients with hypertension in the Philippines attribute their condition to, how these views might be explained and what the implications are for hypertension management. |
| LEWIS, A. C. F., PEREZ, E. F., PRINCE, A. E. R., FLAXMAN, H. R., GOMEZ, L., BROCKMAN, D. G., CHANDLER, P. D., KERMAN, B. J., LEBO, M. S., SMOLLER, J. W., WEISS, S. T., BLOUT ZAWATKSY, C. L., MEIGS, J. B., GREEN, R. C., VASSY, J. L. & KARLSON, E. W. 2022. Patient and provider perspectives on polygenic risk scores: implications for clinical reporting and utilization. Genome Medicine, 14. | Other – expert subject knowledge in review team | Northern America (US) | Mixed-methods (survey and interviews) | 46 patients and primary care providers | To explore patient and primary care provider responses to PRS clinical reporting choice, which can then inform future implementation considerations. |
| LI, S. X., YE, Z., WHELAN, K. & TRUBY, H. 2016. The effect of communicating the genetic risk of cardiometabolic disorders on motivation and actual engagement in preventative lifestyle modification and clinical outcome: a systematic review and meta-analysis of randomised controlled trials. *Br J Nutr,* 116, 924-34. | Other – expert subject knowledge in review team | N/A | Systematic reviews | N/A | To investigate whether genetic risk communication affects motivation and actual behaviour change towards preventative lifestyle modification. |
| LIN, J., MARCUM, C. S., MYERS, M. F. & KOEHLY, L. M. 2017. Put the family back in family health history: A multiple-informant approach. *American Journal of Preventive Medicine,* 52, 640-644. | PsycINFO | Northern America (US) | Quantitative (secondary analysis of family history data) | 127 participants from 45 families of diverse backgrounds (mean age 47.83 years) | To examine whether family members have consistent perceptions of shared familial risk for four common chronic conditions (heart disease, T2D, high cholesterol, and hypertension) using a multiple-informant approach—and whether accounting for inconsistency in family health history reports leads to more accurate risk assessment. |
| LIN, J., MARCUM, C. S., WILKINSON, A. V. & KOEHLY, L. M. 2018. Developing Shared Appraisals of Diabetes Risk Through Family Health History Feedback: The Case of Mexican-Heritage Families. *Annals of behavioral medicine : a publication of the Society of Behavioral Medicine,* 52, 262-271. | Ovid MEDLINE | Northern America (US) | Quantitative (experimental study) | 222 parents (mean age 48.32 years) and 131 children (mean age 22.34 years) | To identify the optimal risk feedback approach that facilitates risk communication between parents and their adult children and helps them develop shared appraisals of family history of T2D. |
| LINDSAY, A. C., WALLINGTON, S. F., LEES, F. D. & GREANEY, M. L. 2018. Exploring How the Home Environment Influences Eating and Physical Activity Habits of Low-Income, Latino Children of Predominantly Immigrant Families: A Qualitative Study. *INTERNATIONAL JOURNAL OF ENVIRONMENTAL RESEARCH AND PUBLIC HEALTH,* 15. | Web of Science | Northern America (US) | Qualitative (focus group discussions) | 33 low-income Latino parents of preschool children (aged 2 to 5 years) | To explore low-income Latino parents’ beliefs, parenting styles, and parenting practices related to their children’s eating and physical activity behaviours while at home. |
| LIPPA, N. C. & SANDERSON, S. C. 2012. Impact of information about obesity genomics on the stigmatization of overweight individuals: An experimental study. *Obesity,* 20, 2367-2376. | PsycINFO | N/A | Quantitative (randomised online experiment) | 396 participants (mean age 42.7 years) | To examine the impact of genetic versus non-genetic information on obesity stigma among self-perceived non-overweight individuals. |
| MARKOWITZ, S. M., PARK, E. R., DELAHANTY, L. M., O’BRIEN, K. E. & GRANT, R. W. 2011. Perceived impact of diabetes genetic risk testing among patients at high phenotypic risk for type 2 diabetes. *Diabetes care,* 34, 568-73. | Ovid MEDLINE | Northern America (US) | Qualitative (interview study) | 22 overweight participants at high phenotypic risk for T2D (mean age 57.7 years) | To explore perceptions of diabetes genetic risk testing compared with currently available prediction using nongenetic risk factors (e.g., family history, abnormal fasting glucose, obesity). |
| MARTEAU, T. & LERMAN, C. 2001. Genetic risk and behavioural change. *BMJ (Clinical research ed.),* 322, 1056. | Other – expert subject knowledge in review team | N/A | Systematic reviews | N/A | To review the limited evidence concerning behavioural responses to genetic information on risk. |
| MARTEAU, T., SENIOR, V., HUMPHRIES, S. E., BOBROW, M., CRANSTON, T., CROOK, M. A., DAY, L., FERNANDEZ, M., HORNE, R., IVERSEN, A., JACKSON, Z., LYNAS, J., MIDDLETON-PRICE, H., SAVINE, R., SIKORSKI, J., WATSON, M., WEINMAN, J., WIERZBICKI, A. S., WRAY, R. & GENETIC RISK ASSESSMENT FOR, F. H. T. S. G. 2004. Psychological impact of genetic testing for familial hypercholesterolemia within a previously aware population: a randomized controlled trial. *Am J Med Genet A,* 128A, 285-93. | Other – expert subject knowledge in review team | Europe (UK) | Quantitative (RCT) | 341 families, comprising 341 hypercholesterolemia probands and 128 adult relatives (mean age ranging from 44.1 years to 56.2 years) | To investigate the psychological impact of using genetic testing to make or confirm a clinical diagnosis of hypercholesterolemia. |
| MARTEAU, T. M., KINMONTH, A. L., PYKE, S. & THOMPSON, S. G. 1995. READINESS FOR LIFE-STYLE ADVICE - SELF-ASSESSMENTS OF CORONARY RISK PRIOR TO SCREENING IN THE BRITISH FAMILY HEART-STUDY. *BRITISH JOURNAL OF GENERAL PRACTICE,* 45, 5-8. | Web of Science | Europe (UK) | Quantitative (cross-sectional survey) | 3,725 participants who accepted an invitation to attend health screening as part of the British family heart study (mean age 50.1 years for men and 47.8 years for women) | To document how attenders at primary care cardiovascular screening clinics perceived their risks of CHD prior to screening; the degree of similarity between perceived level of risk and an epidemiologically derived risk score; and the relative importance assigned to individual risk factors by subjects compared with those assigned by the risk score. |
| MARTEAU, T. M. & WEINMAN, J. 2006. Self-regulation and the behavioural response to DNA risk information: A theoretical analysis and framework for future research. *Social Science and Medicine,* 62, 1360-1368. | EMBASE | N/A | Systematic reviews | N/A | To explain and predict the  characteristics of risk information that are more and less likely to motivate behaviour change, drawing upon the CSM-SR. |
| MCVAY, M. A., BEADLES, C., WU, R., GRUBBER, J., COFFMAN, C. J., YANCY, W. S., REINER, I. L. & VOILS, C. I. 2015. Effects of provision of type 2 diabetes genetic risk feedback on patient perceptions of diabetes control and diet and physical activity self-efficacy. *Patient education and counseling,* 98, 1600‐1607. | Cochrane Central Register of Controlled Trials (CENTRAL) | Northern America (US) | Quantitative (secondary analysis of RCT) | 531 participants, predominantly Black American (mean age 54.0 years in intervention group and 54.9 years in control group) | To examine the effects of providing genetic risk feedback for T2D on perceived diabetes control and self-efficacy for diet and physical activity. |
| MEULENKAMP, T. M., TIBBEN, A., MOLLEMA, E. D., VAN LANGEN, I. M., WIEGMAN, A., DE WERT, G. M., DE BEAUFORT, I. D., WILDE, A. A. M. & SMETS, E. M. A. 2008. Predictive genetic testing for cardiovascular diseases: impact on carrier children. *American journal of medical genetics. Part A,* 146A, 3136-46. | Ovid MEDLINE | Europe (the Netherlands) | Qualitative (semi-structured interviews) | 33 children who tested positive for Long QT Syndrome (LQTS), Hypertrophic Cardiomyopathy (HCM), Familial Hypercholesterolemia (FH) and their parents (children aged 8 to 18 years) | To study the experiences of children identified by family screening who were found to be a mutation carrier for a genetic cardiovascular disease. |
| MIDDLEMASS, J. B., YAZDANI, M. F., KAI, J., STANDEN, P. J. & QURESHI, N. 2014. Introducing genetic testing for cardiovascular disease in primary care: a qualitative study. *The British journal of general practice : the journal of the Royal College of General Practitioners,* 64, e282-9. | Ovid MEDLINE | Europe (UK) | Qualitative (interview study) | 29 adults who consented to genetic testing after having had a conventional cardiovascular risk assessment (median age 59 years) | To explore how patients who have had a recent conventional cardiovascular risk assessment, perceive additional information from genetic testing for CHD. |
| MUNOZ, L. R., ETNYRE, A., ADAMS, M., HERBERS, S., WITTE, A., HORLEN, C., BAYNTON, S., ESTRADA, R. & JONES, M. E. 2010. Awareness of heart disease among female college students. *Journal of women’s health (2002),* 19, 2253-9. | Ovid MEDLINE | Northern America (US) | Quantitative (cross-sectional survey) | 320 women from a private university (mean age 23 years) | To evaluate the level of awareness and knowledge of heart disease in women among college students. |
| MURPHY, B., WORCESTER, M., HIGGINS, R., LE GRANDE, M., LARRITT, P. & GOBLE, A. 2005. Causal attributions for coronary heart disease among female cardiac patients. *Journal of Cardiopulmonary Rehabilitation,* 25, 135-145. | Scopus | Oceania (Australia) | Mixed-methods (survey and semi-structured interviews) | 260 women admitted to hospital after an acute myocardial infarction or for coronary artery bypass graft surgery (mean age 68.6 years) | To investigate causal attributions and their associations with actual risk profiles in female cardiac patients. |
| NISHIGAKI, M., KOBAYASHI, K., KATO, N., SEKI, N., YOKOMURA, T., YOKOYAMA, M. & KAZUMA, K. 2009. Preventive advice given by patients with type 2 diabetes to their offspring. *The British journal of general practice : the journal of the Royal College of General Practitioners,* 59, 37-42. | Ovid MEDLINE | Asia (Japan) | Quantitative (cross-sectional survey) | 221 parents with T2D who had offspring aged 20 to 49 years (mean age 64.2 years) | To investigate the conditions under which patients with T2D offer advice to their offspring and to assess the factors that facilitate advice giving. |
| NISHIGAKI, M., TOKUNAGA-NAKAWATASE, Y., NISHIDA, J. & KAZUMA, K. 2014. The effect of genetic counseling for adult offspring of patients with type 2 diabetes on attitudes toward diabetes and its heredity: a randomized controlled trial. *Journal of genetic counseling,* 23, 762-9. | Ovid MEDLINE | Asia (Japan) | Quantitative (RCT) | 145 healthy adults who have a family history of T2D in their first degree relatives (mean age 46.9 years in intervention group and 44.9 years in control group) | To investigate the effect of diabetes genetic counselling—based on the HBM—on attitudes toward diabetes and its heredity in relatives of patients with T2D. |
| O’DONOVAN, C. E., SKINNER, J. R. & BROADBENT, E. 2020. Perceptions of Risk of Cardiac Arrest in Individuals Living With a Cardiac Inherited Disease: Are the Doctor and the Patient on the Same Page? *Heart Lung and Circulation,* 29, 851-858. | EMBASE | Oceania (New Zealand) | Quantitative (postal survey) | 177 patients and clinicians (mean age 45 years) | To examine whether patients’ and clinician’s risk perceptions about cardiac inherited disease correlate—and factors associated with patient perceptions. |
| OGDEN, J., DALKOU, M., KOUSANTONI, M., VENTURA, S. S. & REYNOLDS, R. 2017. Body weight, the home environment, and eating behaviour across three generations of women: A quasi‐longitudinal study in four Mediterranean and non‐Mediterranean countries. *Australian Psychologist,* 52, 442-452. | PsycINFO | Europe (UK, Greece, Malta) and Oceania (Australia) | Quantitative (quasi‐longitudinal study) | 170 women coming from three generations of families (mean age 21.7 years for daughters, 51.1 years for mothers and 77.3 years for grandmothers) | To explore how changes in the home environment reflect body weight and eating behaviours in three generations of women across two non‐Mediterranean (UK and Australia) and two Mediterranean countries (Greece and Malta). |
| OLIVERI, S., FERRARI, F., MANFRINATI, A. & PRAVETTONI, G. 2018. A systematic review of the psychological implications of genetic testing: A comparative analysis among cardiovascular, neurodegenerative and cancer diseases. *Frontiers in Genetics,* 9. | EMBASE | N/A | Systematic reviews | N/A | To review the psychological implication of undergoing genetic testing for cardiovascular, neurodegenerative and cancer diseases. |
| OROM, H., SCHOFIELD, E., KIVINIEMI, M. T., WATERS, E. A. & HAY, J. L. 2021. Agency beliefs are associated with lower health information avoidance. *Health Education Journal,* 80, 272-286. | EMBASE | Northern America (US) | Quantitative (cross-sectional survey) | 1,605 participants recruited from an Internet survey panel (GfK KnowledgePanel) (mean age 46.93 years in Sample 1 and 47.32 years in Sample 2) | To explore the association between risk perception and health information avoidance. |
| OSUJI, N. A., OJO, O. S., MALOMO, S. O., SOGUNLE, P. T., EGUNJOBI, A. O. & ODEBUNMI, O. O. 2018. Relationship between glycemic control and perceived family support among people with type 2 diabetes mellitus seen in a rich kinship network in Southwest Nigeria. *FAMILY MEDICINE AND COMMUNITY HEALTH,* 6, 168-177. | Web of Science | Africa (Nigeria) | Quantitative (cross-sectional survey) | 316 adults with T2D who attended a medical outpatient clinic (mean age 60.96 years) | To investigate the relationship between glycaemic control and perceived family support among Nigerians with T2D. |
| PAPPA, E., KONTODIMOPOULOS, N., PAPADOPOULOS, A. A., PALLIKARONA, G., NIAKAS, D. & TOUNTAS, Y. 2009. Factors Affecting Use of Preventive Tests for Cardiovascular Risk among Greeks. *INTERNATIONAL JOURNAL OF ENVIRONMENTAL RESEARCH AND PUBLIC HEALTH,* 6, 2712-2724. | Web of Science | Europe (Greece) | Quantitative (cross-sectional survey) | 1,005 participants residing in urban and rural areas of the country (aged 18 years and above) | To investigate socio-demographic, self-perceived health, and health risk factors that determine the use of cardiovascular preventive tests (blood pressure, cholesterol and blood glucose). |
| PATEL, N. R. 2012. *The role of illness beliefs and social networks in South Asian people: A mixed-methods study.* | Other – reference chaining | Europe (UK) | Theses and dissertations | 67 South Asian participants with T1D or T2D (mean age 61 years) | To provide a better  understanding of diabetes management in people of South Asian origin in the UK. |
| PATEL, N. R., CHEW-GRAHAM, C., BUNDY, C., KENNEDY, A., BLICKEM, C. & REEVES, D. 2015. Illness beliefs and the sociocultural context of diabetes self-management in British South Asians: a mixed methods study. *BMC Fam Pract,* 16, 58. | Other – reference chaining | Europe (UK) | Mixed-methods (survey and interviews) | 67 South Asian participants with T1D or T2D (mean age 61 years) | To explore the influence of sociocultural  context on illness beliefs and diabetes self-management in British South Asians. |
| PELEG, O., HADAR, E. & COHEN, A. 2020. Individuals with type 2 diabetes: An exploratory study of their experience of family relationships and coping with the illness. *The Diabetes Educator,* 46, 83-93. | PsycINFO | Middle East (Israel) | Qualitative (interview study) | 32 Israeli Jewish and Arab individuals with T2D recruited from a community population (mean age 54.1 years) | To explore familial patterns that may be related to T2D and to patients’ ways of coping with the illness. |
| PERSKY, S., BOUHLAL, S., GOLDRING, M. R. & MCBRIDE, C. M. 2017. Beliefs about genetic influences on eating behaviors: Characteristics and associations with weight management confidence. *EATING BEHAVIORS,* 26, 93-98. | Web of Science | Northern America (US) | Quantitative (cross-sectional survey) | 261 participants (mean age 34 years) | To characterise healthy individuals’ beliefs about the notion that there are genetic influences on eating behaviour in comparison with beliefs about genetic influences on body weight. |
| PERSKY, S. & ECCLESTON, C. P. 2011. Impact of Genetic Causal Information on Medical Students’ Clinical Encounters with an Obese Virtual Patient: Health Promotion and Social Stigma. *ANNALS OF BEHAVIORAL MEDICINE,* 41, 363-372. | Web of Science | Northern America (US) | Quantitative (randomised experiment) | 110 third- and fourth-year medical students (mean age 26.22 years) | To explore whether information about the genetics of obesity reduces medical student stigmatisation of patients with obesity, and how it affects rates of health behaviour-related referral. |
| PERSKY, S., GOLDRING, M. R., EL-TOUKHY, S., FERRER, R. A. & HOLLISTER, B. 2019. Parental Defensiveness about Multifactorial Genomic and Environmental Causes of Children’s Obesity Risk. *Childhood obesity (Print),* 15, 289-297. | Ovid MEDLINE | Northern America (US) | Quantitative (randomised experiment) | 324 self-identified overweight adults recruited, with children aged 3 to 13 years (mean age 35.34 years to 37.16 years across conditions) | To examine the psychological consequences (in terms of risk perception and guilt) of parental exposure to genomics-oriented media reports about obesity risk among children. |
| PERSKY, S. & YAREMYCH, H. E. 2020. Parents’ genetic attributions for children’s eating behaviors: Relationships with beliefs, emotions, and food choice behavior. *APPETITE,* 155. | Web of Science | Northern America (US) | Quantitative (randomised experiment) | 190 parents with children aged 4 to 7 years (mean age 37.71 years) | To assess parental genetic attributions for their child's eating behaviour, and relationships between these attributions and self-efficacy, guilt, and feeding behaviours. |
| PETR, E. J., AYERS, C. R., PANDEY, A., DE LEMOS, J. A., POWELL-WILEY, T. M., KHERA, A., LLOYD-JONES, D. M. & BERRY, J. D. 2014. Perceived Lifetime Risk for Cardiovascular Disease (from the Dallas Heart Study). *AMERICAN JOURNAL OF CARDIOLOGY,* 114, 53-58. | Web of Science | Northern America (US) | Quantitative (cross-sectional analysis of participants in the Dallas Heart Study) | 2,998 participants who took part in a follow-up visit of the Dallas Heart Study (aged 30 to 65 years) | To determine the perception of lifetime risk for CVD by comparing Dallas Heart Study participants’ perceived lifetime risk with their predicted lifetime risk for CVD using a previously published algorithm. |
| PHG FOUNDATION 2019. Polygenic scores, risk and cardiovascular disease. | Other – expert subject knowledge in review team | N/A | Reports and policy documents | N/A | To the field of PRSs from the perspective of CVD prevention, to assess evidence and  readiness for clinical implementation. |
| PHG FOUNDATION 2021. Implementing polygenic scores for cardiovascular disease into NHS Health Checks. | Other – expert subject knowledge in review team | N/A | Reports and policy documents | N/A | To explore the changes needed to implement and deliver PRS analysis within  existing practice, using the NHS Health Check programme as an exemplar for early implementation. |
| PICCININO, L., GRIFFEY, S., GALLIVAN, J., LOTENBERG, L. D. & TUNCER, D. 2015. Recent Trends in Diabetes Knowledge, Perceptions, and Behaviors: Implications for National Diabetes Education. *Health education & behavior : the official publication of the Society for Public Health Education,* 42, 687-96. | Ovid MEDLINE | Northern America (US) | Reports and policy documents | 6,075 participants who engaged in three population-based National Diabetes Education Program National Diabetes Surveys at 2006, 2008, and 2011 (aged 35 years and above) | To examine trends in diabetes-related knowledge, perceptions, and behaviour among US adults with and without a diagnosis of diabetes and among subpopulations at risk. |
| PIERCE, M., HARDING, D., RIDOUT, D., KEEN, H. & BRADLEY, C. 2001. Risk and prevention of type II diabetes: Offspring’s views. *British Journal of General Practice,* 51, 194-199. | Scopus | Europe (UK) | Quantitative (cross-sectional survey) | 105 individuals with a parent affected by T2D (median age 38 years) | To explore beliefs about personal risk of diabetes and prevention in people with a parent with T2D. |
| PIERCE, M., HAYWORTH, J., WARBURTON, F., KEEN, H. & BRADLEY, C. 1999. Diabetes mellitus in the family: Perceptions of offspring’s risk. *Diabetic Medicine,* 16, 431-436. | Scopus | Europe (UK) | Quantitative (cross-sectional survey) | 213 patients with T2D (median age 67 years) | To explore the beliefs and concerns of people with T2D about their children’s risk of developing the disease and the possibilities for prevention. |
| PIERCE, M., RIDOUT, D., HARDING, D., KEEN, H. & BRADLEY, C. 2000. More good than harm: a randomised controlled trial of the effect of education about familial risk of diabetes on psychological outcomes. *The British journal of general practice : the journal of the Royal College of General Practitioners,* 50, 867-71. | Ovid MEDLINE | Europe (UK) | Quantitative (RCT) | 105 individuals with a parent affected by T2D (median age 35 years to 44 years across conditions) | To examine the cognitive and psychological effects of education about personal risk of T2D to individuals with a parent affected by T2D. |
| PIJL, M., TIMMERMANS, D. R., CLAASSEN, L., JANSSENS, A. C., NIJPELS, G., DEKKER, J. M., MARTEAU, T. M. & HENNEMAN, L. 2009. Impact of communicating familial risk of diabetes on illness perceptions and self-reported behavioral outcomes: a randomized controlled trial. *Diabetes Care,* 32, 597-9. | Other – reference chaining | Europe (the Netherlands) | Quantitative (RCT) | 118 participants identified to be at risk for T2D through a diabetes screening programme (mean age 67.1 years) | To assess the potential effectiveness of communicating familial risk of diabetes on illness perceptions and self-reported behavioural outcomes. |
| PODURI, A. & GRISSO, J. A. 1998. Cardiovascular risk factors in economically disadvantaged women: A study of prevalence and awareness. *Journal of the National Medical Association,* 90, 531-536. | PsycINFO | Northern America (US) | Quantitative (cross-sectional survey) | 99 women recruited from community sites in Philadelphia (mean age 37.6 years) | To examine the prevalence of cardiovascular risk factors among low-income women and assessed the level of awareness and attitudes about these risk factors in the community. |
| POLACSEK, M., ORR, J., O’BRIEN, L. M., ROGERS, V. W., FANBURG, J. & GORTMAKER, S. L. 2014. Sustainability of Key Maine Youth Overweight Collaborative Improvements: A Follow-Up Study. *CHILDHOOD OBESITY,* 10, 326-333. | Web of Science | Northern America (US) | Quantitative (quasi-experimental field trial) | 7 Maine Youth Overweight Collaborative sites and 2 control sites | To evaluate the effect of the Maine Youth Overweight Collaborative on provider knowledge, beliefs, practices, patient experience, and office systems in 2012, three years post-intervention. |
| POLLEY, B. A., JAKICIC, J. M., VENDITTI, E. M., BARR, S. & WING, R. R. 1997. The effects of health beliefs on weight loss in individuals at high risk for NIDDM. *Diabetes Care,* 20, 1533-1538. | Scopus | Northern America (US) | Quantitative (RCT) | 154 participants recruited with newspaper advertisements for a behavioural program to reduce the risk of developing T2D (mean age 45.7 years) | To determine whether perceived risk and other health beliefs held by individuals at high risk for developing T2D predict weight loss and behaviour change during a behavioural weight loss program to reduce the risk of T2D. |
| RAZALI, S., ISMAIL, Z., ABDULLAH, N. & NAWAWI, H. M. 2019. Illness Perception, Level of Education and Presence of Cardiovascular Disease among Patients with Familial Hypercholesterolaemia. *ENVIRONMENT-BEHAVIOUR PROCEEDINGS JOURNAL.* | Web of Science | Asia (Malaysia) | Quantitative (cross-sectional survey) | 100 participants, mainly from lower socioeconomic households (mean age 49.8 years) | To describe the illness perceptions of  patients with FH and investigate their associations with sociodemographic and illness-related factors. |
| REGO, S., DAGAN‐ROSENFELD, O., BIVONA, S. A., SNYDER, M. P. & ORMOND, K. E. 2019. Much ado about nothing: A qualitative study of the experiences of an average‐risk population receiving results of exome sequencing. *Journal of Genetic Counseling,* 28, 428-437. | PsycINFO | Northern America (US) | Qualitative (interview study) | 12 participants recruited from a longitudinal multi-omics profiling study that included exome sequencing (aged 45 to 74 years) | To assess 1) participants’ reasons for participating in the multi-omics profiling study; 2) perceptions of the risks and benefits of exome sequencing; 3) expectations for results and 4) reaction to the results. |
| REID, G., WALTER, F. & EMERY, J. 2011. Assessing the psychosocial impact of family history screening in the Australian primary care setting. *Familial Cancer,* 10, S86. | EMBASE | Oceania (Australia) | Qualitative (semi-structured telephone interviews) | 28 patients already enrolled in a family history screening study through their family physician (mean age 40.7 years) | To 1) explore the experience and impact of family history collection via a novel family history questionnaire and subsequent familial risk assessment and 2) assess the acceptability and feasibility of using the questionnaire in Australian primary care. |
| REYNA, V. F. 2008. A theory of medical decision making and health: Fuzzy trace theory. *Medical Decision Making,* 28, 850-865. | EMBASE | N/A | Commentaries and opinion pieces | N/A | To summarise the tenets of fuzzy trace theory, with respect to their relevance to health and medical decision-making. |
| ROBINSON, C. L., JOUNI, H., KRUISSELBRINK, T. M., AUSTIN, E. E., CHRISTENSEN, K. D., GREEN, R. C. & KULLO, I. J. 2016. Disclosing genetic risk for coronary heart disease: effects on perceived personal control and genetic counseling satisfaction. *Clinical genetics,* 89, 251-7. | Ovid MEDLINE | Northern America (US) | Quantitative (RCT) | 207 participants with no history of CHD or other atherosclerotic vascular diseases (aged 45 to 65 years) | To investigate whether disclosure of CHD genetic risk influences perceived personal control and genetic counselling satisfaction. |
| ROBINSON, C. L., JOUNI, H., KRUISSELBRINK, T. M., CHRISTENSEN, K. D., GREEN, R. C. & KULLO, I. J. 2014. The effect of disclosing genetic risk for coronary heart disease on perceived personal control and genetic counseling satisfaction: The MI-genes study. *Circulation,* 130. | EMBASE | Northern America (US) | Quantitative (RCT) | 207 participants (mean age 58.8 years) | To investigate whether disclosure of CHD genetic risk influences perceived personal control and genetic counselling satisfaction in the myocardial infarction genes (MI-GENES) study. |
| RODRIGUES, A. & CARRINGTON, M. 2017. Relationship between cardio-metabolic disease risk and health beliefs, perceptions and behaviours: A regional perspective. *Heart Lung and Circulation,* 26, S231. | EMBASE | Oceania (Australia) | Conference papers and proceedings | 277 patients with metabolic syndrome (aged 40 to 70 years) | To explore if one’s sense of self-efficacy, locus-of-control and perception of CVD risk can influence health behaviours and mediate cardiometabolic risk. |
| SANDERSON, S. C., DIEFENBACH, M. A., ZINBERG, R., HOROWITZ, C. R., SMIRNOFF, M., ZWEIG, M., STREICHER, S., JABS, E. W. & RICHARDSON, L. D. 2013. Willingness to participate in genomics research and desire for personal results among underrepresented minority patients: a structured interview study. *J Community Genet,* 4, 469-82. | Other – expert subject knowledge in review team | Northern America (US) | Mixed-methods (survey and interview study) | 205 patients in an inner-city hospital outpatient clinic (aged 22 to 85 years) | To examine willingness to participate in genomics research on four complex conditions (obesity, cancer, heart disease and T2D) among a sample of underrepresented minority patients, including their reasons for or against that willingness, and their desire for personal results from it. |
| SANDERSON, S. C., PERSKY, S. & MICHIE, S. 2010. Psychological and behavioral responses to genetic test results indicating increased risk of obesity: does the causal pathway from gene to obesity matter? *Public health genomics,* 13, 34-47. | Ovid MEDLINE | Northern America (US) | Quantitative (experimental study) | 191 participants, recruited mostly through a university setting (mean age 29.2 years) | To investigate whether people respond differently to a personal test result indicating increased risk of obesity based on a genetic or non-genetic biomarker which is described as exerting its obesogenic effect through an eating or a metabolic causal pathway. |
| SCALZI, L. V., BALLOU, S. P., PARK, J. Y., REDLINE, S. & KIRCHNER, H. L. 2008. Cardiovascular disease risk awareness in systemic lupus erythematosus patients. *Arthritis and rheumatism,* 58, 1458-64. | Ovid MEDLINE | Northern America (US) | Quantitative (cross-sectional survey) | 226 patients diagnosed with systemic lupus erythematosus (mean age 45.2 years) | To identify factors associated with patients’ recognition of systemic lupus erythematosus as an independent risk factor for CVD and their perception of their personal CVD risk. |
| SCHNEIDER, K. I. & SCHMIDTKE, J. 2014. Patient compliance based on genetic medicine: A literature review. *Journal of Community Genetics,* 5, 31-48. | EMBASE | N/A | Systematic reviews | N/A | To review studies on patient compliance after genetic risk assessment, focusing on conditions where secondary or tertiary preventive options exist, namely cancer syndromes (BRCA-related cancer, HNPCC/colon cancer), hemochromatosis, thrombophilia, smoking cessation, and obesity. |
| SCOLLAN-KOLIOPOULOS, M. 2005. *Type 2 diabetes illness representation, self-care, and multigenerational legacies of diabetes: Three reports.* 66, ProQuest Information & Learning. | PsycINFO | Northern America (US) | Theses and dissertations | 123 volunteers with T2D and a family history of diabetes | To develop a way to measure recollections about diabetes of family members and to estimate the associations between these recollections and participants’ own experience of diabetes. |
| SEABORN, C., SUTHER, S., LEE, T., KIROS, G. E., BECKER, A., CAMPBELL, E. & COLLINS-ROBINSON, J. 2016. Utilizing Genomics through Family Health History with the Theory of Planned Behavior: Prediction of Type 2 Diabetes Risk Factors and Preventive Behavior in an African American Population in Florida. *PUBLIC HEALTH GENOMICS,* 19, 69-80. | Web of Science | Northern America (US) | Quantitative (cross-sectional survey) | 394 African Americans recruited from faith-based entities  located in North Florida (aged 18 to 78 years) | To assess to what extent African Americans’ knowledge and awareness of family health history and related risk factors of developing T2D influence their likelihood of adopting a preventive behaviour. |
| SEABORN, C. A. E. 2016. *Introducing genomics via family history utilizing the theory of planned behavior: An assessment of type 2 diabetes mellitus risk factors and its influence on healthy behavior in an African American population in Florida.* 76, ProQuest Information & Learning. | PsycINFO | Northern America (US) | Theses and dissertations | 394 African Americans recruited from faith-based entities  located in North Florida (aged 18 to 78 years) | To assess to what extent African Americans’ knowledge and awareness of family health history and related risk factors of developing T2D influence their likelihood of adopting a preventive behaviour. |
| SEGAL, M. E., POLANSKY, M. & SANKAR, P. 2007. Predictors of uptake of obesity genetic testing among affected adults. *HUMAN GENETICS,* 120, 641-652. | Web of Science | Northern America (US) | Qualitative (focus groups) | 64 respondents recruited from notices in physician’s offices, diabetes centres, newspapers, and calls from a market research company (aged 18 to 70 years) | To explore characteristics of individuals who would be most likely to obtain future genetic testing for obesity. |
| SEGAL, M. E., SANKAR, P. & REED, D. R. 2004. Research issues in genetic testing of adolescents for obesity. *NUTRITION REVIEWS,* 62, 307-320. | Web of Science | N/A | Commentaries and opinion pieces | N/A | To examine issues related to the eventual likelihood of genetic tests for obesity targeted to adolescents: family involvement; comprehension of the test’s meaning; how knowledge of genetic status may affect psychological adaptation; minors’ ability to control events; parental/child autonomy; ability to make informed medical decisions; self-esteem; unclear distinctions between early/late onset for this condition; and social stigmatisation. |
| SENIOR, V. & MARTEAU, T. M. 2007. Causal attributions for raised cholesterol and perceptions of effective risk-reduction: Self-regulation strategies for an increased risk of coronary heart disease. *Psychology & Health,* 22, 699-717. | PsycINFO | Europe (UK) | Quantitative (cross-sectional survey) | 317 participants clinically diagnosed with FH (mean age 54.6 years) | To investigate self-regulation strategies in 317 people with FH. |
| SENIOR, V., WEINMAN, J. & MARTEAU, T. M. 2002. The influence of perceived control over causes and responses to health threats: A vignette study. *British Journal of Health Psychology,* 7, 203-211. | EMBASE | Europe (UK) | Quantitative (experimental study) | 108 nursing students (mean age 23.81 years) | To test the hypothesis that a disease caused by controllable, compared with uncontrollable, factors is more likely to result in negative emotions and less likelihood of disclosure to significant others. |
| SHERMAN, K., CAMERON, L., BROWN, P. & MARTEAU, T. 2009. Effect of worry and monitoring processing style on cognitive and affective responses to genetic risk information. *Psycho-Oncology,* 18, S139-S140. | EMBASE | Europe (UK) and Oceania (Australia, New Zealand) | Conference papers and proceedings | 752 healthy individuals recruited from Australia, New Zealand and the UK to participate in an online study | To evaluate the role of pre-manipulation worry and monitoring processing style as predictors and moderators of responses to genetic risk information. |
| SHERMAN, K., SHAW, L.-K., CHAMPION, K., CALDEIRA, F. & MCCASKILL, M. 2015. The effect of disease risk probability and disease type on interest in clinic-based versus direct-to-consumer genetic testing services. *Journal of behavioral medicine,* 38, 706-14. | Ovid MEDLINE | Oceania (Australia) | Quantitative (experimental study) | 309 participants recruited through a pool of first year Psychology students at Macquarie University, and general community members (mean age 27.54 years) | To asses the effect of disease risk probability and disease type on interest in clinic-based versus direct-to-consumer genetic testing services. |
| SHILOH, S., DEHEER, H. D., PELEG, S., HENSLEY ALFORD, S., SKAPINSKY, K., ROBERTS, J. S. & HADLEY, D. W. 2015. The impact of multiplex genetic testing on disease risk perceptions. *Clinical Genetics,* 87, 117-123. | EMBASE | Northern America (US) | Quantitative (cross-sectional survey) | 216 healthy adults (aged 25 to 40 years) | To assess the effects of multiplex genetic testing on disease risk perceptions. |
| SHILOH, S., WADE, C. H., ROBERTS, J. S., ALFORD, S. H. & BIESECKER, B. B. 2013. Associations between risk perceptions and worry about common diseases: a between- and within-subjects examination. *Psychology & health,* 28, 434-49. | Ovid MEDLINE | Northern America (US) | Quantitative (secondary analysis of the Multiplex Initiative) | 294 people recruited through the Multiplex Initiative, in which a genetic susceptibility test for 8 common diseases was offered to healthy adults (mean age 34.61 years) | To test the relationships between worry and perceptions of likelihood and severity (two indicators of risk perception) across eight common diseases, and to examine contributions of individual and disease variability in worry and risk perceptions. |
| STOL, D. M., HOLLANDER, M., DAMMAN, O. C., NIELEN, M. M. J., BADENBROEK, I. F., SCHELLEVIS, F. G. & DE WIT, N. J. 2020. Mismatch between self-perceived and calculated cardiometabolic disease risk among participants in a prevention program for cardiometabolic disease: a cross-sectional study. *BMC public health,* 20, 740. | Ovid MEDLINE | Europe (the Netherlands) | Quantitative (secondary analysis of RCT data) | 7,547 participants recruited from primary care (aged 45 to 70 years) | To assess the impact of communicating an individualised cardiometabolic disease risk score on perceived risk and to identify risk factors and demographic characteristics associated with risk perception among high-risk participants of a prevention program for cardiometabolic diseases. |
| STUTTGEN, K., PACYNA, J., KULLO, I. & SHARP, R. 2020. Neutral, negative, or negligible? Changes in patient perceptions of disease risk following receipt of a negative genomic screening result. *Journal of Personalized Medicine,* 10. | EMBASE | Northern America (US) | Quantitative (cross-sectional survey) | 1,442 participants who had hyperlipidaemia and/or colon polyps (mean age 60.8 years) | To determine whether patients had lower disease risk perception after receiving a negative genomic screening result, to describe factors that might be associated with a tendency to downgrade perceived disease risk in light of receiving negative results, and to investigate the effects of such changes on risk perception, including whether participants who lowered disease risk perception shared theirs result with family members. |
| SULTAN, N. & SWINGLEHURST, D. 2021. Self-Management in Older Pakistanis Living With Multimorbidity in East London. *Qual Health Res,* 31, 2111-2122. | Other – expert subject knowledge in review team | Europe (UK) | Qualitative (biographical narrative interviews) | 15 first-generation Pakistani migrants living with multimorbidity in East London (aged 53 to 87 years) | To explore how older British Pakistani people experience multimorbidity (defined as the coexistence of two or more medical conditions) and engage with self-management within the context of their life histories and relationships. |
| SWEET, K., GORDON, E. S., STURM, A. C., SCHMIDLEN, T. J., MANICKAM, K., TOL, A. E., KELLER, M. A., STACK, C. B., FELIPE GARCÍA-ESPAÑA, J., BELLAFANTE, M., TAYAL, N., EMBI, P., BINKLEY, P., HERSHBERGER, R. E., SADEE, W., CHRISTMAN, M. & MARSH, C. 2014. Design and implementation of a randomized controlled trial of genomic counseling for patients with chronic disease. *Journal of Personalized Medicine,* 4, 1-19. | EMBASE | Northern America (US) | Quantitative (RCT) | 210 participants enrolled in a clinical setting (mean age 58.1 years) | To describe the development and implementation of an RCT to investigate the impact of genomic counselling on a cohort of patients with heart failure or hypertension, managed at a large academic medical centre. |
| TANG, J. W., CAMERON, K. A., PUMARINO, J., PEACEMAN, A. & ACKERMANN, R. T. 2013. Perceived risk for type 2 diabetes among women with a history of gestational diabetes. *Journal of General Internal Medicine,* 28, S144. | EMBASE | Northern America (US) | Mixed-methods (survey and interview study) | 74 women who were diagnosed with GDM during a recent pregnancy and were within 18 months of delivery (mean age 33.8 years) | To explore whether women with a history of GDM are aware of their increased risk for developing T2D and the extent to which they seek primary care follow-up after delivery. |
| TURNWALD, B. P., GOYER, J. P., BOLES, D. Z., SILDER, A., DELP, S. L. & CRUM, A. J. 2019. Learning one’s genetic risk changes physiology independent of actual genetic risk. *Nature human behaviour,* 3, 48-56. | Ovid MEDLINE | Northern America (US) | Quantitative (experimental study) | 271 participants genotyped for two experiments (mean age 25.3 years) | To test whether merely learning one’s genetic risk for disease alters one’s actual risk by making people more likely to exhibit the expected changes in gene-related physiology, behaviour, and subjective experience. |
| VAN ESCH, S. C. M., CORNEL, M. C., GEELHOED-DUIJVESTIJN, P. H. L. M. & SNOEK, F. J. 2012. Family communication as strategy in diabetes prevention: an observational study in families with Dutch and Surinamese South-Asian ancestry. *Patient education and counseling,* 87, 23-9. | Ovid MEDLINE | Europe (the Netherlands) | Quantitative (cross-sectional survey) | 468 patients with T2D of Dutch and Surinamese origin (mean age 67.5 years and 58.1 years, respectively) | To explore the possibility of utilising family communication as a diabetes prevention strategy, specifically targeting high-risk families with South Asian ancestry in the Netherlands. |
| VAN ESCH, S. C. M., NIJKAMP, M. D., CORNEL, M. C. & SNOEK, F. J. 2012. Patients’ intentions to inform relatives about Type 2 diabetes risk: the role of worry in the process of family risk disclosure. *Diabetic medicine : a journal of the British Diabetic Association,* 29, e461-7. | Ovid MEDLINE | Europe (the Netherlands) | Quantitative (cross-sectional survey) | 546 patients with T2D recruited from 4 primary care practices (mean age 63.6 years) | To further our understanding of factors that influence the decisional process of familial risk disclosure in patients with diabetes. |
| VAN ESCH, S. C. M., NIJKAMP, M. D., CORNEL, M. C. & SNOEK, F. J. 2014. Illness representations of type 2 diabetes patients are associated with perceptions of diabetes threat in relatives. *Journal of health psychology,* 19, 358-68. | Ovid MEDLINE | Europe (the Netherlands) | Quantitative (cross-sectional survey) | 546 patients with T2D recruited from 4 primary care practices (mean age 63.6 years) | To explore whether and to what extent patients’ illness representations are related to three determinants in family risk disclosure—patients’ perceptions of T2D risk in relatives, their worries about relatives developing the disease and their beliefs with regard to the possibility of T2D prevention in relatives. |
| VASSY, J. L., O’BRIEN, K. E., WAXLER, J. L., PARK, E. R., DELAHANTY, L. M., FLOREZ, J. C., MEIGS, J. B. & GRANT, R. W. 2012. Impact of Literacy and Numeracy on Motivation for Behavior Change After Diabetes Genetic Risk Testing. *MEDICAL DECISION MAKING,* 32, 606-615. | Web of Science | Northern America (US) | Quantitative (cross-sectional analysis of RCT) | 175 patients at high phenotypic risk for T2D, recruited for a clinical trial of diabetes genetic risk testing (mean age 58 years) | To investigate the association of health literacy, genetic literacy, and health numeracy with patient responses to diabetes genetic risk. |
| VOILS, C. I., COFFMAN, C. J., GRUBBER, J. M., EDELMAN, D., SADEGHPOUR, A., MACIEJEWSKI, M. L., BOLTON, J., CHO, A., GINSBURG, G. S. & YANCY, W. S., JR. 2015. Does Type 2 Diabetes Genetic Testing and Counseling Reduce Modifiable Risk Factors? A Randomized Controlled Trial of Veterans. *Journal of general internal medicine,* 30, 1591-8. | Ovid MEDLINE | Northern America (US) | Quantitative (RCT) | 601 patients recruited from primary care panels at the Durham Veterans Affairs Medical Centre and two satellite clinics (mean age 54.1 years) | To examined the clinical utility of supplementing T2D risk counselling with genetic test results and counselling. |
| VORNANEN, M., AKTAN-COLLAN, K., HALLOWELL, N., KONTTINEN, H. & HAUKKALA, A. 2019. Lay Perspectives on Receiving Different Types of Genomic Secondary Findings: a Qualitative Vignette Study. *Journal of genetic counseling,* 28, 343-354. | Ovid MEDLINE | Europe (Finland) | Qualitative (focus group study) | 23 participants recruited via an announcement in the Helsinki area Metro newspaper (mean age 49 years) | To explore how lay people react to different types of hypothetical genomic secondary findings. |
| VORNANEN, M., KONTTINEN, H., KAARIAINEN, H., MANNISTO, S., SALOMAA, V., PEROLA, M. & HAUKKALA, A. 2016. Family history and perceived risk of diabetes, cardiovascular disease, cancer, and depression. *Preventive medicine,* 90, 177-83. | Ovid MEDLINE | Europe (Finland) | Quantitative (cross-sectional analysis of the FINRISK 2007 study) | 6,258 participants recruited from the FINRISK 2007 study sample (mean age 50.8 years) | To examine how family history relates to perceived risk of T2D, CVD, cancer, and depression, and whether these associations are independent of or moderated by sociodemographic factors, health behaviour/weight status (smoking, alcohol consumption, physical activity, BMI), or depressive symptoms. |
| VU, A. V., TURK, N., DURU, O. K., MANGIONE, C., PANCHAL, H., AMAYA, S. A., NORRIS, K. C. & MOIN, T. 2020. The impact of type 2 diabetes mellitus risk perception on adoption of preventive strategies in women with a history of gestational diabetes. *Diabetes,* 69. | EMBASE | Northern America (US) | Quantitative (cross-sectional survey) | 264 women with a history of GDM who had not progressed to T2D (aged 18 to 50 years) | To identify predictors of T2D risk perception in women with GDM and to determine if their level of risk perception impacts adoption of evidence-based health behaviours for T2DM prevention. |
| WALTER, F. M. & EMERY, J. 2005. ‘Coming down the line’—patients’ understanding of their family history of common chronic disease. *Annals of family medicine,* 3, 405-14. | Ovid MEDLINE | Europe (UK) | Qualitative (semi-structured interviews) | 30 patients recruited from general practice who had a family history of cancer, heart disease, or diabetes (aged 20 years and above) | To explore how patients in primary care understand and come to terms with their family history of cancer, heart disease, or diabetes and how family history might affect consultations about disease risk and management. |
| WALTER, F. M., EMERY, J., BRAITHWAITE, D. & MARTEAU, T. M. 2004. Lay understanding of familial risk of common chronic diseases: A systematic review and synthesis of qualitative research. *Annals of Family Medicine,* 2, 583-594. | EMBASE | N/A | Systematic reviews | N/A | To review and synthesise the qualitative literature exploring understanding about familial risk held by persons with a family history of cancer, coronary artery disease, and T2D. |
| WANG, C., GONZALEZ, R. & MERAJVER, S. D. 2004. Assessment of genetic testing and related counseling services: current research and future directions. *SOCIAL SCIENCE & MEDICINE,* 58, 1427-1442. | Web of Science | N/A | Commentaries and opinion pieces | N/A | To provide an overview of the potential outcomes of genetic services and highlight constructs for future research in this area. |
| WANG, C., SEN, A., RUFFIN, M. T., NEASE, D. E., GRAMLING, R., ACHESON, L. S., O’NEILL, S. M. & RUBINSTEIN, W. S. 2012. Family history assessment: Impact on disease risk perceptions. *American Journal of Preventive Medicine,* 43, 392-398. | EMBASE | Northern America (US) | Quantitative (cluster-randomised trial) | 3,786 patients recruited from 41 primary care practices among 13 states between 2005 and 2007 (mean age 50.6 years) | To examine the impact of FHITr on modifying disease risk perceptions, particularly among those who initially underestimated their risk for certain diseases. |
| WATERS, E. A., ACKERMAN, N. & WHEELER, C. S. 2019. Cognitive and Affective Responses to Mass-media Based Genetic Risk Information in a Socio-demographically Diverse Sample of Smokers. *JOURNAL OF HEALTH COMMUNICATION,* 24, 700-710. | Web of Science | Northern America (US) | Quantitative (secondary analysis of RCT) | 392 smokers recruited from public locations (mean age 44.5 years) | To examine how informing adult smokers about the genetic basis for nicotine addiction influences smoking-related health cognitions and affect and whether responses vary by socio-demographics or genetics beliefs. |
| WAXLER, J. L., O’BRIEN, K. E., DELAHANTY, L. M., MEIGS, J. B., FLOREZ, J. C., PARK, E. R., POBER, B. R. & GRANT, R. W. 2012. Genetic counseling as a tool for type 2 diabetes prevention: a genetic counseling framework for common polygenetic disorders. *Journal of genetic counseling,* 21, 684-91. | Ovid MEDLINE | Northern America (US) | Quantitative (intervention study) | 72 participants at phenotypically high-risk for T2D (mean age 58.1 years) | To describe a framework for developing the diabetes genetic risk counselling intervention created for the Genetic Counselling/Lifestyle Change for Diabetes Prevention Study Study and also provide some initial feedback from study participants based on their experience with this counselling intervention. |
| WENZEL, L. & GLANZ, K. 2004. Behavioral Aspects of Genetic Risk for Disease: Cancer Genetics as a Prototype for Complex Issues in Health Psychology. *In:* BOLL, T. J., FRANK, R. G., BAUM, A. & WALLANDER, J. L. (eds.) *Handbook of clinical health psychology: Volume 3. Models and perspectives in health psychology.* Washington, DC: American Psychological Association. | PsycINFO | N/A | Book chapter | N/A | To illuminate burgeoning research areas in which health psychologists provide an important contribution as the sciences of genetics, epidemiology, and public health unite toward common goals. |
| WESSEL, J., GUPTA, J. & DE GROOT, M. 2016. Factors Motivating Individuals to Consider Genetic Testing for Type 2 Diabetes Risk Prediction. *PloS one,* 11, e0147071. | Ovid MEDLINE | Northern America (US) | Quantitative (cross-sectional survey) | 598 adults recruited from public libraries, online registry and a safety net hospital emergency department (mean age 37 years) | To identify attitudes and perceptions of willingness to participate in genetic testing for T2D risk prediction in the general population. |
| WIDEN, E., JUNNA, N., RUOTSALAINEN, S., SURAKKA, I., MARS, N., RIPATTI, P., PARTANEN, J. J., ARO, J., MUSTONEN, P., TUOMI, T., PALOTIE, A., SALOMAA, V., KAPRIO, J., PARTANEN, J., HOTAKAINEN, K., POLLANEN, P. & RIPATTI, S. 2022. How Communicating Polygenic and Clinical Risk for Atherosclerotic Cardiovascular Disease Impacts Health Behavior: an Observational Follow-up Study. *Circ Genom Precis Med,* 15, e003459. | Other – expert subject knowledge in review team | Europe (Finland) | Quantitative (intervention study) | 7,342 individuals in the population-based GeneRISK-study cohort (mean age 56 years) | To test how returning personal risk information about atherosclerotic CVD with an interactive web-tool impacted on participants’ health behaviour. |
| WIJDENES, M., HENNEMAN, L., QURESHI, N., KOSTENSE, P. J., CORNEL, M. C. & TIMMERMANS, D. R. M. 2013. Using web-based familial risk information for diabetes prevention: a randomized controlled trial. *BMC public health,* 13, 485. | Ovid MEDLINE | Europe (the Netherlands) | Quantitative (RCT) | 1,174 healthy adults with a BMI ≥ 25 (aged 35 to 65 years) | To determine if diabetic familial risk information by using a web-based tool leads to improved self-reported risk-reducing behaviour among individuals with a diabetic family history, without causing false reassurance among those without a family history. |
| WIJDENES-PIJL, M., DONDORP, W. J., TIMMERMANS, D. R. M., CORNEL, M. C. & HENNEMAN, L. 2011. Lay perceptions of predictive testing for diabetes based on DNA test results versus family history assessment: a focus group study. *BMC PUBLIC HEALTH,* 11. | Web of Science | Europe (the Netherlands) | Qualitative (focus group interviews) | 45 individuals with and without a family history of diabetes (aged 35 to 70 years) | To assess lay perceptions of issues related to predictive genetic testing for multifactorial diseases |
| WRIGHT, A. J., SUTTON, S. R., HANKINS, M., WHITWELL, S. C. L., MACFARLANE, A. & MARTEAU, T. M. 2012. Why does genetic causal information alter perceived treatment effectiveness? An analogue study. *British journal of health psychology,* 17, 294-313. | Ovid MEDLINE | Europe (UK) | Quantitative (experimental study) | 647 adults recruited from shopping centres (mean age 39.3 years) | To establish which beliefs mediate the effect of genetic causal information on perceived effectiveness of treatments for various common diseases. |
| WU, R. R., MYERS, R. A., HAUSER, E. R., VORDERSTRASSE, A., CHO, A., GINSBURG, G. S. & ORLANDO, L. A. 2017. Impact of Genetic Testing and Family Health History Based Risk Counseling on Behavior Change and Cognitive Precursors for Type 2 Diabetes. *Journal of genetic counseling,* 26, 133-140. | Ovid MEDLINE | Northern America (US) | Quantitative (RCT) | 391 patients without diabetes (mean age 50.9 years in intervention group and 49.3 years in control group) | To determine the impact of T2D family health history and genetic risk counselling on behaviour and its cognitive precursors. |
| YANG, K., BANIAK, L. M., IMES, C. C., CHOI, J. & CHASENS, E. R. 2018. Perceived Versus Actual Risk of Type 2 Diabetes by Race and Ethnicity. *DIABETES EDUCATOR,* 44, 269-277. | Web of Science | Northern America (US) | Quantitative (cross-sectional survey) | 10,999 participants (aged 20 years and above) | To examine associations between perceived risk and actual risk of type 2 diabetes by race and/or ethnicity. |
| YOUNG-HYMAN, D., HERMAN, L. J., SCOTT, D. L. & SCHLUNDT, D. G. 2000. Care giver perception of children’s obesity-related health risk: A study of African American families. *OBESITY RESEARCH,* 8, 241-248. | Web of Science | Northern America (US) | Quantitative (cross-sectional survey) | 111 families screened for participation in a diabetes prevention study | To examine care giver perception of children's weight-related health risk in African American families. |
| ZLOT, A. I., BLAND, M. P., SILVEY, K., EPSTEIN, B., MIELKE, B. & LEMAN, R. F. 2009. Influence of Family History of Diabetes on Health Care Provider Practice and Patient Behavior Among Nondiabetic Oregonians. *PREVENTING CHRONIC DISEASE,* 6. | Web of Science | Northern America (US) | Quantitative (secondary analysis of the 2005 Oregon Behavioural Risk Factor Surveillance System) | 6,039 participants without diabetes (aged 18 years and above) | To evaluate, among patients without diabetes, associations between family history of diabetes and patients’ reports of health care provider practices, patients’ perceived risk of developing diabetes, and patients’ behaviours associated with diabetes. |
| ZLOT, A. I., VALDEZ, R., HAN, Y., SILVEY, K. & LEMAN, R. F. 2010. Influence of family history of cardiovascular disease on clinicians’ preventive recommendations and subsequent adherence of patients without cardiovascular disease. Public health genomics, 13, 457-66. | Ovid MEDLINE | Northern America (US) | Quantitative (secondary analysis of the 2005 Oregon Behavioural Risk Factor Surveillance System) | 2,566 participants without CVD (mean age 45.2 years) | To examines associations between family history of CVD and 1) preventive practices and recommendations issued by clinicians; 2) patients’ perceived risk for developing CVD; 3) patients’ adoption of preventive and screening behaviours and 4) presence of risk factors for CVD among persons without CVD. |
